# Longitudinal proteomics defines stage-specific molecular signatures in Guillain-Barré syndrome

**DOI:** 10.64898/2026.05.23.26353948

**Authors:** Roger Collet-Vidiella, Paula Villatoro-González, Cinta Lleixà, Marta Caballero-Ávila, Clara Tejada-Illa, Elba Pascual-Goñi, Tania Mederer-Fernández, Paula Llarch, Juan Castilla-Silgado, Alberto de Lorenzo, Lara Panicot-Buj, Gemma Riesco-Navarro, Helena Codes, María José Sedano-Tous, Carlos Casasnovas, Gerardo Gutiérrez-Gutiérrez, Julio Pardo-Fernández, Álvaro Carbayo, Eduard Gallardo, Ana Vesperinas, Laura Llansó, David Reyes-Leiva, Elena Cortés-Vicente, Thais Armengué, Judit Llanos-Ramos, Álvaro García-Osuna, Jesús M. Martín-Campos, Elena Muiño, Israel Fernandez-Cadenas, Lorena Martín-Aguilar, Luis Querol, SPAiN Consortium

## Abstract

Guillain-Barré syndrome is an acute immune-mediated polyradiculoneuropathy with heterogeneous outcomes and limited molecular biomarkers for diagnosis, disease monitoring, and prognosis. To elucidate the circulating proteomic profile of this disorder and identify candidate biomarkers associated with disease activity and recovery, we measured over 6,500 proteins using an aptamer-based proteomic platform. We analysed paired, longitudinal sera from 20 patients at disease onset and one-year follow-up, alongside 15 healthy controls. Unbiased differential protein abundance and gene-set enrichment analyses were performed. Candidate proteins were validated using conventional immunoassays in a cohort including healthy and disease controls.

We identified 39 differentially abundant proteins between the acute and recovery phases and 248 proteins altered in acute Guillain-Barré syndrome compared to controls. The acute phase was characterised by a marked enrichment in systemic immune cascades and muscle sarcomere proteins, alongside a significant depletion of axonal adhesion molecules. Serum amyloid A1 (SAA1) emerged as the most strongly increased protein in the acute phase. Validation through independent immunoassays confirmed robust serum amyloid A elevations at disease onset relative to the one-year recovery phase, healthy controls, and relevant post-infectious and neuromuscular disease controls (acute disseminated encephalomyelitis and myasthenia gravis), underscoring a peripheral nerve-specific inflammatory response. Furthermore, unexpected elevations of cardiac troponin T (cTnT) were observed at disease onset. Clinical validation using high-sensitivity assays demonstrated that cTnT exceeded the diagnostic 99^th^ percentile upper reference limit in 25.5% of acute Guillain-Barré syndrome patients. A similarly high frequency of elevation in the myasthenia gravis disease control group (42.1%) suggests these increases predominantly reflect neuromuscular damage rather than myocardial injury. Finally, Mendelian randomisation provided causal genetic evidence linking specific systemic proteins to disease susceptibility, identifying robust roles for SERPING1 (plasma protease C1 inhibitor), CNDP1 (an antioxidant protein), and CRISPLD2 (a lipopolysaccharide-binding protein that regulates endotoxin function).

Together, this comprehensive proteomic characterisation reveals distinct, stage-specific molecular signatures in Guillain-Barré syndrome. Importantly, it suggests SAA1 as a robust marker of acute peripheral nerve inflammation and challenges the conventional interpretation of elevated cTnT in severe neuropathies and neuromuscular disorders. Furthermore, this work provides a novel dataset to explore future targeted therapeutic development in Guillain-Barré syndrome.

## Introduction

Guillain-Barré syndrome is an acute, severe, immune-mediated polyradiculoneuropathy that constitutes the leading cause of acute flaccid paralysis worldwide.^1^ It typically follows an infectious trigger and presents with rapidly progressive weakness, variable sensory symptoms, and autonomic dysfunction.^1^ Although immunomodulatory therapies such as intravenous immunoglobulin and plasma exchange, together with advances in modern critical care, have significantly improved outcomes, many patients still require intensive care or mechanical ventilation during the acute phase, and a considerable proportion are left with long-term disability.

The pathophysiology of Guillain-Barré syndrome involves a complex interplay between innate and adaptive immune responses directed against peripheral nerves and nerve roots. Yet, much of the mechanistic understanding is derived from animal models or cross-sectional human studies,^1,2^ and longitudinal analyses in well-characterized cohorts remain scarce. As a result, the molecular signatures that define disease onset, progression, and recovery are still incompletely understood.

Serum neurofilament light chain (sNfL) levels are markedly increased in Guillain-Barré syndrome and have emerged as a promising prognostic biomarker, particularly for predicting long-term functional outcome.^3–5^ However, the limited specificity of sNfL for peripheral axonal injury may restrict its clinical utility.^6,7^ Additional candidate biomarkers reflecting peripheral nerve axonal or myelin damage have recently been proposed,^8,9^ but these require further validation, and robust biomarkers that can support early diagnosis, monitor disease activity and recovery, or assess treatment response are still lacking.^10^ Importantly, Guillain-Barré syndrome offers a unique model for fluid biomarker discovery in autoimmune neuropathies, as its abrupt onset produces substantial tissue injury, and its monophasic course allows patients to serve as their own biological controls once recovery is underway. Using follow-up samples from the same individual decreases background heterogeneity, thereby maximising the signal-to-noise ratio for discovering acute-phase specific proteins.

High-throughput proteomic technologies offer a powerful resource for characterising systemic biological changes in Guillain-Barré syndrome and for identifying circulating proteins with diagnostic or prognostic potential. Among these platforms, the SomaScan^®^ assay uses Slow Off-rate Modified Aptamers (SOMAmer reagents) to quantify thousands of proteins with high sensitivity and specificity, enabling broad and unbiased profiling of the circulating proteome. This technology has already generated mechanistic insights and potential biomarkers in several neurological and inflammatory conditions, including Alzheimer’s disease, amyotrophic lateral sclerosis, and post-infectious syndromes such as long COVID.^11–13^

Here, we applied the 7k SomaScan^®^ platform to longitudinal serum samples from a multicentre, well-phenotyped cohort of patients with Guillain-Barré syndrome collected at disease onset and at one-year follow-up. Our objectives were to: (i) identify differentially abundant proteins in Guillain-Barré syndrome; (ii) define the biological pathways associated with these changes using gene-set enrichment analysis; (iii) evaluate circulating proteins associated with long-term prognosis; (iv) investigate the causal effects of candidate proteins on Guillain-Barré susceptibility via Mendelian randomisation (MR); and (v) validate candidate biomarkers using conventional immunoassay platforms in a cohort including both healthy and disease controls.

By combining large-scale proteomic profiling with longitudinal sampling and orthogonal validation, this study aims to delineate the systemic molecular signatures of Guillain-Barré syndrome and to identify circulating proteins with potential utility as biomarkers of disease activity, recovery, and clinical outcome.

## Materials and methods

### Population and study design

This study utilised a longitudinal cohort design for biomarker discovery, followed by a cross-sectional case-control phase for biomarker validation.

#### Discovery phase

Twenty patients fulfilling the diagnostic criteria of the European Academy of Neurology/Peripheral Nerve Society for Guillain-Barré syndrome from whom pre-treatment baseline and one-year follow-up samples were available were included in the SomaScan^®^ discovery analysis.^10^ All patients underwent a structured neurological assessment by a trained neurologist, and serum samples were obtained at disease onset prior to initiation of immunomodulatory treatment. Follow-up evaluations and a second blood draw were performed 52 weeks after onset (one-year follow-up).

This multicentre cohort included patients from five hospitals in Spain between February 2014 and March 2023. Serum samples from 15 age- and sex-matched healthy controls were collected at Hospital de la Santa Creu i Sant Pau and Hospital Sant Joan de Déu from individuals without active or recent acute illness. Participants were predominantly Caucasian, and ethnicity was not systematically recorded. Given the exploratory nature of this high-throughput proteomic study and the absence of pre-existing effect size estimates for circulating proteins in Guillain-Barré syndrome, a formal a priori sample size calculation was not performed. Instead, the study size was determined by the availability of high-quality paired longitudinal sera and pragmatic resource constraints.

#### Validation phase

Serum samples from patients with Guillain-Barré syndrome, together with healthy and disease controls, were analysed using three independent immunoassay platforms to validate candidate proteins identified in the SomaScan^®^ analysis. Candidate selection was based on statistical significance (adjusted *P*-values), fold-change magnitude, biological plausibility, and clinical availability.

Serum amyloid A (SAA) concentrations were initially measured using a Meso Scale Discovery electrochemiluminescence immunoassay in paired samples from patients with Guillain-Barré syndrome collected during the acute phase and at one-year follow-up (*n* = 21; comprising 14 samples also included in the SomaScan^®^ analysis and 7 additional patients included due to insufficient remaining sample volume from the original discovery cohort), as well as from healthy controls (*n* = 22).

Subsequently, ELISAs were performed to further evaluate candidate proteins in an expanded cohort comprising acute-phase samples from patients with Guillain-Barré syndrome (*n* = 22; 13 also analysed by SomaScan^®^), patients with myasthenia gravis positive for anti-acetylcholine receptor antibodies (treatment-naïve *n* = 20; relapse *n* = 2), patients with acute disseminated encephalomyelitis (*n* = 14), and healthy controls (*n* = 22). Samples from Guillain-Barré syndrome and healthy controls were used to validate all candidate proteins, while myasthenia gravis and acute disseminated encephalomyelitis samples were analysed exclusively for SAA to assess disease specificity. Myasthenia gravis served as a prototypical autoimmune neuromuscular disorder and acute disseminated encephalomyelitis as a representative post-infectious demyelinating disease not affecting the peripheral nervous system. All samples were collected at Hospital de la Santa Creu i Sant Pau, except for acute disseminated encephalomyelitis cases, which were recruited at Hospital Sant Joan de Déu.

High-sensitivity cardiac troponin T (hs-cTnT) was measured in samples from patients with acute Guillain-Barré syndrome (*n* = 51; 3 also included in the SomaScan^®^ analysis), healthy controls (*n* = 26), myasthenia gravis patients (treatment-naïve *n* = 17; relapse *n* = 2), and multiple sclerosis patients (*n* = 20).

### Serum processing and SomaScan^®^ proteomics

Following venous blood sampling, serum was aliquoted and stored at −80°C until analysis. Proteomic profiling was performed using the SomaScan^®^ 7k Assay (v4.1, SomaLogic Inc.). The assay uses 7335 modified single-stranded DNA aptamers (SOMAmer^®^ reagents), of which 7289 target 6596 unique human proteins, along with 46 internal controls, as previously described.^11,14^ Protein-aptamer complexes were quantified by fluorescence hybridization to DNA microarrays. Laboratory personnel were blinded to the clinical diagnoses.

Only aptamers annotated as human proteins were included. Measurements that failed manufacturer-defined quality control criteria were excluded, resulting in 6939 retained analytes. Signal intensities were median-normalised by SomaLogic to correct for assay-intrinsic variation, and values were log_2_-transformed prior to downstream analysis. Technical reproducibility and full quality control metrics are provided in the Supplementary Material, Supplementary Fig. 1 and 2.

### SomaScan^®^ data analysis and gene set enrichment analysis

Differential protein abundance across diagnostic groups (acute Guillain-Barré syndrome, recovery-phase Guillain-Barré syndrome at one-year follow-up, and healthy controls) was assessed using the *limma* framework (Linear Models for Microarray Data) in R.^15^ Paired linear models including patient ID as a blocking factor were used to compare acute and recovery samples. Comparisons versus healthy controls used unpaired models adjusted for age and sex. Moderated t-statistics were computed using *limma’*s empirical Bayes method, and *P*-values were corrected using the Benjamini-Hochberg false discovery rate (FDR). Post-hoc diagnostic checks confirmed the normality of residuals for the candidate proteins selected for validation (SAA1, ACTN2, and TNNI2; Shapiro-Wilk test *P* > 0.05), supporting the validity of the linear model assumptions for these analytes.

Proteins were considered differentially abundant if they showed an absolute log_2_ fold-change (logCFC) ≥ 1 and FDR < 0.05. For proteins with multiple aptamers, the probe with the largest absolute t-statistic was retained. This selection process resulted in a final analytical dataset of 6408 unique human proteins mapped to Entrez Gene IDs. Phase-specific proteins were defined by requiring consistent directionality across relevant contrasts (e.g., acute > control and acute > recovery for acute-specific). Complementing this unbiased approach, we assessed the abundance of candidate proteins previously reported as biomarkers in peripheral neuropathies. Given their established relevance, these candidates were evaluated based on statistical significance (FDR < 0.05) independent of the strict fold-change we applied for novel discovery. These included BDNF,^16^ the contactin family (CNTN1-6),^17,18^ dynactins,^19^ ENO2,^16^ GFAP,^16^ GLRX,^19^ IRAK4,^16^ MAPT,^16^ MMP-9,^20^ NCAM1 and NCAM2,^16^ neurofilaments (NEFH, NEFL),^4,16,21^ NGF and NGFR,^16^ NT5C3A,^19^ PRX,^9^ S100A9,^16^ SPP1,^16^ SUGT1,^19^ and TMPRSS5.^22^

For pathway analysis, proteins were ranked by moderated t-statistic and subjected to gene-set enrichment analysis using *fgsea* in R, incorporating gene sets from the MSigDB Hallmark, Reactome, and Gene Ontology Biological Process collections.^23–27^ Enriched pathways (FDR < 0.05) were visualised in Cytoscape (v3.10.3) using the EnrichmentMap (v3.5.0) and AutoAnnotate (v1.5.2).^28^

### Prognostic analysis

Associations between protein abundance measured during the acute phase of Guillain–Barré syndrome and functional outcome at one-year follow-up, defined as the ability to run, were assessed using the *limma* framework in R. Linear models adjusted for age were fitted for each protein, and moderated t-statistics were computed using *limma*’s empirical Bayes approach, followed by FDR multiple comparisons correction. To evaluate the prognostic predictive value of the proteomic data, elastic net regression was applied to identify proteins jointly predictive of outcome.

### Mendelian Randomisation Analyses

To investigate the potential causal effects of circulating proteins on Guillain-Barré syndrome susceptibility, we performed a two-sample MR analysis. We evaluated the 248 candidate proteins identified as differentially abundant in the acute phase relative to healthy controls. As exposures, we used genome-wide association studies (GWAS) of plasma protein levels measured by the SomaScan platform, obtained from the publicly available deCODE genetics repository. Of the 248 proteins, 188 had a GWAS summary in this repository, and only 82 showed *cis*-protein quantitative trait loci (*cis*-pQTLs). As an outcome, we used the GWAS for acute infective polyneuritis/Guillain-Barré syndrome in the UK Biobank European ancestry cohort (174 cases and 420,299 non-cases). All MR analyses were executed in R using the *TwoSampleMR* package. The inverse-variance weighted (IVW) method was employed as the primary analytical approach, with MR-Egger, weighted median (WM), pleiotropy residual sum and outlier test (MR-PRESSO), and adjusted profile score (MR-RAPS) as robust methods. We also performed sensitivity analysis to assess the robustness of the results, and colocalization to provide complementary evidence of association. A detailed description of the methodology is provided in the Supplementary Material.

### Protein quantifications by conventional immunoassays

Laboratory personnel performing the Meso Scale Discovery immunoassay, ELISAs, and the hs-cTnT electrochemiluminescence immunoassay (ECLIA) were blinded to the clinical diagnoses.

SAA concentrations were first quantified using the Meso Scale Discovery V-PLEX Human SAA kit on a MESO QuickPlex SQ 120MM Analyzer (Meso Scale Discovery, Rockville, MD, USA) at the Fundació barcelonaβeta Brain Research Center, following the manufacturer’s protocol. The intra-plate coefficient of variation (CV) was 3.6% and the inter-plate CV was 15.5%.

Subsequently, SAA, α-actinin-2 (ACTN2), and troponin I, fast skeletal muscle (TNNI2), were quantified using ELISA kits (Invitrogen^™^ KHA0011 for SAA; MyBioSource MBS2500935 for ACTN2; MyBioSource MBS927961 for TNNI2). Serum samples were thawed on ice, centrifuged to remove debris, and analysed in duplicate. Absorbance was read at 450 nm, and concentrations were calculated from standard curves using four-parameter logistic regression. For SAA, the mean intra-plate CV was 2.5% (all <15%), and the inter-plate CV was 13.6%.

An automated ECLIA on the Roche Cobas® e801 platform (Roche Diagnostics GmbH, Mannheim, Germany) was used to measure hs-cTnT. The measuring range was 3–10,000 ng/L. A concentration of >14 ng/L, representing the 99th percentile upper reference limit for a healthy population,^29^ was predefined as the threshold for elevation.

### Statistical Analysis

Descriptive statistics are shown as the mean ± standard deviation, median with interquartile range, or absolute frequency (percentage), as appropriate. For the validation phase, continuous biomarker concentrations were log_10_-transformed primarily for data visualisation. Homoscedasticity was evaluated using Levene’s test, and the normality of residuals was assessed using the Shapiro-Wilk test. Because parametric assumptions were not fully satisfied, non-parametric testing was employed. Cross-sectional multi-group comparisons were conducted using the Kruskal-Wallis test, with Dunn’s post-hoc tests for pairwise comparisons. Differences between two groups for unpaired and paired continuous variables were evaluated using the Mann-Whitney *U* test and the Wilcoxon signed-rank test, respectively. Categorical variables were analysed using Fisher’s exact test, with pairwise post-hoc comparisons. To adjust for multiple testing, the Benjamini-Hochberg false discovery rate method was applied to all post-hoc pairwise comparisons. A two-sided *P* < 0.05 was considered statistically significant. All analyses and data visualisation were performed in R (version 4.4.1; RStudio) using the *SomaDataIO*, *limma, dplyr, fgsea*, *pheatmap*, *stringr, tibble, readr, readxl, msigdbr*, *ggplot2, viridis, TwoSampleMR, patchwork,* and *ggrepel* packages.^15,23^ Reporting followed STROBE (Strengthening the Reporting of Observational Studies in Epidemiology) guidelines.^30^

### Ethics

The study protocol was approved by the Ethics Committee of Hospital de la Santa Creu i Sant Pau (IIBSP-NAI-2022-88). All participants provided written informed consent.

## Results

### Clinical profile of the exploratory Guillain-Barré syndrome cohort

Twenty patients with Guillain-Barré syndrome and 15 healthy controls were included in the SomaScan^®^ discovery analysis. The mean age of patients was 56.8 years (50% female), whereas controls had a mean age of 46.1 years and were predominantly men (53.3%). Most patients exhibited the sensorimotor variant (70%) and demyelinating features on nerve conduction studies (70%). The median baseline and maximum Guillain-Barré syndrome Disability Scale was 4. A preceding infection, most commonly respiratory, was reported in 80% of patients, and anti-GM1 IgG antibodies were identified in 25%. Additional demographic and clinical characteristics are presented in Table 1 and Supplementary Table 1.

**Table 1.** Demographic and clinical characteristics of the exploratory cohort.

| Baseline characteristics | Guillain-Barré syndrome | Healthy controls |
| --- | --- | --- |
| n | 20 | 15 |
| Age (years), mean ± SD | 56.8 ± 17.1 | 46.1 ± 25.4 |
| Sex, n (%) |  |  |
| Female | 10 (50) | 7 (46.7) |
| Male | 10 (50) | 8 (53.3) |
| Time from onset (days), median (IQR) | 3 (2-4) | NA |
| Initial GDS, median (IQR) | 4 (3-4) | NA |
| Ability to run at 52 weeks, n (%) | 15 (75) | NA |

GDS: Guillain-Barré Syndrome Disability Scale; IQR: interquartile range; NA: not applicable; SD: standard deviation.

### Acute Guillain-Barré syndrome serum exhibits a distinct proteomic signature

Pairwise comparisons between acute and recovery samples revealed 16 proteins elevated and 23 reduced in the acute phase (Fig. 1A, Supplementary Table 2). Relative to healthy controls, 248 proteins were differentially abundant in acute Guillain-Barré syndrome (187 increased, 61 decreased; Fig. 1B, Supplementary Table 3). Among these, four exhibited an acute-specific pattern: three were elevated and one reduced (Table 2). Fig. 1D shows the relative abundance of the leading candidate proteins across each diagnostic category.

**Figure 1.**
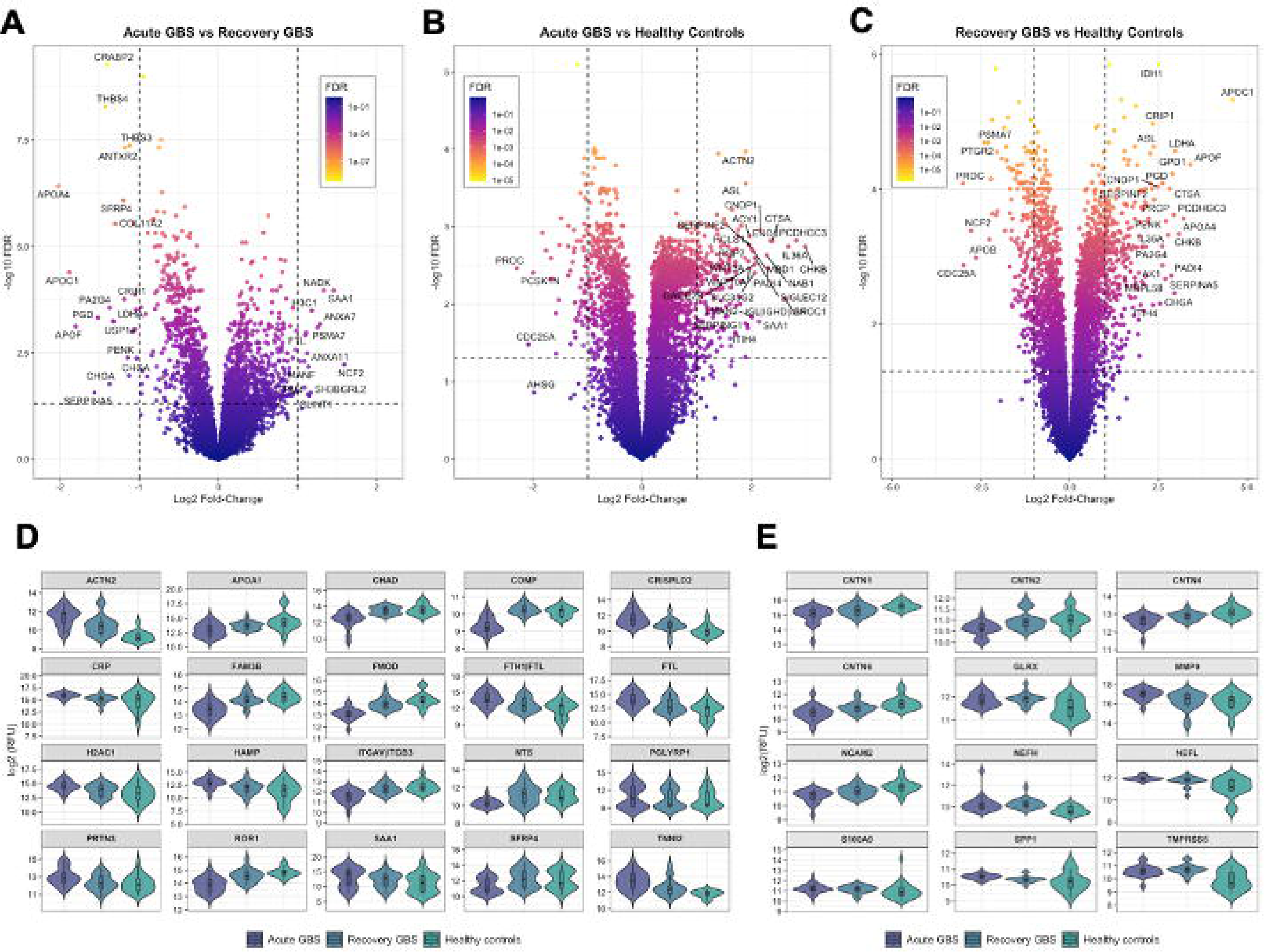
SomaScan discovery phase results. **(A–C)** Volcano plots illustrating differentially abundant proteins. Comparisons shown are: **(A)** acute Guillain-Barré syndrome compared to one-year follow-up, **(B)** acute Guillain-Barré syndrome compared to healthy controls, and **(C)** Guillain-Barré syndrome at one-year follow-up compared to healthy controls. **(D)** Violin plots with overlaid boxplots depicting the relative abundance of selected proteins across diagnostic subgroups. Displayed proteins represent those with the largest absolute log□(fold-change) in acute Guillain-Barré syndrome compared to one-year follow-up among proteins significantly altered in both contrasts. **(E)** Violin plots with overlaid boxplots showing the relative abundance of previously reported peripheral nerve biomarkers. Only prior biomarkers showing a statistically significant difference (false discovery rate < 0.05) between acute Guillain-Barré syndrome and healthy controls in our analysis are pictured. For all boxplots, the centre line represents the median, box limits represent the interquartile range, and whiskers extend to the most extreme data points within 1.5 times the interquartile range. For **D** and **E** proteins are ordered alphabetically, and the y-axes are scaled individually for each protein to optimize visualization. ACTN2, actinin alpha 2; APOA1, apolipoprotein A-I; CHAD, chondroadherin; CNTN, contactin; COMP, cartilage oligomeric matrix protein; CRISPLD2, cysteine-rich secretory protein LCCL domain-containing 2; CRP, C-reactive protein; FAM3B, family with sequence similarity 3 member B; FMOD, fibromodulin; FTH1, ferritin heavy chain 1; FTL, ferritin light chain; GBS, Guillain-Barré syndrome; GLRX, glutaredoxin-1; H2AC1, H2A clustered histone 1; HAMP, hepcidin antimicrobial peptide; ITGAV, integrin subunit alpha V; ITGB3, integrin subunit beta 3; MMP9, matrix metalloproteinase-9; NCAM, neural cell adhesion molecule; NEFH, neurofilament heavy chain; NEFL, neurofilament light chain; NTS, neurotensin; PGLYRP1, peptidoglycan recognition protein 1; PRTN3, proteinase 3; RFU, relative fluorescence units; ROR1, receptor tyrosine kinase-like orphan receptor 1; S100A9, calgranulin B; SAA1, serum amyloid A1; SFRP4, secreted frizzled-related protein 4; SPP1, osteopontin; TMPRSS5, transmembrane protease serine 5; TNNI2, troponin I2 fast skeletal type. Alt text: Five graphs labelled A to E illustrating proteomic discovery results. A, B, and C are volcano plots plotting statistical significance against fold-change to show differentially abundant proteins across the different comparisons. D is a set of violin plots with overlaid boxplots comparing the relative abundance of selected novel candidate proteins across the three diagnostic subgroups. E is a set of violin plots with overlaid boxplots showing the relative abundance of previously reported peripheral nerve biomarkers across the same subgroups.

**Table 2.**
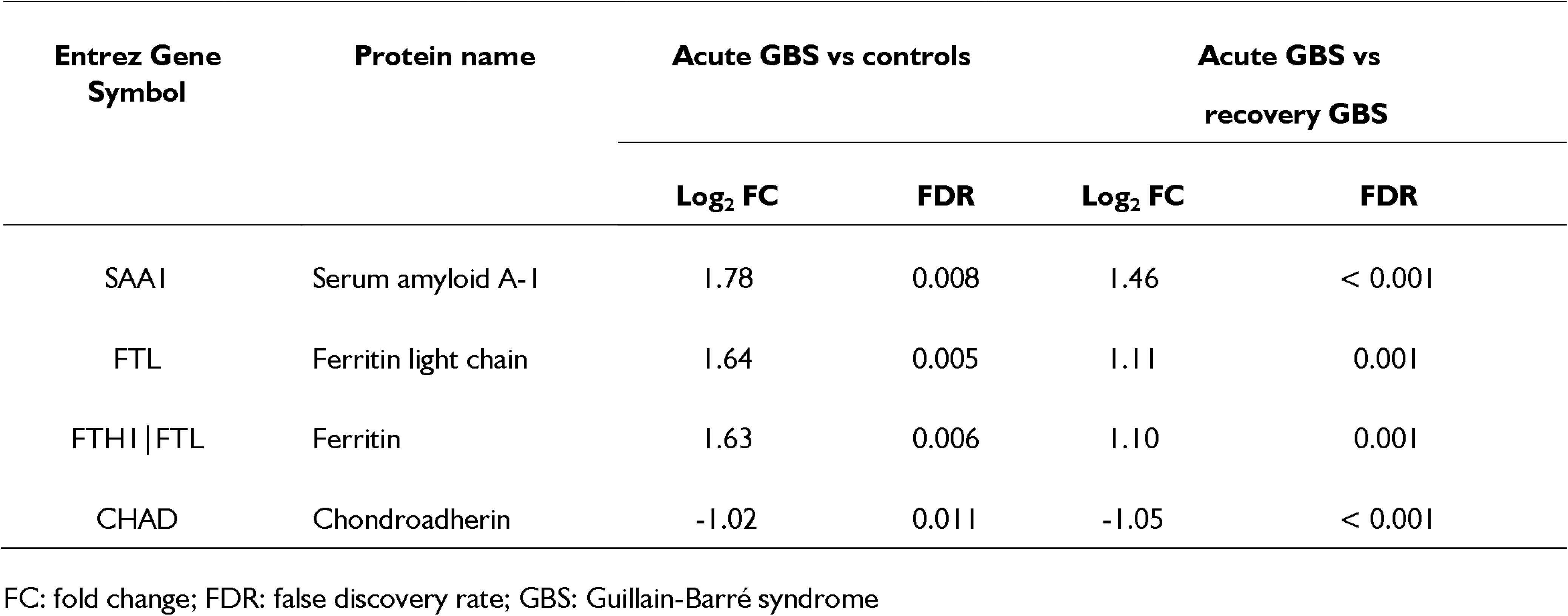
Acute-specific differentially abundant proteins in Guillain-Barré syndrome.

### Recovery-phase serum displays its own molecular phenotype

At one-year follow-up, 252 proteins were increased and 169 decreased compared with healthy controls (Fig. 1C, Supplementary Table 4). Thirty-two proteins showed a recovery-specific pattern, including 20 elevated and 12 reduced relative to both acute Guillain-Barré syndrome and controls. The ten most differentially abundant proteins compared to controls are summarised in Table 3, with the complete list in Supplementary Table 5.

**Table 3.** Recovery-specific differentially abundant proteins in Guillain-Barré Syndrome.

| Entrez Gene Symbol | Protein name | Recovery GBS vs controls |  | Recovery GBS vs acute GBS |  |
| --- | --- | --- | --- | --- | --- |
|  |  | Log<sub>2</sub> FC | FDR | Log<sub>2</sub> FC | FDR |
| APOC1 | Apolipoprotein C-1 | 4.59 | < 0.001 | 1.88 | < 0.001 |
| APOA4 | Apolipoprotein A-IV | 3.21 | < 0.001 | 2.02 | < 0.001 |
| APOF | Apolipoprotein F | 3.40 | < 0.001 | 1.80 | < 0.001 |
| SERPINA5 | Plasma serine protease inhibitor | 2.95 | 0.003 | 1.56 | 0.027 |
| LDHA | L-lactate dehydrogenase A chain | 2.97 | < 0.001 | 1.33 | < 0.001 |
| PGD | 6-phosphogluconate dehydrogenase; decarboxylating | 2.62 | < 0.001 | 1.52 | < 0.001 |
| PA2G4 | Proliferation-associated protein 2G4 | 2.61 | < 0.001 | 1.38 | < 0.001 |
| CHGA | Chromogranin-A | 2.54 | 0.005 | 1.38 | 0.017 |
| PENK | Proenkephalin-A | 2.71 | < 0.001 | 1.15 | 0.004 |
| NCF2 | Neutrophil cytosol factor 2 | -2.77 | < 0.001 | -1.59 | 0.006 |

FC: fold change; FDR: false discovery rate; GBS: Guillain-Barré syndrome

### SomaScan recovers established neuropathy biomarkers in acute Guillain-Barré syndrome

We next assessed the abundance of fluid biomarkers previously reported in peripheral neuropathies (Fig. 1E).^9,16,17,19^ Several markers linked to axonal or myelin injury and neuroinflammation were significantly elevated in acute Guillain-Barré syndrome compared with healthy controls, including neurofilament light chain (log□FC = 0.90), transmembrane protease serine 5 (TMPRSS5; log□FC = 0.76), matrix metalloproteinase-9 (MMP-9; log□FC = 0.65), neurofilament heavy chain (log□FC = 0.59), calgranulin B (S100A9; log□FC = 0.47), osteopontin (SPP1; log□FC = 0.42) and glutaredoxin-1 (GLRX; log□FC = 0.39). In contrast, multiple axonal adhesion molecules were significantly reduced, most prominently contactin-6 (CNTN6; log□FC = –0.73), neural cell adhesion molecule 2 (NCAM2; log□FC = –0.73), contactin-1 (CNTN1; log□FC = – 0.64), contactin-2 (CNTN2; log□FC = –0.53) and contactin-4 (CNTN4; log□FC = –0.51). A full list of previously reported biomarkers and their comparison statistics is provided in Supplementary Table 6.

### Immune and sarcomere pathways are increased, whereas axonal adhesion and development pathways are decreased in acute Guillain-Barré syndrome

Gene-set enrichment analysis revealed striking biological differences between the acute and recovery phases of Guillain-Barré syndrome. Acute-phase samples showed strong enrichment of immune-related pathways, including B-cell signalling, interleukin and JAK-STAT pathways, cell-cycle regulation, and protein degradation, suggesting robust systemic immune activation. In contrast, pathways involved in axon development, synapse assembly, and extracellular matrix organisation were negatively enriched, consistent with structural and functional impairment of peripheral nerves (Fig. 2A, Supplementary Table 7).

**Figure 2.**
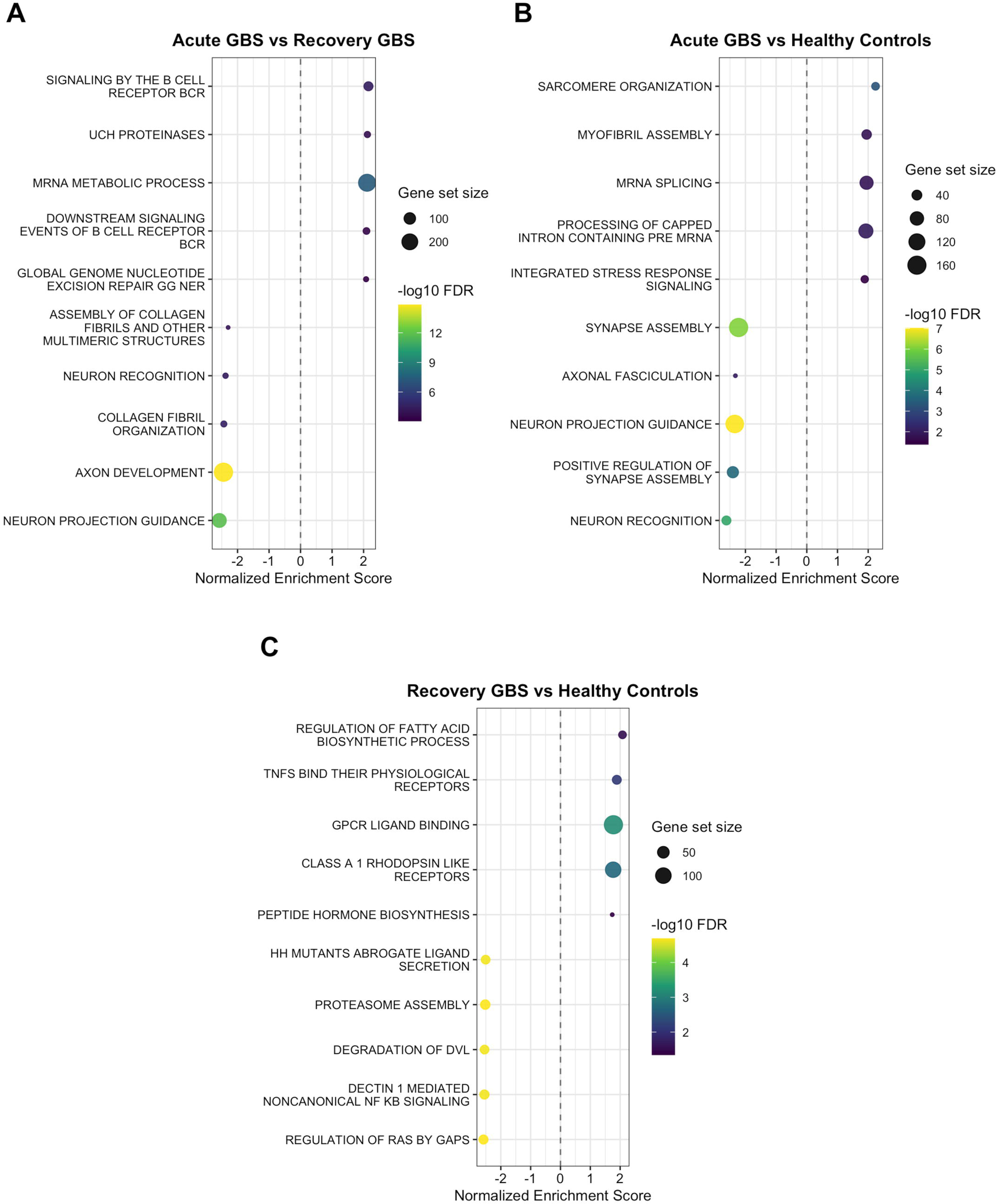
Gene Set Enrichment Analysis (GSEA) across clinical stages of Guillain-Barré syndrome. Dot plots display the top five positively and negatively enriched pathways based on the normalized enrichment score using the *Reactome* and *Gene Ontology: Biological Process* gene sets. Panels represent comparisons between: **(A)** acute Guillain-Barré syndrome compared to one-year follow-up, **(B)** acute Guillain-Barré syndrome compared to healthy controls, and **(C)** Guillain-Barré syndrome at one-year follow-up compared to healthy controls. Only statistically significant results (FDR < 0.05) are shown. FDR, false discovery rate; GBS, Guillain-Barré syndrome. Alt text: Three dot plots labelled A to C displaying Gene Set Enrichment Analysis results. In all plots, biological pathways are listed on the y-axis, with dot size and color gradients representing the normalized enrichment score and statistical significance.

When compared with controls, acute-phase serum additionally showed marked positive enrichment of muscle sarcomere-related proteins, alongside the same negative enrichment patterns in axonal and synaptic pathways (Fig. 2B, Supplementary Table 8).

### Recovery-phase serum shows enrichment of lipid metabolism and TNF-superfamily pathways

Recovery-phase samples demonstrated positive enrichment of pathways involved in fatty acid biosynthesis and tumour necrosis factor (TNF)-superfamily signalling, whereas several inflammatory pathways were negatively enriched compared with healthy controls (Fig. 2C, Supplementary Table 9).

### Unbiased acute proteomic profiling does not identify prognostic signatures

No novel individual proteins measured during the acute phase were significantly associated with long-term prognosis after correcting for multiple testing. Consistent with the lack of differential abundance, elastic net regression failed to identify a robust multivariable proteomic signature predictive of functional outcome. The complete set of results from this analysis is available via the project repository.

### Mendelian randomisation implicates specific circulating proteins in disease susceptibility

Of the 82 evaluated proteins, MR revealed significant causal associations between genetically determined levels of several circulating proteins and the risk of developing Guillain-Barré syndrome. In particular, a higher risk of Guillain-Barré syndrome was associated with predicted lower levels of PENK, CFHR4, CNDP1, CRISPLD2, GSTM4, and SERPING1, as well as genetically predicted higher levels of FCGR2B, MMP8, and SORD. Among these, the causal estimates for CNDP1 (Beta-Ala-His dipeptidase), CRISPLD2 (Cysteine-rich secretory protein LCCL domain-containing 2), and SERPING1 (Plasma protease C1 inhibitor) were particularly robust (Table 4), since they remained statistically significant after correction for multiple testing. Subsequent colocalization analyses did not yield positive results, likely reflecting the limited statistical power of the currently available Guillain-Barré syndrome GWAS summary statistics. The complete MR results are detailed in the Supplementary Material.

**Table 4.** Mendelian randomisation estimates for the causal effects of circulating proteins on Guillain-Barré syndrome susceptibility.

| Exposure | Method<sup>a</sup> | IV | $\beta^b$ | OR (95% CI) | p-value |
| --- | --- | --- | --- | --- | --- |
| CNDP1 | IVW | 62 | -0.505 | 0.60 (0.45 - 0.81) | < 0.001 |
|  | MR-Egger | 62 | -0.850 |  | 0.005 |
|  | Weighted median | 62 | -0.675 |  | 0.004 |
|  | MR-PRESSO | 62 | -0.505 |  | 0.001 |
|  | MR-RAPS | 62 | -0.508 |  | < 0.001 |
| CRISPLD2 | IVW | 48 | -0.750 | 0.47 (0.33 - 0.68) | < 0.001 |
|  | MR-Egger | 48 | -0.762 |  | 0.017 |
|  | Weighted median | 48 | -0.762 |  | 0.006 |
|  | MR-PRESSO | 48 | -0.750 |  | < 0.001 |
|  | MR-RAPS | 48 | -0.758 |  | < 0.001 |
| SERPING1 | IVW | 53 | -0.497 | 0.61 (0.48 - 0.77) | < 0.001 |
|  | MR-Egger | 53 | -0.611 |  | 0.013 |
|  | Weighted median | 53 | -0.466 |  | 0.010 |
|  | MR-PRESSO | 53 | -0.497 |  | < 0.001 |
|  | MR-RAPS | 53 | -0.499 |  | < 0.001 |

CI, confidence interval; IV, instrumental variables; IVW, inverse variance weighted; MR, Mendelian randomisation; OR, odds ratio.
<sup>a</sup> Mendelian Randomisation method.
<sup>b</sup> Estimation of the causal relationship between exposure and outcome.

### Validation of candidate proteins

Serum amyloid A1 (SAA1) showed the most pronounced decrease from the acute to the recovery phase, and it remained significantly elevated relative to controls in the recovery phase. In the validation cohort assessed using the Meso Scale Discovery platform, total SAA levels were significantly higher in acute Guillain-Barré syndrome (median [IQR]: 9408 ng/mL [4315–25169]) than in both recovery samples (4428 ng/mL [3033–6999]; adjusted *P* < 0.001) and controls (2389 ng/mL [965–3576]; adjusted *P* < 0.001). Furthermore, SAA levels during the recovery phase remained significantly elevated compared to healthy controls (adjusted *P* = 0.016, Fig. 3A-B).

**Figure 3.**
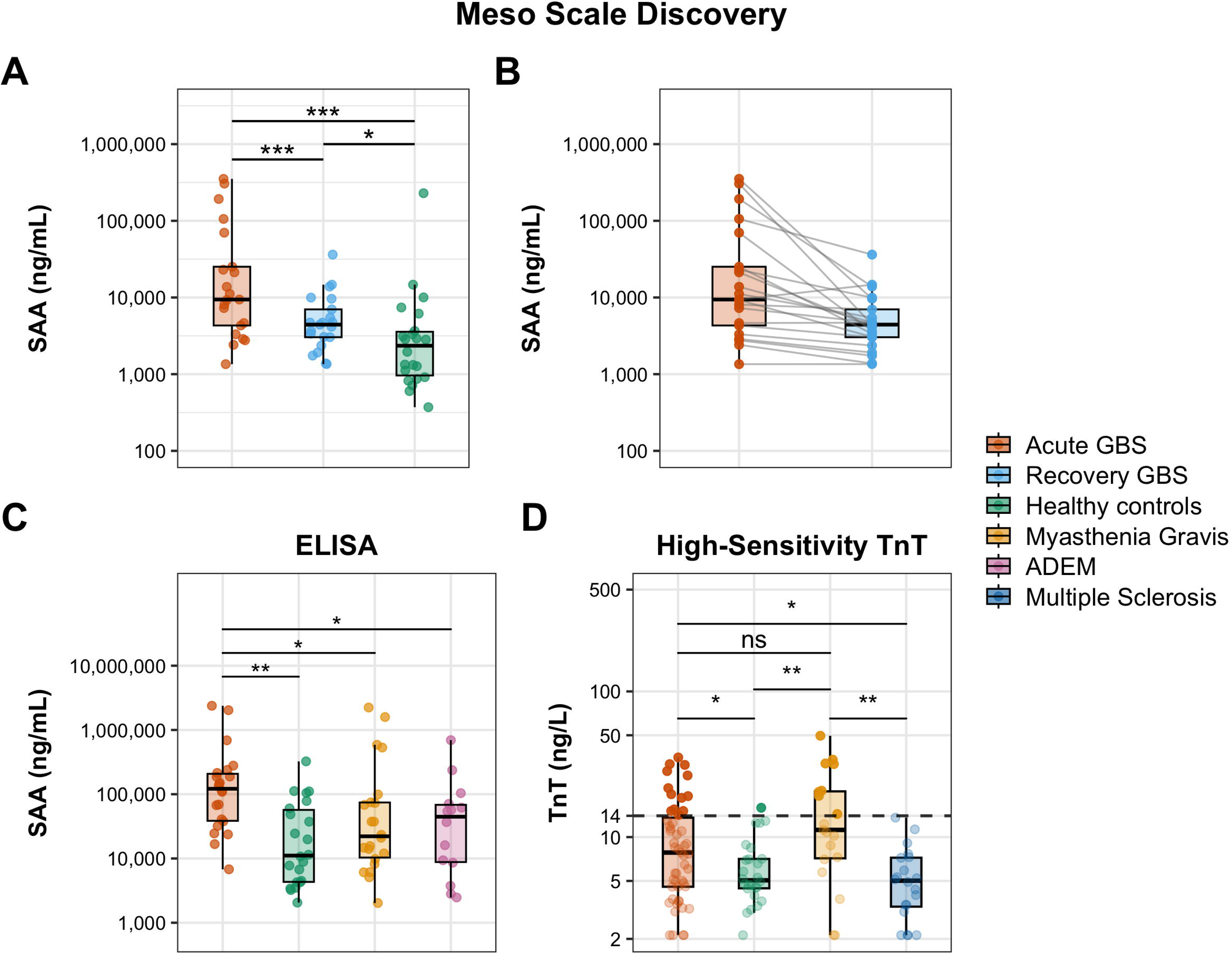
Orthogonal validation of candidate biomarkers. All data is presented on a log_10_ scale. **(A–B)** Meso Scale Discovery analysis of serum amyloid A (SAA) levels. **(A)** Comparative analysis of SAA concentrations in acute GBS (*n* = 21), 1-year recovery GBS (*n* = 21), and healthy controls (*n* = 20). **(B)** Pairwise trajectories illustrating the decline of SAA levels between the acute phase and the 1-year follow-up in individual GBS patients. **(C)** Validation of SAA levels using standard ELISA in acute GBS (*n* = 22), healthy controls (*n* = 22), myasthenia gravis (*n* = 22), and ADEM (*n* = 14). **(D)** High-sensitivity Troponin T levels in acute GBS (*n* = 51), healthy controls (*n* = 26), myasthenia gravis (*n* = 19), and multiple sclerosis (*n* = 20). The horizontal dashed line indicates the 99^th^ percentile upper reference limit (14 ng/L), and the samples above this threshold are highlighted. For all boxplots, the centre horizontal line indicates the median, the box bounds represent the interquartile range, and the whiskers extend to the minimum and maximum values. ∗*P* < 0.05, ∗∗*P* < 0.01, ∗∗∗*P* < 0.001; ns, non-significant. ADEM, acute disseminated encephalomyelitis; GBS, Guillain-Barré syndrome; hs-cTnT, high-sensitivity cardiac troponin T; SAA, serum amyloid A. ALT TEXT: Four boxplots labelled A to D validating candidate biomarkers. A to C show serum amyloid A concentrations, while D shows high-sensitivity cardiac troponin T levels across diagnostic categories.

To assess disease specificity, total SAA levels were quantified by ELISA in a broader cohort that included samples from patients with myasthenia gravis and acute disseminated encephalomyelitis. SAA concentrations were significantly higher in acute Guillain-Barré syndrome (median [IQR]: 121,913 ng/mL [38,557–208,067]) compared with healthy controls (11,102 ng/mL [4,322–57,258]; adjusted *P* = 0.001). Furthermore, this elevation was disease-specific, with levels significantly exceeding those observed in both myasthenia gravis (22,166 ng/mL [10,378–74,365]; adjusted *P* = 0.043) and acute disseminated encephalomyelitis (45,500 ng/mL [8,807–68,499]; adjusted *P* = 0.034; Fig. 3C). Importantly, while absolute concentrations were higher on the ELISA platform compared to the Meso Scale Discovery assay, a subset of samples analysed on both platforms (*n* = 34) demonstrated a strong positive correlation (ρ = 0.96; *P* < 0.001), confirming robust inter-assay agreement in relative quantification (Supplementary Fig. 3).

Two muscle associated proteins, TNNI2 and ACTN2, were elevated in acute Guillain-Barré syndrome in the SomaScan^®^ dataset. Given their relevance to a disease presenting with acute weakness, validation was pursued. However, ELISA could not reliably detect either protein in patient or control serum, and validation was therefore not possible.

Additionally, both skeletal and cTnT isoforms (TNNT3 and TNNT2) showed significant elevation relative to healthy controls in the discovery dataset (log□FC = 0.61 and 0.69, respectively). Although this fold-change fell below our pre-established threshold for differential abundance, the high clinical accessibility of cTnT testing, together with recent reports of elevated cTnT in serum from patients with amyotrophic lateral sclerosis,^13,31,32^ prompted us to pursue validation. Notably, 25.5% (13/51) of the Guillain-Barré syndrome cohort exhibited levels above the predefined 14 ng/L threshold typically used for myocardial injury diagnosis (Fig. 3D).^29^ The frequency of hs-cTnT elevation varied significantly across the diagnostic categories (*P* < 0.001). Specifically, this prevalence was remarkably high in the Guillain-Barré syndrome and myasthenia gravis (42.1%; 8/19) cohorts, significantly exceeding that of healthy controls (3.8%; 1/26) and patients with multiple sclerosis (0%). Furthermore, the frequency of elevation did not differ significantly between the Guillain-Barré syndrome and myasthenia gravis groups. Demographic characteristics of the validation cohorts are presented in Supplementary Tables 10 and 11.

## Discussion

This study provides the first longitudinal, large-scale proteomic characterisation of Guillain–Barré syndrome using an aptamer-based platform. By analysing paired serum samples obtained at disease onset and one year later, and comparing them with healthy controls, we identified distinct stage-specific proteomic signatures that delineate the acute and recovery phases of the disease. Gene-set enrichment analysis revealed coordinated shifts in immune, neurostructural, and metabolic pathways across the disease course. Furthermore, MR provided causal genetic evidence linking specific systemic proteins directly to disease susceptibility. Finally, selected differentially abundant proteins were validated using independent immunoassay platforms, supporting the robustness of our discovery-phase findings.

The acute phase of the disease was characterised by a marked increase of classical acute-phase reactants and inflammatory mediators, including SAA1, ferritin light and heavy chains, and osteopontin. Components of the muscle contractile apparatus, such as ACTN2 and TNNI2, were also elevated, likely reflecting secondary muscle involvement. In parallel, several neurostructural adhesion molecules, including the contactin family and NCAM2, were significantly reduced, consistent with a loss of axonal integrity and paranodal disruption. Although reduced contactin-1 has been described in autoimmune nodopathies, particularly anti-contactin-1 autoimmune nodopathy,^17,18^ decreased circulating contactins have not previously been reported in Guillain-Barré syndrome, reinforcing their potential as serum markers of paranodal damage. The reduction of other extracellular matrix adhesion molecules, such as chondroadherin, further supports structural damage at the peripheral nerve level. Interestingly, although we observed lower NCAM2 levels in acute Guillain-Barré syndrome, elevated soluble NCAM and NCAM1 have been reported in both acquired and hereditary demyelinating neuropathies.^33,34^ These discrepancies may reflect isoform-specific differences or temporal dynamics, where acute depletion corresponds to nerve injury and later increases reflect regenerative responses, although we did not observe such increases at the one-year follow-up.

Gene-set enrichment analysis supported these findings, revealing activation of B-cell, interleukin, JAK–STAT, and sarcomere-related pathways, alongside suppression of axonogenesis, synapse assembly, and extracellular matrix organisation. Together, these data highlight the convergence of intense immune activation and axonal-paranodal injury that characterises acute Guillain-Barré syndrome.

Among the differentially abundant proteins, SAA1 showed the largest fold change. Given this, and the widespread availability of SAA assays in routine clinical practice, we pursued validation using conventional immunoassays. Biologically, SAA1 is an inducible acute-phase reactant predominantly produced by hepatocytes under interleukin-6 regulation, with roles in monocyte recruitment, complement activation, and lipid metabolism.^35^ Experimental evidence also indicates that Schwann cells can produce SAA1 in response to injury, promoting macrophage chemotaxis and myelin clearance.^36^ Thus, elevated circulating SAA1 in Guillain-Barré syndrome may reflect a combination of local Schwann-cell–derived response and systemic inflammation from the disease or the antecedent infection. To differentiate between these possibilities, we used acute disseminated encephalomyelitis patients, a prototypical post-infectious neurological disorder, as a disease control. The higher levels observed in Guillain-Barré syndrome suggest that this increase is primarily driven by the disease itself rather than being a residual effect of the infection. However, although SAA levels were significantly higher in acute samples compared with recovery and controls, considerable overlap limits its diagnostic specificity. Importantly, whereas the SomaScan^®^ platform quantified SAA isoforms separately, the immunoassays used for validation measured total SAA to align with tests already available in clinical practice. Recent reports suggest increased SAA2 and SAA4, but not SAA1, in Guillain-Barré syndrome, pointing to possible isoform-specific regulation.^37^ However, our proteomic data did not show significant changes in SAA2 (log□FC = 0.67, FDR = 0.06) or SAA4 (log□FC = 0.004, FDR = 0.99). This discrepancy highlights a predominant contribution of SAA1 to the acute inflammatory signature in our cohort.

Among biomarker candidates, several sarcomere structural proteins emerged in our proteomic analysis, likely reflecting neuromuscular involvement in acute Guillain-Barré syndrome. In line with this, creatine kinase MM was significantly elevated in the acute phase compared to controls (log□FC = 0.81) and remained abnormally high during the recovery phase (log□FC = 0.83), demonstrating no significant longitudinal change. This sustained elevation observed across multiple sarcomeric proteins is consistent with ongoing muscle denervation. In contrast, other sarcomere structural proteins like ACTN2 and TNNI2 successfully differentiated the acute phase from the recovery phase. However, when attempting orthogonal validation, ACTN2 and TNNI2 fell below the immunoassay detection threshold. This suggests either circulating concentrations below the detection limit of standard ELISAs (as it also happens with sNfL) or potential aptamer cross-reactivity in the discovery platform. Interestingly, a divergent pattern emerged within the troponin family. While the skeletal muscle isoform of troponin I was elevated in acute samples, the cardiac isoform did not differ across groups. Conversely, both skeletal and cTnT isoforms demonstrated significant elevation relative to healthy controls. Given the diagnostic availability of hs-cTnT in clinical practice, we validated this finding using a high-sensitivity ECLIA. Notably, 25.5% of the Guillain-Barré cohort showed hs-cTnT levels above the upper reference limit,^29^ a proportion significantly higher than that of healthy controls and patients with multiple sclerosis (3.8% and 0%, respectively). The source of this elevation warrants careful interpretation. Although acute myocardial injury, such as Takotsubo cardiomyopathy, has been described in Guillain-Barré syndrome,^38^ its low incidence is unlikely to account for the elevations observed in over a quarter of our cohort. Instead, skeletal muscle is increasingly recognised as a non-cardiac source of cTnT, being elevated in inflammatory myopathies and in amyotrophic lateral sclerosis.^13,31,32,39^ This is further supported by our disease control cohort, where patients with myasthenia gravis also displayed a remarkably high frequency of hs-cTnT elevation (42.1%). Together, these data indicate that mild-to-moderate hs-cTnT increases in Guillain-Barré syndrome likely reflect neuromuscular damage rather than occult cardiac ischemia, highlighting a limitation when evaluating suspected cardiovascular complications in these patients.

Our analysis also recapitulated several biomarkers previously associated with peripheral nerve injury, including neurofilament light and heavy chains, S100A9, and osteopontin, supporting both the technical validity and biological relevance of SomaScan^®^ profiling in Guillain-Barré syndrome. Of particular interest, TMPRSS5, a transmembrane serine protease expressed by myelinating Schwann cells, was markedly elevated in acute Guillain-Barré syndrome. TMPRSS5 has been described in Charcot–Marie–Tooth disease type 1A,^22^ a hereditary demyelinating neuropathy, but, to our knowledge, has not previously been linked to inflammatory neuropathies, making its elevation here notable and potentially informative regarding Schwann-cell involvement in acute disease. In contrast, periaxin, another proposed marker of Schwann-cell injury, was not significantly increased in acute samples (log□FC = 0.09, FDR = 0.36), consistent with prior reports indicating that periaxin is detectable in plasma but not consistently in serum.^9^ Other relevant biomarkers, such as peripherin,^8^ were not assessed because they are not included in the SomaScan^®^ panel.

At one-year follow-up, the serum proteome showed enrichment of pathways related to lipid metabolism, protein turnover, and TNF-superfamily signalling. The increased abundance of certain apolipoproteins (APOC1, APOA4, APOF) has previously been associated with Guillain-Barré syndrome,^40^ and, together with elevated levels of proteasomal proteins (PSMA7, USP14), may suggest metabolic reprogramming and remyelination-associated repair processes. The enrichment of TNF-superfamily pathways is noteworthy, as TNF-α gene polymorphisms have been linked to increased susceptibility to acute motor axonal and motor-sensory axonal neuropathies,^41^ and Guillain-Barré syndrome has been reported as a complication of anti-TNF therapy.^42^ Altogether, this evidence further supports a role of the TNF-superfamily signalling in the pathophysiology of Guillain-Barré syndrome.

Our MR analyses provide evidence that specific circulating proteins are not merely reactive biomarkers, but may lie on the causal pathway to Guillain-Barré syndrome susceptibility. By utilizing naturally randomly allocated genetic variants as lifelong proxies for protein levels, MR minimises environmental confounding and reverse causation to infer true directional causality. The biological functions of our most robust causal factors outline a complex genetic vulnerability to Guillain-Barré syndrome. First, genetically predicted low levels of plasma protease C1 inhibitor, a major inhibitor of the classical and lectin complement pathway, are linked to increased disease susceptibility. This causal implication perfectly aligns with established Guillain-Barré syndrome pathogenesis, where data from human pathological studies and animal models suggest that the complement pathway has an important role.^43,44^ Although not as robust, decreased levels of complement factor H-related protein 4, which enhances the activity of the complement inhibitor factor H, are also linked to disease susceptibility, highlighting again the role of the complement cascade in disease pathogenesis. Second, we found that low levels of CRISPLD2 also increase Guillain-Barré syndrome risk. CRISPLD2 is a circulating protein that binds lipopolysaccharide (LPS) and reduces LPS-induced pro-inflammatory cytokine production.^45^ Given that infection with Gram-negative bacteria expressing LPS, such as *Campylobacter jejuni*, is an important trigger for Guillain-Barré syndrome, the reduced levels of CRISPLD2 may result in a more extensive immune-response to the antecedent infection, facilitating the cross-reactivity that drives the disease. Finally, the implication of CNDP1, an oxidative stress regulator, suggests that a baseline deficit in metabolic resilience may affect nerve vulnerability. In line with this, lower levels of GSTM4, a glutathione transferase, also show association with disease susceptibility, although less robust. Together, these findings suggest that the progression from a common antecedent infection to a severe immune-mediated neuropathy is marked by a specific intrinsic host baseline characterised by disrupted complement regulation, impaired pathogen buffering, and oxidative defence.

The main limitation of this study is its modest sample size, determined by the exploratory nature of the study, the cost of the technology and the scarcity of samples. This sample size may have reduced our statistical power to detect subtle protein changes and limited our ability to interrogate clinical and biological heterogeneity within Guillain-Barré syndrome. By analysing demyelinating and axonal variants, different preceding infections, and varying ganglioside seropositivity as a single cohort, we may have obscured phenotype-specific proteomic signatures that larger, stratified studies could resolve. Furthermore, while healthy controls provide a necessary baseline, they are imperfect controls for a post-infectious disorder, as some observed changes may still reflect the antecedent infection rather than the neuropathy itself. Importantly, collecting the samples at hospital admission may affect the results, as some biomarkers may have delayed kinetics. As an example, sNfL typically peaks approximately two weeks after symptom onset.^4^ Additionally, the SomaScan^®^ platform does not include some proteins of interest in peripheral nerve biology, and serum measurements may not fully capture processes occurring within the peripheral nerve microenvironment. Validation efforts were necessarily limited to a targeted subset of proteins, and additional orthogonal confirmation will be required to clarify their robustness and clinical applicability. Moreover, because our clinical cohort and the genomic datasets utilised for MR were predominantly of European ancestry, subsequent studies in diverse populations are necessary to confirm the generalisability of these findings. Finally, although the longitudinal design provides valuable temporal resolution, mechanistic studies will be needed to determine whether the identified proteins play a causal role in disease pathogenesis or predominantly reflect downstream consequences of immune-mediated nerve injury.

Despite these limitations, the study has several strengths, including a well-phenotyped multicentre cohort and the use of a high-throughput aptamer-based platform complemented by independent immunoassay validation. A critical asset of our design was the use of paired samples collected at clearly defined disease stages. By allowing each patient to serve as their own control, this approach minimises the confounding effect of inter-individual proteomic variability, and ensures that the observed changes reflect disease-driven biological processes rather than background noise. Together, these elements provide a robust framework for identifying biologically meaningful proteomic signatures in Guillain–Barré syndrome. In addition, the dataset emerging from this study will certainly be useful to inform the design and interpretation of future studies using high-throughput technologies and as a reference dataset to validate other biomarkers described in the future.

In conclusion, aptamer-based serum proteomics reveals distinct molecular fingerprints of the acute and recovery phases of Guillain–Barré syndrome. Larger, deeply phenotyped longitudinal cohorts integrating proteomic, genomic, metagenomic, metabolomic, and detailed clinical data will be essential to delineate mechanistic pathways and to support the translation of promising protein biomarkers into clinical practice.

## Supporting information

Mendelian Randomisation Full Methods and Results

STROBE checklist

## Data availability

The raw anonymised SomaScan® proteomic datasets, the complete differential abundance and prognosis results, and the custom R scripts used for all statistical analyses and data visualisation have been deposited in a public repository and are openly available on GitHub at https://github.com/NeRveLabBCN/GBS-Proteomics. Anonymised data not published within this article will be made available by request from any qualified investigator.

## Acknowledgements

The authors thank the Department of Medicine, Universitat Autònoma de Barcelona, Spain for their support in the development of this work. The authors also thank all our patients for their support and collaboration. Several authors of this publication are members of the European Reference Network for rare neuromuscular diseases (EURO-NMD). The authors utilised a Large Language Model (Google Gemini) to assist with copyediting and refining the language of the manuscript; the authors reviewed, edited, and take full responsibility for all final published content.

## Supplementary material

Supplementary material is available alongside the online version of this preprint.

### Appendix

SPAiN Consortium collaborators:

Ana Baltasar Corral (Hospital Universitario Ramón y Cajal, Madrid), Rodrigo Álvarez Velasco (Hospital Universitario Ramón y Cajal, Madrid), Ana Pinel González (Hospital Universitario de Getafe, Madrid), María Sánchez Puche (Hospital Universitario de Getafe, Madrid), Antonio Miguel Caliz Rodríguez (Hospital Universitario de Getafe, Madrid), María Teresa Cachero García (Hospital Universitario de Getafe, Madrid), Amaia Jauregui Barrutia (Hospital Universitario Cruces), Amaia Martínez Arroyo (Hospital Galdakao-Usansolo), Alejandra Collia Fernández (Hospital Universitario Araba), Álvaro Laviana Marín (Hospital Universitario Virgen Macarena, Sevilla), Eva María Martínez Fernández (Hospital Universitario Virgen Macarena, Sevilla), Yolanda Morgado Linares (Hospital Universitario Virgen Macarena, Sevilla), Arnau Llauradó (Hospital Universitari Vall d’Hebron, Barcelona), Lidia Girame (Hospital Universitari Vall d’Hebron, Barcelona), Beatriz Catón Sanz (Hospital Universitario de Burgos), Cecilia Gil Polo (Hospital Universitario de Burgos), Mónica Bártulos Iglesias (Hospital Universitario de Burgos), Neus Guiu González (Hospital Universitario de Burgos), Sandra Jorge Roldán (Hospital Universitario de Burgos), Begoña Zapata Macías (Hospital Universitario Puerta del Mar, Cádiz), Carla Marco Cazcarra (Hospital Universitario de Bellvitge, Barcelona), Roser Velasco Fargas (ICO Bellvige, Barcelona), María Antonia Albertí Aguiló (Hospital Universitario de Bellvitge, Barcelona), Valentina Vélez Santamaria (Hospital Universitario de Bellvitge, Barcelona), Velina Nedkova Hristova (Hospital Universitario de Bellvitge, Barcelona), Moisés Morales de la Prida (Hospital Universitario de Bellvitge, Barcelona), Carlos Pablo de Fuenmayor Fernández de la Hoz (Hospital Universitario 12 de Octubre, Madrid), Juan Francisco Gonzalo Martínez (Hospital Universitario 12 de Octubre, Madrid), María Paz Guerrero (Hospital Universitario 12 de Octubre, Madrid), Paloma Martín Jiménez (Hospital Universitario 12 de Octubre, Madrid), Carmen Valderrama Martín (Hospital Universitario Virgen de las Nieves, Granada), Daniel Santirso Rodríguez (Hospital Universitario Central de Asturias, Oviedo), Germán Morís de la Tassa (Hospital Universitario Central de Asturias, Oviedo), Elva Murillo (Hospital Universitario Virgen de Valme, Sevilla), Eva María Martínez Acebes (Hospital Universitario Infanta Leonor, Madrid), Raquel Cuenca Hernández (Hospital Universitario Infanta Leonor, Madrid), Francisco Javier Cabello Murgui (Hospital La Fe, Valencia), Inmaculada Pitarch Castellano (Hospital La Fe, Valencia), Jesús Jiménez Jiménez (Hospital La Fe, Valencia), Pilar Martí Martínez (Hospital La Fe, Valencia), Teresa Sevilla Mantecón (Hospital La Fe, Valencia), Francisco Javier Rodríguez de Rivera Garrido (Hospital Universitario La Paz, Madrid), Laura Lacruz Ballester (Hospital Universitario La Paz, Madrid), Giuseppe Lucente (Hospital Universitario Germans Trias i Pujol, Barcelona), Guillermina García Martín (Hospital Regional Universitario de Málaga), Íñigo Rojas-Marcos (Hospital Universitario Virgen del Rocío, Sevilla), Javier Almeida Velasco (Hospital de Urduliz), Patricia Rodrigo Armenteros (Hospital Universitario Basurto, Bilbao), Solange Kapetanovic García (Hospital Universitario Basurto, Bilbao), Roberto Fernández Torrón (Hospital Universitario Donostia), Marta Rodríguez Camacho (Hospital Universitario Torrecárdenas, Almería), Beatriz Vélez Gómez (Hospital Universitario Torrecárdenas, Almería), María del Mar Martínez Salmeron (Hospital Universitario Torrecárdenas, Almería), Beatrice Canneti Heredia (Hospital Clínico de Santiago de Compostela), Tania García Sobrino (Hospital Clínico de Santiago de Compostela), Jose María Cabrera-Maqueda (Hospital Clínic de Barcelona), Lidia Sabater (Hospital Clínic de Barcelona), Raquel Ruiz-Garcia (Hospital Clínic de Barcelona), Eugenia Martínez Hernández (Hospital Clínic de Barcelona).

## References

1. Leonhard SE, Papri N, Querol L, Rinaldi S, Shahrizaila N, Jacobs BC. Guillain–Barré syndrome. Nat Rev Dis Primer. 2024;10(1). doi:10.1038/s41572-024-00580-4

2. Willison HJ, Jacobs BC, Van Doorn PA. Guillain-Barré syndrome. The Lancet. 2016;388(10045):717–727. doi:10.1016/S0140-6736(16)00339-1

3. Martín-Aguilar L, Camps-Renom P, Lleixà C, et al. Serum neurofilament light chain predicts long-term prognosis in Guillain-Barré syndrome patients. J Neurol Neurosurg Psychiatry. 2021;92(1):70–77. doi:10.1136/jnnp-2020-323899

4. Van Tilburg SJ, Teunissen CE, Maas CCHM, et al. Dynamics and prognostic value of serum neurofilament light chain in Guillain-Barré syndrome. eBioMedicine. 2024;102:105072. doi:10.1016/j.ebiom.2024.105072

5. Thomma RCM, Luijten LWG, Van Tilburg SJ, et al. Neurofilament light chain improves clinical prognostic models for Guillain-Barré syndrome. J Neurol Neurosurg Psychiatry. Published online May 2, 2025:jnnp-2025-336046. doi:10.1136/jnnp-2025-336046

6. Gaetani L, Blennow K, Calabresi P, Di Filippo M, Parnetti L, Zetterberg H. Neurofilament light chain as a biomarker in neurological disorders. J Neurol Neurosurg Psychiatry. 2019;90(8):870–881. doi:10.1136/jnnp-2018-320106

7. Khalil M, Teunissen CE, Lehmann S, et al. Neurofilaments as biomarkers in neurological disorders — towards clinical application. Nat Rev Neurol. 2024;20(5):269–287. doi:10.1038/s41582-024-00955-x

8. Keddie S, Smyth D, Keh RYS, et al. Peripherin is a biomarker of axonal damage in peripheral nervous system disease. Brain. 2023;146(11):4562–4573. doi:10.1093/brain/awad234

9. Bellanti R, Keh RYS, Keddie S, et al. Plasma periaxin is a biomarker of peripheral nerve 2 demyelination.

10. Van Doorn PA, Van den Bergh PYK, Hadden RDM, et al. European Academy of Neurology/Peripheral Nerve Society Guideline on diagnosis and treatment of GUILLAIN–BARRÉ syndrome. Eur J Neurol. 2023;30(12):3646–3674. doi:10.1111/ene.16073

11. Cervia-Hasler C, Brüningk SC, Hoch T, et al. Persistent complement dysregulation with signs of thromboinflammation in active Long Covid. Science. 2024;383(6680). doi:10.1126/science.adg7942

12. Belbasis L, Morris S, Van Duijn C, Bennett D, Walters R. Mendelian randomization identifies proteins involved in neurodegenerative diseases. Brain. 2025;148(7):2412–2428. doi:10.1093/brain/awaf018

13. Dergai O, Wuu J, Koziczak-Holbro M, et al. Skeletal Muscle Biomarkers of Amyotrophic Lateral Sclerosis: A Large□Scale, Multi□Cohort Proteomic Study. Ann Neurol. 2026;99(2):393–407. doi:10.1002/ana.78046

14. Gold L, Ayers D, Bertino J, et al. Aptamer-Based Multiplexed Proteomic Technology for Biomarker Discovery. PLoS ONE. 2010;5(12):e15004. doi:10.1371/journal.pone.0015004

15. Ritchie ME, Phipson B, Wu D, et al. limma powers differential expression analyses for RNA-sequencing and microarray studies. Nucleic Acids Res. 2015;43(7):e47–e47. doi:10.1093/nar/gkv007

16. Wieske L, Smyth D, Lunn MP, Eftimov F, Teunissen CE. Fluid Biomarkers for Monitoring Structural Changes in Polyneuropathies: Their Use in Clinical Practice and Trials. Neurotherapeutics. 2021;18(4):2351–2367. doi:10.1007/s13311-021-01136-0

17. Wieske L, Martín-Aguilar L, Fehmi J, et al. Serum Contactin-1 in CIDP: A Cross-Sectional Study. Neurol Neuroimmunol Neuroinflammation. 2021;8(5):e1040. doi:10.1212/NXI.0000000000001040

18. Caballero-Ávila M, Martín-Aguilar L, Pascual-Goñi E, et al. Long□Term Follow Up in Anti□Contactin□1 Autoimmune Nodopathy. Ann Neurol. 2025;97(3):529–541. doi:10.1002/ana.27142

19. Wieske L, Michael MR, In ‘T Veld SGJG, et al. Proximity extension assay-based discovery of biomarkers for disease activity in chronic inflammatory demyelinating polyneuropathy. J Neurol Neurosurg Psychiatry. 2024;95(7):595–604. doi:10.1136/jnnp-2023-332398

20. Kieseier BC, Clements JM, Pischel HB, et al. Matrix metalloproteinases MMP□9 and MMP□7 are expressed in experimental autoimmune neuritis and the guillain□barré syndrome. Ann Neurol. 1998;43(4):427–434. doi:10.1002/ana.410430404

21. Martín-Aguilar L, Camps-Renom P, Lleixà C, et al. Serum neurofilament light chain predicts long-term prognosis in Guillain-Barré syndrome patients. J Neurol Neurosurg Psychiatry. 2021;92(1):70–77. doi:10.1136/jnnp-2020-323899

22. Wang H, Davison M, Wang K, et al. Transmembrane protease serine 5: a novel Schwann cell plasma marker for CMT1A. Ann Clin Transl Neurol. 2020;7(1):69–82. doi:10.1002/acn3.50965

23. Korotkevich G, Sukhov V, Budin N, Shpak B, Artyomov MN, Sergushichev A. Fast gene set enrichment analysis. Cold Spring Harbor Laboratory. Preprint posted online June 20, 2016. doi:10.1101/060012

24. Gillespie M, Jassal B, Stephan R, et al. The reactome pathway knowledgebase 2022. Nucleic Acids Res. 2022;50(D1):D687–D692. doi:10.1093/nar/gkab1028

25. Ashburner M, Ball CA, Blake JA, et al. Gene Ontology: tool for the unification of biology. Nat Genet. 2000;25(1):25–29. doi:10.1038/75556

26. The Gene Ontology Consortium, Aleksander SA, Balhoff J, et al. The Gene Ontology knowledgebase in 2023. Baryshnikova A, ed. GENETICS. 2023;224(1):iyad031. doi:10.1093/genetics/iyad031

27. Liberzon A, Birger C, Thorvaldsdóttir H, Ghandi M, Mesirov JP, Tamayo P. The Molecular Signatures Database Hallmark Gene Set Collection. Cell Syst. 2015;1(6):417–425. doi:10.1016/j.cels.2015.12.004

28. Merico D, Isserlin R, Stueker O, Emili A, Bader GD. Enrichment Map: A Network-Based Method for Gene-Set Enrichment Visualization and Interpretation. Ravasi T, ed. PLoS ONE. 2010;5(11):e13984. doi:10.1371/journal.pone.0013984

29. Thygesen K, Alpert JS, Jaffe AS, et al. Fourth universal definition of myocardial infarction (2018). Eur Heart J. 2019;40(3):237–269. doi:10.1093/eurheartj/ehy462

30. Von Elm E, Altman DG, Egger M, Pocock SJ, Gøtzsche PC, Vandenbroucke JP. The Strengthening the Reporting of Observational Studies in Epidemiology (STROBE) statement: guidelines for reporting observational studies. J Clin Epidemiol. 2008;61(4):344–349. doi:10.1016/j.jclinepi.2007.11.008

31. Castro-Gomez S, Radermacher B, Tacik P, Mirandola SR, Heneka MT, Weydt P. Teaching an old dog new tricks: serum troponin T as a biomarker in amyotrophic lateral sclerosis. Brain Commun. 2021;3(4):fcab274. doi:10.1093/braincomms/fcab274

32. Lindenborn P, Fabian R, Grehl T, et al. Combination of Serum Neurofilament Light Chain and Serum Cardiac Troponin T as Biomarkers Improves Diagnostic Accuracy in Amyotrophic Lateral Sclerosis. Ann Neurol. Published online October 24, 2025:ana.78066. doi:10.1002/ana.78066

33. Jennings MJ, Kagiava A, Vendredy L, et al. NCAM1 and GDF15 are biomarkers of Charcot-Marie-Tooth disease in patients and mice. Brain. 2022;145(11):3999–4015. doi:10.1093/brain/awac055

34. Niezgoda A, Michalak S, Losy J, Kalinowska-Łyszczarz A, Kozubski W. sNCAM as a specific marker of peripheral demyelination. Immunol Lett. 2017;185:93–97. doi:10.1016/j.imlet.2017.03.011

35. Sack GH. Serum amyloid A – a review. Mol Med. 2018;24(1):46. doi:10.1186/s10020-018-0047-0

36. Jang SY, Shin YK, Lee HY, et al. Local production of serum amyloid a is implicated in the induction of macrophage chemoattractants in Schwann cells during wallerian degeneration of peripheral nerves. Glia. 2012;60(10):1619–1628. doi:10.1002/glia.22382

37. Yao X, Qiao B, Shan F, et al. Elevated Serum Amyloid A2 and A4 in Patients With Guillain–Barré Syndrome. J Clin Neurol. 2025;21(3):213. doi:10.3988/jcn.2024.0469

38. Terayama A, Kuwahara M, Yoshikawa K, et al. Takotsubo cardiomyopathy in Guillain–Barré syndrome. J Neurol. 2024;271(7):4067–4074. doi:10.1007/s00415-024-12295-3

39. Du Fay De Lavallaz J, Prepoudis A, Wendebourg MJ, et al. Skeletal Muscle Disorders: A Noncardiac Source of Cardiac Troponin T. Circulation. 2022;145(24):1764–1779. doi:10.1161/CIRCULATIONAHA.121.058489

40. Jin T, Hu L-S., Chang M, Wu J, Winblad B, Zhu J. Proteomic identification of potential protein markers in cerebrospinal fluid of GBS patients. Eur J Neurol. 2007;14(5):563–568. doi:10.1111/j.1468-1331.2007.01761.x

41. Liu J, Lian Z, Chen H, et al. Associations between tumor necrosis factor-α gene polymorphisms and the risk of Guillain-Barré syndrome and its subtypes: A systematic review and meta-analysis. J Neuroimmunol. 2017;313:25–33. doi:10.1016/j.jneuroim.2017.10.003

42. Patwala K, Crump N, De Cruz P. Guillain-Barré syndrome in association with antitumour necrosis factor therapy: a case of mistaken identity. BMJ Case Rep. 2017;2017:bcr-2017-219481. doi:10.1136/bcr-2017-219481

43. Wanschitz J. Distinct time pattern of complement activation and cytotoxic T cell response in Guillain-Barre syndrome. Brain. 2003;126(9):2034–2042. doi:10.1093/brain/awg207

44. Halstead SK, Zitman FMP, Humphreys PD, et al. Eculizumab prevents anti-ganglioside antibody-mediated neuropathy in a murine model. Brain. 2008;131(5):1197–1208. doi:10.1093/brain/awm316

45. Wang ZQ, Xing WM, Fan HH, et al. The Novel Lipopolysaccharide-Binding Protein CRISPLD2 Is a Critical Serum Protein to Regulate Endotoxin Function. J Immunol. 2009;183(10):6646–6656. doi:10.4049/jimmunol.0802348

