## Supplemental methods, Figures 1-3, Tables 1-11 for "Longitudinal proteomics defines stage-specific molecular signatures in Guillain-Barré syndrome"

### SUPPLEMENTARY MATERIAL

**Contents** ........................................................................................................................................... **Page**

**Supplementary Methods**

#### SomaScan® proteomics technical consistency and quality controls

All samples included were processed in a single analytical batch. To evaluate technical consistency and detect potential outliers, we inspected SomaLogic-provided hybridization and normalization scale factors, performed principal component analysis (PCA) on $\log_2$-transformed relative fluorescence units (Supplementary Fig. 1), and generated sample-to-sample correlation heatmaps using Pearson's correlation (Supplementary Fig. 2). All samples passed the vendor quality metric thresholds. Unsupervised PCA revealed that the first two components captured 22.7% and 12.1% of the total variance for PC1 and PC2, respectively. To formalize outlier detection, 95% confidence intervals (CI) ellipses were constructed for each diagnostic category. PCA identified four samples that fell outside the 95% CI for their category. In the sample-sample correlation heatmaps half of the healthy controls clustered together, although there were six healthy controls mixed with the cases, which could be explained due to variability within the healthy population. In general, we saw that patients with Guillain-Barré syndrome, irrespective of chronology, were mixed, which suggests that the global proteomic profile is dominated by patient vs control differences more than between Guillain-Barré syndrome recovery and acute. One healthy control sample consistently appeared as an outlier (low inter-sample correlation and outside the 95% CI in PCA). Because all samples were processed in the same batch, and passed internal SomaLogic quality metrics, all samples were retained as representations of true biological variance. To ensure statistical robustness, all analyses were repeated with and without this sample as a sensitivity check. Results were unchanged, and the full dataset is reported unless otherwise specified.

#### Supplementary Figures

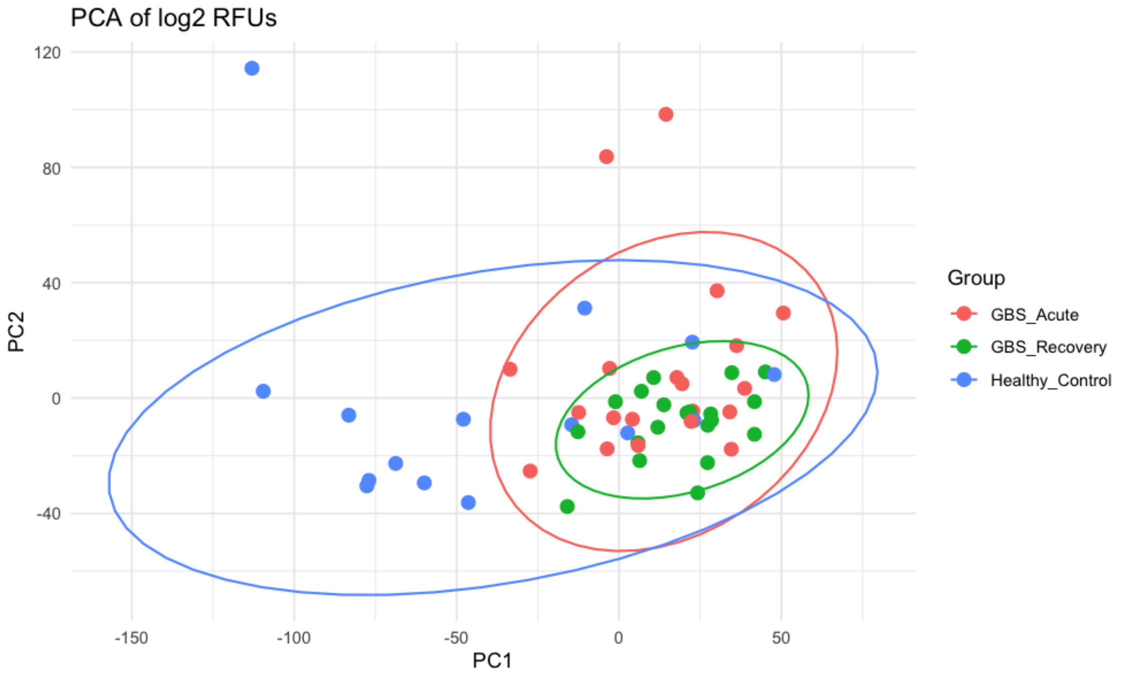

**Supplementary Fig. 1. Principal Component Analysis (PCA) of global proteomic profiles.** Unsupervised PCA was performed on $log_2$-transformed relative fluorescence units. The ellipses represent 95% confidence intervals for each diagnostic category. Four samples fell at the periphery of their respective group boundaries (two acute Guillain-Barré syndrome, one recovery Guillain-Barré syndrome, and one healthy control.

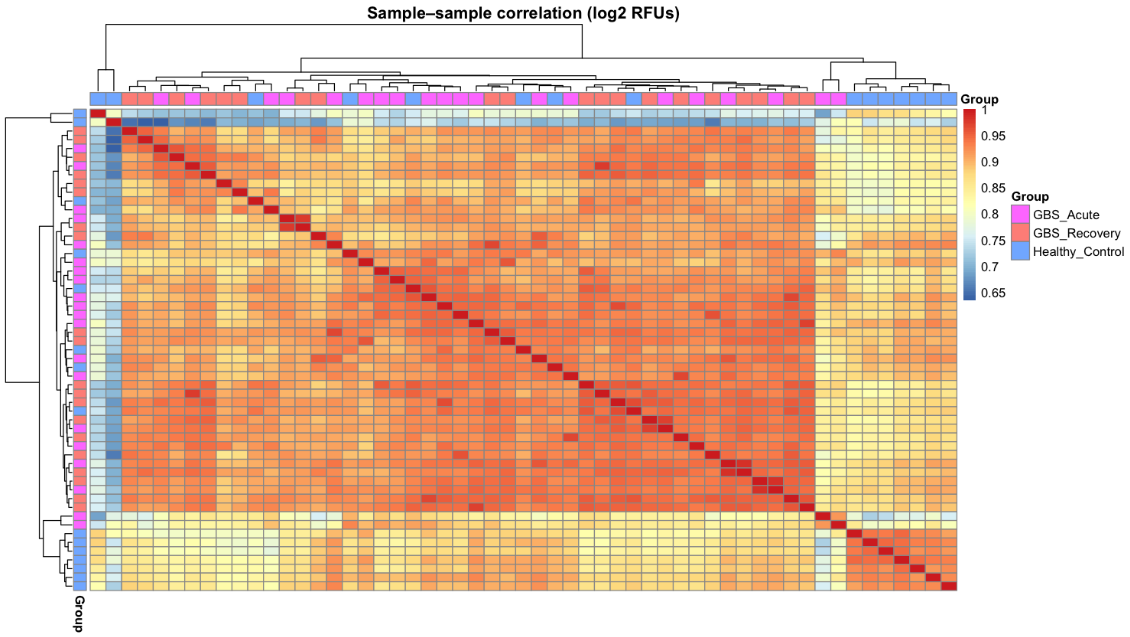

**Supplementary Fig. 2. Sample-to-sample correlation heatmap split by diagnostic groups.** Pearson correlation coefficients between global proteomic profiles are visualized. Warm colors (red) denote high inter-sample correlation, whereas cold colors (blue) denote low correlation.

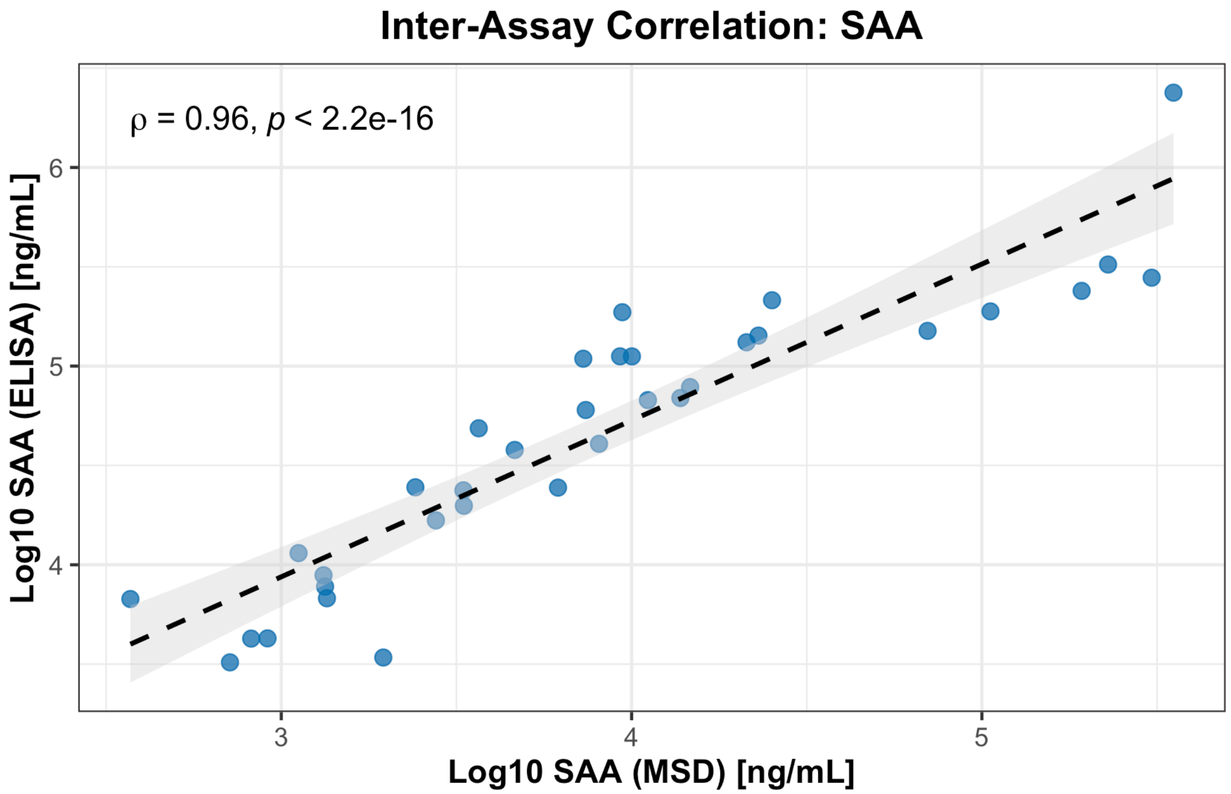

**Supplementary Fig. 3. Inter-assay correlation of serum amyloid A (SAA) quantification.** Scatter plot depicting the correlation of SAA concentrations measured in a subset of matched serum samples (n = 34) using an ELISA versus the Meso Scale Discovery platform. A strong positive correlation is observed (Spearman's $\rho = 0.96$, $P < 0.001$), demonstrating robust inter-assay agreement in relative protein quantification.

### Supplementary Tables

**Supplementary Table 1. Demographic and clinical characteristics of the exploratory cohort.**

| Baseline characteristics | Guillain-Barré syndrome | Healthy controls |
|---|---|---|
| n | 20 | 15 |
| Age, mean ± SD | 56.8 ± 17.1 | 46.1 ± 25.4 |
| Sex, n (%) | | |
| Female | 10 (50) | 7 (46.7) |
| Male | 10 (50) | 8 (53.3) |
| Infectious antecedent, n (%) | | NA |
| Diarrhoea | 5 (25) | |
| Respiratory infection | 11 (55) | |
| None | 4 (20) | |
| Time from onset (days), median (IQR) | 3 (2-4) | NA |
| Initial GDS, median (IQR) | 4 (3-4) | NA |
| GDS at nadir, median (IQR) | 4 (4-4) | NA |
| GBS variant, n (%) | | NA |
| Sensorimotor | 14 (70) | |
| Pure motor | 6 (30) | |
| EMG classification, n (%) | | NA |
| AIDP | 14 (70) | |
| AMAN | 2 (10) | |
| AMSAN | 1 (5) | |
| Equivocal | 1 (5) | |
| Normal | 1 (5) | |
| Missing | 1 (5) | |
| Ventilation, n (%) | 2 (10) | NA |
| Antiganglioside antibodies, n (%) | | |
| GM1 IgG | 5 (25) | NA |
| GM1 IgM | 6 (30) | NA |
| GD1a IgG | 1 (5) | NA |

| | | |
|---|---|---|
| GQ1b IgG | 3 (15) | NA |

AIDP: acute inflammatory demyelinating polyradiculoneuropathy; AMAN: acute motor axonal neuropathy; AMSAN: acute motor-sensory axonal neuropathy; EMG: electromyography; GBS: Guillain-Barré Syndrome; GDS: Guillain-Barré Syndrome Disability Scale; IQR: interquartile range; NA: not applicable; SD: standard deviation,

**Supplementary Table 2. Differentially abundant proteins (*n* = 39) between the acute and recovery phases of Guillain-Barré syndrome.** $Log_2$ fold-change ($Log_2FC$) indicates the magnitude of relative protein abundance. The moderated t-statistic is calculated using empirical Bayes estimation, and reflects the difference standardized by a variance estimate. For both metrics, positive values denote higher abundance in the acute phase, while negative values denote higher abundance in the recovery phase. *P*-values were adjusted for multiple testing using the Benjamini-Hochberg false discovery rate (FDR) method. Proteins are listed in alphabetical order by gene symbol.

| Entrez Gene Symbol | Target Full Name | $Log_2FC$ | t | *P*-Value | FDR |
|---|---|---|---|---|---|
| ADH1C | Alcohol dehydrogenase 1C | -1.037 | -5.151 | <0.001 | <0.001 |
| AGR3 | Anterior gradient protein 3 | -1.074 | -7.256 | <0.001 | <0.001 |
| ANTXR2 | Anthrax toxin receptor 2 | -1.181 | -8.895 | <0.001 | <0.001 |
| ANXA11 | Annexin A11 | 1.120 | 4.084 | <0.001 | 0.003 |
| ANXA7 | Annexin A7 | 1.282 | 4.808 | <0.001 | 0.001 |
| APOA4 | Apolipoprotein A-IV | -2.021 | -8.178 | <0.001 | <0.001 |
| APOC1 | Apolipoprotein C-I | -1.884 | -6.058 | <0.001 | <0.001 |
| APOF | Apolipoprotein F | -1.804 | -4.739 | <0.001 | 0.001 |
| CHAD | Chondroadherin | -1.055 | -5.480 | <0.001 | <0.001 |
| CHGA | Chromogranin-A | -1.127 | -3.523 | 0.001 | 0.011 |
| CLINT1 | Clathrin interactor 1 | 1.147 | 2.951 | 0.005 | 0.033 |
| COL11A2 | Collagen alpha-2(XI) chain | -1.304 | -7.266 | <0.001 | <0.001 |
| CRABP2 | Cellular retinoic acid-binding protein 2 | -1.407 | -11.117 | <0.001 | <0.001 |
| CRIP1 | Cysteine-rich protein 1 | -1.191 | -5.376 | <0.001 | <0.001 |
| EHD3 | EH domain-containing protein 3 | 1.086 | 3.194 | 0.003 | 0.021 |
| FTH1\|FTL | Ferritin | 1.098 | 4.505 | <0.001 | 0.001 |
| FTL | Ferritin light chain | 1.108 | 4.571 | <0.001 | 0.001 |
| H3C1 | Histone H3.1 | 1.180 | 5.132 | <0.001 | <0.001 |
| HMGCS1 | Hydroxymethylglutaryl-CoA synthase; cytoplasmic | -1.047 | -4.661 | <0.001 | 0.001 |
| LDHA | L-lactate dehydrogenase A chain | -1.333 | -4.897 | <0.001 | 0.001 |
| MANF | Mesencephalic astrocyte-derived neurotrophic factor | 1.138 | 3.761 | 0.001 | 0.007 |
| NADK | NAD kinase | 1.332 | 5.603 | <0.001 | <0.001 |
| NCF2 | Neutrophil cytosol factor 2 | 1.589 | 3.815 | <0.001 | 0.006 |
| PA2G4 | Proliferation-associated protein 2G4 | -1.375 | -5.196 | <0.001 | <0.001 |
| PENK | Proenkephalin-A | -1.147 | -3.978 | <0.001 | 0.004 |
| PGD | 6-phosphogluconate dehydrogenase; decarboxylating | -1.520 | -4.975 | <0.001 | 0.000 |
| PRCP | Lysosomal Pro-X carboxypeptidase | -1.083 | -4.583 | <0.001 | 0.001 |
| PSMA7 | Proteasome subunit alpha type-7 | 1.253 | 4.715 | <0.001 | 0.001 |
| SAA1 | Serum amyloid A-1 protein | 1.459 | 5.608 | <0.001 | 0.000 |
| SERPINA5 | Plasma serine protease inhibitor | -1.564 | -3.055 | 0.004 | 0.027 |
| SFRP4 | Secreted frizzled-related protein 4 | -1.202 | -7.857 | <0.001 | <0.001 |
| SH3BGRL2 | SH3 domain-binding glutamic acid-rich-like protein 2 | 1.165 | 3.052 | 0.004 | 0.027 |
| SUMO2 | Small ubiquitin-related modifier 2 | 1.030 | 3.891 | <0.001 | 0.005 |
| TAGLN2 | Transgelin-2 | 1.085 | 3.893 | <0.001 | 0.005 |

| | | | | | |
|---|---|---|---|---|---|
| THBS3 | Thrombospondin-3 | -1.122 | -9.047 | <0.001 | <0.001 |
| THBS4 | Thrombospondin-4 | -1.425 | -9.936 | <0.001 | <0.001 |
| TPM4 | Tropomyosin alpha-4 chain | 1.132 | 3.007 | 0.005 | 0.030 |
| UBASH3B | Ubiquitin-associated and SH3 domain-containing protein B | -1.032 | -3.966 | <0.001 | 0.004 |
| USP14 | Ubiquitin carboxyl-terminal hydrolase 14 | -1.325 | -4.880 | <0.001 | 0.001 |

**Supplementary Table 3. Differentially abundant proteins (*n* = 248) between acute Guillain-Barré syndrome and healthy controls.** Log$_2$ fold-change (Log$_2$FC) indicates the magnitude of relative protein abundance. The moderated t-statistic is calculated using empirical Bayes estimation, and reflects the difference standardized by a variance estimate. For both metrics, positive values denote higher abundance in the acute phase of Guillain-Barré syndrome, while negative values denote higher abundance in healthy controls. *P*-values were adjusted for multiple testing using the Benjamini-Hochberg false discovery rate (FDR) method. Proteins are listed in alphabetical order by gene symbol.

| Entrez Gene Symbol | Target Full Name | Log$_2$FC | t | *P*-Value | FDR |
|---|---|---|---|---|---|
| ACADSB | Short/branched chain specific acyl-CoA dehydrogenase; mitochondrial | -1.085 | -3.093 | 0.004 | 0.019 |
| ACBD6 | Acyl-CoA-binding domain-containing protein 6 | 1.234 | 3.619 | 0.001 | 0.007 |
| ACP5 | Tartrate-resistant acid phosphatase type 5 | 1.396 | 6.715 | <0.001 | <0.001 |
| ACTN2 | Alpha-actinin-2 | 1.897 | 7.001 | <0.001 | <0.001 |
| ACY1 | Aminoacylase-1 | 1.968 | 4.993 | <0.001 | 0.001 |
| ADPRH | [Protein ADP-ribosylarginine] hydrolase | 1.018 | 4.849 | <0.001 | 0.002 |
| AGER | Advanced glycosylation end product-specific receptor; soluble | -1.140 | -3.713 | 0.001 | 0.006 |
| AK1 | Adenylate kinase isoenzyme 1 | 1.886 | 3.120 | 0.004 | 0.018 |
| ALDH1A1 | Retinal dehydrogenase 1 | 1.144 | 4.304 | <0.001 | 0.003 |
| ANPEP | Aminopeptidase N | 1.320 | 3.541 | 0.001 | 0.008 |
| AP1AR | AP-1 complex-associated regulatory protein | 1.365 | 3.946 | <0.001 | 0.004 |
| APAF1 | Apoptotic protease-activating factor 1 | -1.154 | -3.308 | 0.002 | 0.013 |
| APOA1 | Apolipoprotein A-I | -1.579 | -3.284 | 0.002 | 0.013 |
| APOB | Apolipoprotein B | -1.574 | -2.639 | 0.013 | 0.044 |
| APOBEC2 | C->U-editing enzyme APOBEC-2 | 1.143 | 5.009 | <0.001 | 0.001 |
| APOC1 | Apolipoprotein C-I | 2.621 | 3.518 | 0.001 | 0.009 |
| APOD | Apolipoprotein D | 1.457 | 4.119 | <0.001 | 0.003 |
| APOE | Apolipoprotein E | 1.712 | 4.010 | <0.001 | 0.004 |
| ARRB1 | Beta-arrestin-1 | 1.014 | 2.578 | 0.015 | 0.049 |
| ASL | Argininosuccinate lyase | 1.898 | 6.107 | <0.001 | <0.001 |
| ATF6B | Cyclic AMP-dependent transcription factor ATF-6 beta | 1.710 | 4.199 | <0.001 | 0.003 |
| ATG7 | Ubiquitin-like modifier-activating enzyme ATG7 | 1.219 | 3.030 | 0.005 | 0.021 |
| ATP2A3 | Sarcoplasmic/endoplasmic reticulum calcium ATPase 3 | -1.012 | -3.775 | 0.001 | 0.006 |
| ATP4B | Potassium-transporting ATPase subunit beta | 1.578 | 4.301 | <0.001 | 0.003 |
| BAMBI | BMP and activin membrane-bound inhibitor homolog:Extracellular domain | -1.222 | -2.899 | 0.007 | 0.027 |
| BGLAP | Osteocalcin | 1.101 | 3.153 | 0.003 | 0.017 |
| BICD1 | Protein bicaudal D homolog 1 | 1.192 | 3.388 | 0.002 | 0.011 |
| BMP5 | Bone morphogenetic protein 5 | -1.059 | -5.335 | <0.001 | 0.001 |
| BPIFB1 | BPI fold-containing family B member 1 | 1.346 | 3.331 | 0.002 | 0.012 |
| C14orf93 | Uncharacterized protein C14orf93 | 1.258 | 4.705 | <0.001 | 0.002 |
| C1R | Complement C1r subcomponent | -1.691 | -3.873 | <0.001 | 0.005 |
| C4A\|C4B | Complement C4 | 1.377 | 5.163 | <0.001 | 0.001 |
| CA11 | Carbonic anhydrase-related protein 11 | -1.179 | -8.336 | <0.001 | <0.001 |
| CAP1 | Adenylyl cyclase-associated protein 1 | 1.236 | 2.603 | 0.014 | 0.047 |
| CCN4 | WNT1-inducible-signaling pathway protein 1 | 1.204 | 4.318 | 0.000 | 0.003 |
| CDC25A | M-phase inducer phosphatase 1 | -2.082 | -2.788 | 0.009 | 0.033 |
| CDH23 | Cadherin-23 | 1.003 | 3.036 | 0.005 | 0.021 |
| CDH3 | Cadherin-3 | 1.597 | 4.053 | <0.001 | 0.004 |
| CDH6 | Cadherin-6 | 1.435 | 4.386 | <0.001 | 0.003 |
| CFC1 | Cryptic protein | -1.231 | -3.466 | 0.001 | 0.010 |
| CFHR4 | Complement factor H-related protein 4 | 1.890 | 2.588 | 0.014 | 0.048 |
| CHAD | Chondroadherin | -1.021 | -3.379 | 0.002 | 0.011 |

| | | | | | |
|---|---|---|---|---|---|
| CHKB | Choline/ethanolamine kinase | 2.988 | 4.713 | <0.001 | 0.002 |
| CLPSL2 | Colipase-like protein 2 | 1.388 | 4.239 | <0.001 | 0.003 |
| CLU | Clusterin | 1.003 | 2.870 | 0.007 | 0.029 |
| CNDP1 | Beta-Ala-His dipeptidase | 1.811 | 5.257 | <0.001 | 0.001 |
| COMMD8 | COMM domain-containing protein 8 | -1.486 | -5.447 | <0.001 | 0.001 |
| COQ9 | Ubiquinone biosynthesis protein COQ9; mitochondrial | 1.264 | 2.842 | 0.008 | 0.030 |
| CP | Ceruloplasmin | -1.246 | -5.149 | <0.001 | 0.001 |
| CPA2 | Carboxypeptidase A2 | 1.018 | 3.176 | 0.003 | 0.016 |
| CPXM1 | Probable carboxypeptidase X1 | 1.295 | 3.053 | 0.004 | 0.020 |
| CRELD1 | Cysteine-rich with EGF-like domain protein 1 | 1.027 | 3.644 | 0.001 | 0.007 |
| CRIP1 | Cysteine-rich protein 1 | 1.139 | 2.875 | 0.007 | 0.028 |
| CRISPLD2 | Cysteine-rich secretory protein LCCL domain-containing 2 | 1.648 | 5.591 | <0.001 | 0.001 |
| CRP | C-reactive protein | 1.201 | 2.748 | 0.010 | 0.036 |
| CSN2 | Beta-casein | 1.441 | 3.769 | 0.001 | 0.006 |
| CSNK2A1 | Casein kinase II subunit alpha | -1.113 | -3.484 | 0.001 | 0.009 |
| CTAG1A\|CTAG1B | Cancer/testis antigen 1 | 1.028 | 5.064 | <0.001 | 0.001 |
| CTSA | Lysosomal protective protein | 2.386 | 4.836 | <0.001 | 0.002 |
| DDRGK1 | DDRGK domain-containing protein 1 | 1.029 | 3.923 | <0.001 | 0.004 |
| DNAJC4 | DnaJ homolog subfamily C member 4:C-term | 1.416 | 4.096 | <0.001 | 0.004 |
| EFNB3 | Ephrin-B3:Cytoplasmic domain | 1.331 | 4.131 | <0.001 | 0.003 |
| EGLN2 | Egl nine homolog 2 | 1.048 | 4.153 | <0.001 | 0.003 |
| EHMT2 | Histone-lysine N-methyltransferase EHMT2 | -1.041 | -3.772 | 0.001 | 0.006 |
| EIF1AY | Eukaryotic translation initiation factor 1A; Y-chromosomal | 1.368 | 2.686 | 0.011 | 0.040 |
| EIF4A2 | Eukaryotic initiation factor 4A-II | 1.455 | 2.760 | 0.009 | 0.035 |
| EMC4 | ER membrane protein complex subunit 4 | 1.608 | 4.520 | <0.001 | 0.002 |
| ENO1 | Alpha-enolase | 2.356 | 4.879 | <0.001 | 0.002 |
| ENPEP | Glutamyl aminopeptidase | -1.642 | -3.882 | <0.001 | 0.005 |
| ETNK2 | Ethanolamine kinase 2 | -1.310 | -2.795 | 0.009 | 0.033 |
| F9 | Coagulation factor IX | 1.194 | 3.989 | <0.001 | 0.004 |
| FABP3 | Fatty acid-binding protein; heart | 1.754 | 5.123 | <0.001 | 0.001 |
| FAM110A | Protein FAM110A | -1.351 | -5.227 | <0.001 | 0.001 |
| FAM177A1 | Protein FAM177A1 | 1.653 | 3.887 | <0.001 | 0.005 |
| FAM210A | Protein FAM210A | 1.266 | 4.160 | <0.001 | 0.003 |
| FAM3B | Protein FAM3B | -1.041 | -6.072 | <0.001 | <0.001 |
| FAM89B | Leucine repeat adapter protein 25 | 1.170 | 4.256 | <0.001 | 0.003 |
| FBLIM1 | Filamin-binding LIM protein 1 | 1.367 | 3.358 | 0.002 | 0.012 |
| FCGR2B | Low affinity immunoglobulin gamma Fc region receptor II-b | -1.132 | -3.390 | 0.002 | 0.011 |
| FGD2 | FYVE; RhoGEF and PH domain-containing protein 2 | 1.085 | 4.199 | <0.001 | 0.003 |
| FGFBP1 | Fibroblast growth factor-binding protein 1 | 1.543 | 4.000 | <0.001 | 0.004 |
| FKBP1A | FK506 binding protein 1A | -1.078 | -2.770 | 0.009 | 0.034 |
| FLT3 | Receptor-type tyrosine-protein kinase FLT3 | 1.070 | 4.561 | <0.001 | 0.002 |
| FLT4 | Vascular endothelial growth factor receptor 3 | 1.156 | 3.478 | 0.001 | 0.009 |
| FMOD | fibromodulin | -1.080 | -6.244 | <0.001 | <0.001 |
| FTH1\|FTL | Ferritin | 1.630 | 3.716 | 0.001 | 0.006 |
| FTL | Ferritin light chain | 1.644 | 3.876 | <0.001 | 0.005 |
| FTMT | Ferritin; mitochondrial | 1.539 | 4.071 | <0.001 | 0.004 |
| GAGE2B | G antigen 2 | 2.067 | 4.251 | <0.001 | 0.003 |
| GAL | Galanin | 1.014 | 4.112 | <0.001 | 0.004 |
| GALE | UDP-glucose 4-epimerase | 1.728 | 4.644 | <0.001 | 0.002 |
| GALNT14 | Polypeptide N-acetylgalactosaminyltransferase 14 | -1.066 | -3.623 | 0.001 | 0.007 |
| GEM | GTP-binding protein GEM | 1.267 | 3.682 | 0.001 | 0.006 |
| GEMIN7 | Gem-associated protein 7 | 1.471 | 4.327 | <0.001 | 0.003 |
| GIP | Gastric inhibitory polypeptide | 1.026 | 3.903 | <0.001 | 0.005 |
| GOSR2 | Golgi SNAP receptor complex member 2 | 1.158 | 3.042 | 0.005 | 0.021 |
| GPD1 | Glycerol-3-phosphate dehydrogenase [NAD(+)]; cytoplasmic | 1.882 | 4.421 | 0.000 | 0.003 |
| GSTM4 | Glutathione S-transferase Mu 4 | -1.039 | -3.224 | 0.003 | 0.015 |
| GULP1 | PTB domain-containing engulfment adapter protein 1 | 1.333 | 3.875 | <0.001 | 0.005 |
| H2AC1 | Histone H2A type 1-A | 1.303 | 3.073 | 0.004 | 0.019 |

| | | | | | |
|---|---|---|---|---|---|
| H2BC12 | Histone H2B type 1-K | 1.441 | 3.348 | 0.002 | 0.012 |
| H2BU1 | Histone H2B type 3-B | 1.818 | 3.823 | 0.001 | 0.005 |
| HAAO | 3-hydroxyanthranilate 3;4-dioxygenase | 1.373 | 4.228 | <0.001 | 0.003 |
| HAMP | Hepcidin | 1.448 | 3.054 | 0.004 | 0.020 |
| HBE1 | Hemoglobin subunit epsilon | -1.011 | -4.019 | <0.001 | 0.004 |
| HCLS1 | Hematopoietic lineage cell-specific protein | 2.005 | 4.637 | <0.001 | 0.002 |
| HLA-E | HLA class I histocompatibility antigen; alpha chain E | 1.089 | 3.959 | <0.001 | 0.004 |
| HMGCS1 | Hydroxymethylglutaryl-CoA synthase; cytoplasmic | 1.041 | 4.381 | <0.001 | 0.003 |
| HNRNPAB | Heterogeneous nuclear ribonucleoprotein A/B | 1.178 | 4.019 | <0.001 | 0.004 |
| IAPP | Islet amyloid polypeptide | -1.513 | -4.750 | <0.001 | 0.002 |
| IBSP | Bone sialoprotein 2 | 1.529 | 3.224 | 0.003 | 0.015 |
| ICA1 | Islet cell autoantigen 1 | 1.246 | 4.021 | <0.001 | 0.004 |
| IDH1 | Isocitrate dehydrogenase [NADP] cytoplasmic | 1.849 | 6.620 | <0.001 | <0.001 |
| IFNG | Interferon gamma | 1.127 | 4.324 | <0.001 | 0.003 |
| IFT22 | Intraflagellar transport protein 22 homolog | -1.118 | -4.841 | <0.001 | 0.002 |
| IGFBP1 | Insulin-like growth factor-binding protein 1 | -1.353 | -4.726 | <0.001 | 0.002 |
| IGL\|IGHD\|IGK | Immunoglobulin D | 2.016 | 3.221 | 0.003 | 0.015 |
| IL2 | Interleukin-2 | -1.046 | -3.461 | 0.001 | 0.010 |
| IL36A | Interleukin-36 alpha | 2.678 | 4.694 | <0.001 | 0.002 |
| IMMP2L | Mitochondrial inner membrane protease subunit 2 | -1.154 | -3.254 | 0.003 | 0.014 |
| INHBC | Inhibin beta C chain | 1.706 | 4.327 | <0.001 | 0.003 |
| IQCF1 | IQ domain-containing protein F1 | 1.600 | 4.129 | <0.001 | 0.003 |
| IRF6 | Interferon regulatory factor 6 | -1.275 | -4.321 | <0.001 | 0.003 |
| ITGA11\|ITGB1 | Integrin a11b1 | -1.064 | -4.996 | <0.001 | 0.001 |
| ITGAV\|ITGB3 | Integrin alpha V beta 3 | -1.017 | -5.114 | <0.001 | 0.001 |
| ITGB7 | Integrin beta-7 | 1.320 | 5.295 | <0.001 | 0.001 |
| ITIH4 | Inter-alpha-trypsin inhibitor heavy chain H4 | 1.932 | 3.047 | 0.005 | 0.020 |
| ITM2B | Integral membrane protein 2B | -1.314 | -4.060 | <0.001 | 0.004 |
| JPT1 | Hematological and neurological expressed 1 protein | 1.153 | 3.441 | 0.002 | 0.010 |
| KCNA10 | Potassium voltage-gated channel subfamily A member 10 | -1.130 | -5.839 | <0.001 | <0.001 |
| KCNAB3 | Voltage-gated potassium channel subunit beta-3 | 1.118 | 3.617 | 0.001 | 0.007 |
| KCNE5 | Potassium voltage-gated channel subfamily E regulatory beta subunit 5:Cytoplasmic domain | 1.004 | 4.474 | <0.001 | 0.002 |
| KHSRP | Far upstream element-binding protein 2 | 1.055 | 4.425 | <0.001 | 0.003 |
| KLB | Beta-klotho | 1.026 | 4.535 | <0.001 | 0.002 |
| L1CAM | Neural cell adhesion molecule L1 | -1.045 | -5.753 | <0.001 | <0.001 |
| LAG3 | Lymphocyte activation gene 3 protein | 1.614 | 3.714 | 0.001 | 0.006 |
| LAP3 | Cytosol aminopeptidase | 1.208 | 3.335 | 0.002 | 0.012 |
| LAT | Linker for activation of T-cells family member 1 | 1.076 | 2.585 | 0.014 | 0.048 |
| LDHA | L-lactate dehydrogenase A chain | 1.002 | 3.922 | 0.000 | 0.004 |
| LGALS3BP | Galectin-3-binding protein | 1.221 | 3.618 | 0.001 | 0.007 |
| LGMN | Legumain | 1.371 | 5.491 | <0.001 | 0.001 |
| LIMK1 | LIM domain kinase 1 | 1.510 | 4.748 | <0.001 | 0.002 |
| LMAN2 | Vesicular integral-membrane protein VIP36 | 1.991 | 3.231 | 0.003 | 0.014 |
| LRRN1 | Leucine-rich repeat neuronal protein 1:Extracellular domain | -1.077 | -4.676 | <0.001 | 0.002 |
| LSP1 | Lymphocyte-specific protein 1 | 1.192 | 4.533 | <0.001 | 0.002 |
| MACROD2 | O-acetyl-ADP-ribose deacetylase MACROD2 | -1.763 | -4.604 | <0.001 | 0.002 |
| MAGEA4 | Melanoma-associated antigen 4 | -1.531 | -3.504 | 0.001 | 0.009 |
| MAK16 | Protein MAK16 homolog | 1.320 | 3.945 | <0.001 | 0.004 |
| MAP3K11 | Mitogen-activated protein kinase kinase kinase 11 | -1.106 | -3.864 | <0.001 | 0.005 |
| MARCKSL1 | MARCKS-related protein | 1.443 | 4.718 | <0.001 | 0.002 |
| MAT1A | S-adenosylmethionine synthase isoform type-1 | -1.210 | -3.192 | 0.003 | 0.016 |
| MAT2A | S-adenosylmethionine synthetase isoform type-2 | 1.172 | 3.869 | <0.001 | 0.005 |
| MBD1 | Methyl-CpG-binding domain protein 1 | 2.037 | 4.589 | <0.001 | 0.002 |
| MMP13 | Collagenase 3 | -1.073 | -4.610 | <0.001 | 0.002 |
| MMP8 | Neutrophil collagenase | 1.067 | 2.736 | 0.010 | 0.036 |
| MOB1B | MOB kinase activator 1B | 1.462 | 4.259 | <0.001 | 0.003 |
| MRPL21 | 39S ribosomal protein L21; mitochondrial | 1.653 | 4.620 | <0.001 | 0.002 |
| MRPL58 | Peptidyl-tRNA hydrolase ICT1; mitochondrial | 1.718 | 2.946 | 0.006 | 0.025 |

| | | | | | |
|---|---|---|---|---|---|
| MST1 | Hepatocyte growth factor-like protein | 1.158 | 2.586 | 0.014 | 0.048 |
| MUSK | Muscle; skeletal receptor tyrosine-protein kinase | -1.238 | -2.979 | 0.005 | 0.023 |
| MX1 | Interferon-induced GTP-binding protein Mx1 | 1.040 | 2.998 | 0.005 | 0.022 |
| NAALADL1 | N-acetylated-alpha-linked acidic dipeptidase-like protein | 1.094 | 3.856 | 0.001 | 0.005 |
| NAB1 | NGFI-A-binding protein 1 | 2.284 | 4.493 | <0.001 | 0.002 |
| NANS | Sialic acid synthase | 1.036 | 2.763 | 0.009 | 0.035 |
| NAP1L1 | Nucleosome assembly protein 1-like 1 | 1.109 | 2.609 | 0.013 | 0.046 |
| NCF1 | Neutrophil cytosol factor 1 | 1.757 | 4.996 | <0.001 | 0.001 |
| NCF4 | Neutrophil cytosol factor 4 | 1.116 | 4.100 | <0.001 | 0.004 |
| NCMAP | Noncompact myelin-associated protein | 1.415 | 3.686 | 0.001 | 0.006 |
| NRGN | Neurogranin | 1.552 | 3.241 | 0.003 | 0.014 |
| NUDT16 | U8 snoRNA-decapping enzyme | 1.591 | 4.385 | <0.001 | 0.003 |
| OLFML3 | Olfactomedin-like protein 3 | 1.792 | 3.440 | 0.002 | 0.010 |
| OLR1 | Oxidized low-density lipoprotein receptor 1 | 1.093 | 3.781 | 0.001 | 0.006 |
| OPTC | Opticin | 1.110 | 4.133 | <0.001 | 0.003 |
| OTC | Ornithine carbamoyltransferase; mitochondrial | 1.277 | 3.348 | 0.002 | 0.012 |
| PADI4 | Protein-arginine deiminase type-4 | 2.296 | 4.117 | <0.001 | 0.003 |
| PAM | Peptidyl-glycine alpha-amidating monooxygenase | 1.220 | 2.792 | 0.009 | 0.033 |
| PARP16 | Mono [ADP-ribose] polymerase PARP16 | 1.578 | 4.695 | <0.001 | 0.002 |
| PCDHB2 | Protocadherin beta-2 | -1.239 | -4.664 | <0.001 | 0.002 |
| PCDHGC3 | Protocadherin gamma-C3 | 2.821 | 4.926 | <0.001 | 0.002 |
| PCK2 | Phosphoenolpyruvate carboxykinase [GTP]; mitochondrial | -1.206 | -3.155 | 0.003 | 0.017 |
| PCSK1N | ProSAAS | -1.995 | -4.017 | <0.001 | 0.004 |
| PCSK9 | Proprotein convertase subtilisin/kexin type 9 | 1.477 | 3.696 | 0.001 | 0.006 |
| PDE4C | cAMP-specific 3';5'-cyclic phosphodiesterase 4C | 1.351 | 3.686 | 0.001 | 0.006 |
| PDLIM5 | PDZ and LIM domain protein 5 | 1.061 | 2.910 | 0.006 | 0.027 |
| PENK | Proenkephalin-A | 1.504 | 3.039 | 0.005 | 0.021 |
| PGLYRP1 | Peptidoglycan recognition protein 1 | 1.070 | 3.472 | 0.001 | 0.009 |
| PKM | Pyruvate kinase PKM | 1.062 | 3.043 | 0.005 | 0.021 |
| PLA2G4A | Cytosolic phospholipase A2 alpha | -1.353 | -3.953 | <0.001 | 0.004 |
| PMPCA | Mitochondrial-processing peptidase subunit alpha | 1.271 | 4.287 | <0.001 | 0.003 |
| POP7 | Ribonuclease P protein subunit p20 | 2.028 | 4.580 | <0.001 | 0.002 |
| PPA1 | Inorganic pyrophosphatase | 1.511 | 4.505 | <0.001 | 0.002 |
| PPM1F | Protein phosphatase 1F | 1.087 | 3.564 | 0.001 | 0.008 |
| PPP4R3A | Serine/threonine-protein phosphatase 4 regulatory subunit 3A | 1.180 | 3.617 | 0.001 | 0.007 |
| PRDX6 | Peroxiredoxin-6 | 1.704 | 2.971 | 0.005 | 0.023 |
| PRKRA | Interferon-inducible double-stranded RNA-dependent protein kinase activator A | 1.419 | 2.894 | 0.007 | 0.027 |
| PROC | Activated Protein C | -2.300 | -4.125 | <0.001 | 0.003 |
| PROC | Activated Protein C | 1.088 | 3.826 | 0.001 | 0.005 |
| PRTN3 | Myeloblastin | 1.315 | 3.922 | <0.001 | 0.004 |
| PSG2 | Pregnancy-specific beta-1-glycoprotein 2 | -1.114 | -3.154 | 0.003 | 0.017 |
| PSMB9 | Proteasome subunit beta type-9 | -1.193 | -4.834 | <0.001 | 0.002 |
| PTGR2 | Prostaglandin reductase 2 | -1.673 | -4.421 | <0.001 | 0.003 |
| PYGL | Glycogen phosphorylase; liver form | 1.145 | 3.040 | 0.005 | 0.021 |
| PZP | Pregnancy zone protein | 1.619 | 2.764 | 0.009 | 0.035 |
| RCAN1 | Calcipressin-1 | 1.450 | 3.491 | 0.001 | 0.009 |
| REG3A | Regenerating islet-derived protein 3-alpha | -1.245 | -4.552 | <0.001 | 0.002 |
| RNASE3 | Eosinophil cationic protein | 1.371 | 4.513 | <0.001 | 0.002 |
| ROR1 | Inactive tyrosine-protein kinase transmembrane receptor ROR1 | -1.065 | -6.490 | <0.001 | <0.001 |
| RPP25 | Ribonuclease P protein subunit p25 | -1.827 | -5.075 | <0.001 | 0.001 |
| RUFY1 | RUN and FYVE domain-containing protein 1 | 1.168 | 4.009 | <0.001 | 0.004 |
| SAA1 | Serum amyloid A-1 protein | 1.783 | 3.553 | 0.001 | 0.008 |
| SELENOS | Selenoprotein S | 1.445 | 4.886 | <0.001 | 0.002 |
| SEMA3G | Semaphorin-3G | 1.135 | 4.085 | <0.001 | 0.004 |
| SEMA4G | Semaphorin-4G | 1.215 | 5.268 | <0.001 | 0.001 |
| SERPINA10 | Protein Z-dependent protease inhibitor | 1.150 | 2.712 | 0.011 | 0.038 |
| SERPINF2 | Alpha-2-antiplasmin | 1.962 | 4.749 | <0.001 | 0.002 |

| | | | | | |
|---|---|---|---|---|---|
| SERPING1 | Plasma protease C1 inhibitor | 1.924 | 3.100 | 0.004 | 0.019 |
| SFTPD | Pulmonary surfactant-associated protein D | 1.720 | 3.416 | 0.002 | 0.010 |
| SIGLEC12 | Sialic acid-binding Ig-like lectin 12:Ig-like C2-type 2 domain; Isoform short | 2.065 | 4.434 | <0.001 | 0.003 |
| SLC22A16 | Solute carrier family 22 member 16 | 1.485 | 3.014 | 0.005 | 0.022 |
| SLC35G2 | Solute carrier family 35 member G2 | 2.064 | 4.010 | <0.001 | 0.004 |
| SLC6A14 | Sodium- and chloride-dependent neutral and basic amino acid transporter B(0+) | 1.009 | 2.879 | 0.007 | 0.028 |
| SMPDL3A | Acid sphingomyelinase-like phosphodiesterase 3a | 1.059 | 4.702 | <0.001 | 0.002 |
| SNCA | Alpha-synuclein | 1.612 | 2.884 | 0.007 | 0.028 |
| SNX3 | Sorting nexin-3 | 1.160 | 4.439 | <0.001 | 0.003 |
| SORD | Sorbitol dehydrogenase | 1.277 | 4.586 | <0.001 | 0.002 |
| STK24 | Serine/threonine-protein kinase 24 | 1.297 | 3.213 | 0.003 | 0.015 |
| STX17 | Syntaxin-17 | -1.047 | -4.188 | <0.001 | 0.003 |
| TAF8 | Transcription initiation factor TFIID subunit 8 | 1.029 | 3.728 | 0.001 | 0.006 |
| TBC1D5 | TBC1 domain family member 5 | 1.073 | 4.416 | <0.001 | 0.003 |
| TEX29 | Testis-expressed sequence 29 protein | -1.084 | -3.425 | 0.002 | 0.010 |
| THRB | Thyroid hormone receptor beta | -1.028 | -4.457 | <0.001 | 0.002 |
| TK2 | Thymidine kinase 2; mitochondrial | -1.055 | -3.476 | 0.001 | 0.009 |
| TKFC | Dihydroxyacetone kinase | 1.058 | 3.154 | 0.003 | 0.017 |
| TMEM59L | Transmembrane protein 59-like | -1.016 | -4.365 | <0.001 | 0.003 |
| TNFRSF14 | Tumor necrosis factor receptor superfamily member 14 | 1.171 | 4.045 | <0.001 | 0.004 |
| TNNI2 | Troponin I; fast skeletal muscle | 1.382 | 5.026 | <0.001 | 0.001 |
| TP53I11 | Tumor protein p53-inducible protein 11 | 1.539 | 5.323 | <0.001 | 0.001 |
| TPD52L2 | Tumor protein D54 | 1.271 | 2.805 | 0.008 | 0.032 |
| TRIO | Triple functional domain protein | 1.136 | 3.022 | 0.005 | 0.021 |
| TRMT6 | tRNA (adenine(58)-N(1))-methyltransferase non-catalytic subunit TRM6 | 1.506 | 5.428 | <0.001 | 0.001 |
| TYMSOS | TYMS opposite strand protein | 1.234 | 4.246 | <0.001 | 0.003 |
| UBE2D3 | Ubiquitin-conjugating enzyme E2 D3 | 1.024 | 4.693 | <0.001 | 0.002 |
| VAT1 | Synaptic vesicle membrane protein VAT-1 homolog | 1.186 | 3.266 | 0.003 | 0.014 |
| VAV3 | Guanine nucleotide exchange factor VAV3 | 1.046 | 5.366 | <0.001 | 0.001 |
| VCX | Variable charge X-linked protein 1 | 1.585 | 3.853 | 0.001 | 0.005 |
| WDR18 | WD repeat-containing protein 18 | 1.312 | 3.273 | 0.002 | 0.013 |
| WNT10A | Protein Wnt-10a | 1.962 | 4.122 | <0.001 | 0.003 |
| WNT3A | Protein Wnt-3a | 1.946 | 4.348 | <0.001 | 0.003 |
| ZC3H8 | Zinc finger CCCH domain-containing protein 8 | 1.039 | 3.989 | <0.001 | 0.004 |

**Supplementary Table 4. Differentially abundant proteins (*n* = 421) between the recovery phase of Guillain-Barré syndrome and healthy controls.** $Log_2$ fold-change ($Log_2FC$) indicates the magnitude of relative protein abundance. The moderated t-statistic is calculated using empirical Bayes estimation, and reflects the difference standardized by a variance estimate. For both metrics, positive values denote higher abundance in the recovery phase of Guillain-Barré syndrome, while negative values denote higher abundance in healthy controls. *P*-values were adjusted for multiple testing using the Benjamini-Hochberg false discovery rate (FDR) method. Proteins are listed in alphabetical order by gene symbol.

| Entrez Gene Symbol | Target Full Name | logFC | t | *P*-Value | FDR |
|---|---|---|---|---|---|
| ACADSB | Short/branched chain specific acyl-CoA dehydrogenase; mitochondrial | -1.465 | -4.733 | <0.001 | 0.001 |
| ACBD6 | Acyl-CoA-binding domain-containing protein 6 | 1.770 | 5.463 | <0.001 | <0.001 |
| ACP2 | Lysosomal acid phosphatase | -1.090 | -3.760 | 0.001 | 0.004 |
| ACP5 | Tartrate-resistant acid phosphatase type 5 | 1.536 | 7.556 | <0.001 | <0.001 |
| ACTN2 | Alpha-actinin-2 | 1.096 | 4.050 | <0.001 | 0.002 |
| ACY1 | Aminoacylase-1 | 1.735 | 5.056 | <0.001 | <0.001 |
| ADH1C | Alcohol dehydrogenase 1C | 1.590 | 5.510 | <0.001 | <0.001 |
| ADM | Adrenomedullin | -1.210 | -4.545 | <0.001 | 0.001 |
| AGT | Angiotensinogen | 1.004 | 2.791 | 0.009 | 0.028 |
| AK1 | Adenylate kinase isoenzyme 1 | 2.618 | 4.311 | <0.001 | 0.001 |
| ALDH1A1 | Retinal dehydrogenase 1 | 1.265 | 4.950 | <0.001 | <0.001 |
| ALDOA | Fructose-bisphosphate aldolase A | 1.218 | 4.220 | <0.001 | 0.002 |
| ALDOC | Fructose-bisphosphate aldolase C | 1.181 | 4.584 | <0.001 | 0.001 |
| ALPL | Alkaline phosphatase; tissue-nonspecific isozyme | -1.307 | -4.756 | <0.001 | 0.001 |
| ANPEP | Aminopeptidase N | 1.836 | 5.552 | <0.001 | <0.001 |
| ANTXR2 | Anthrax toxin receptor 2 | 1.358 | 5.833 | <0.001 | <0.001 |
| ANXA11 | Annexin A11 | -1.632 | -5.001 | <0.001 | <0.001 |
| ANXA7 | Annexin A7 | -2.187 | -7.428 | <0.001 | <0.001 |
| AOC1 | Amiloride-sensitive amine oxidase [copper-containing] | 1.231 | 3.537 | 0.001 | 0.006 |
| AP1AR | AP-1 complex-associated regulatory protein | 1.388 | 3.909 | <0.001 | 0.003 |
| AP1B1 | AP-1 complex subunit beta-1 | -1.327 | -4.216 | <0.001 | 0.002 |
| APAF1 | Apoptotic protease-activating factor 1 | -1.343 | -3.976 | <0.001 | 0.003 |
| APOA4 | Apolipoprotein A-IV | 3.211 | 5.228 | <0.001 | <0.001 |
| APOB | Apolipoprotein B | -2.640 | -4.455 | <0.001 | 0.001 |
| APOC1 | Apolipoprotein C-I | 4.588 | 7.961 | <0.001 | <0.001 |
| APOD | Apolipoprotein D | 1.327 | 3.710 | 0.001 | 0.004 |
| APOE | Apolipoprotein E | 2.141 | 5.486 | <0.001 | <0.001 |
| APOF | Apolipoprotein F | 3.401 | 6.363 | <0.001 | <0.001 |
| ARC | Activity-regulated cytoskeleton-associated protein | -1.106 | -4.095 | <0.001 | 0.002 |
| ARHGAP1 | Rho GTPase-activating protein 1:Cellular retinaldehyde-TRIO domain | -1.344 | -4.664 | <0.001 | 0.001 |
| ARL5B | ADP-ribosylation factor-like protein 5B | -1.178 | -5.645 | <0.001 | <0.001 |
| ARPC3 | Actin-related protein 2/3 complex subunit 3 | -1.071 | -3.536 | 0.001 | 0.006 |
| ARRB1 | Beta-arrestin-1 | 1.354 | 3.976 | <0.001 | 0.003 |
| ASL | Argininosuccinate lyase | 2.369 | 6.830 | <0.001 | <0.001 |
| ATF6B | Cyclic AMP-dependent transcription factor ATF-6 beta | 1.888 | 4.664 | <0.001 | 0.001 |
| ATG7 | Ubiquitin-like modifier-activating enzyme ATG7 | 1.546 | 4.093 | <0.001 | 0.002 |
| ATP2A3 | Sarcoplasmic/endoplasmic reticulum calcium ATPase 3 | -1.051 | -3.870 | <0.001 | 0.003 |
| ATP4B | Potassium-transporting ATPase subunit beta | 2.012 | 5.773 | <0.001 | <0.001 |
| ATP5PO | ATP synthase subunit O; mitochondrial | -1.046 | -2.678 | 0.011 | 0.034 |
| BAMBI | BMP and activin membrane-bound inhibitor homolog:Extracellular domain | -1.560 | -3.887 | <0.001 | 0.003 |
| BICD1 | Protein bicaudal D homolog 1 | 1.259 | 3.759 | 0.001 | 0.004 |
| BIN2 | Bridging integrator 2 | -1.080 | -3.203 | 0.003 | 0.012 |
| BPIFB1 | BPI fold-containing family B member 1 | 2.081 | 5.755 | <0.001 | <0.001 |
| BZW2 | Basic leucine zipper and W2 domain-containing protein 2 | 1.108 | 3.484 | 0.001 | 0.007 |

| | | | | | |
|---|---|---|---|---|---|
| C14orf93 | Uncharacterized protein C14orf93 | 1.128 | 4.231 | <0.001 | 0.002 |
| C1R | Complement C1r subcomponent | -1.814 | -4.997 | <0.001 | <0.001 |
| C1orf226 | Uncharacterized protein C1orf226 | -1.224 | -4.892 | <0.001 | <0.001 |
| C4A\|C4B | Complement C4 | 1.860 | 7.749 | <0.001 | <0.001 |
| C5 | C5a anaphylatoxin | -1.775 | -6.440 | <0.001 | <0.001 |
| C5orf38 | Protein CEI | 1.173 | 3.987 | <0.001 | 0.003 |
| C6orf89 | Bombesin receptor-activated protein C6orf89 | -1.077 | -5.857 | <0.001 | <0.001 |
| CAMP | Antibacterial protein LL-37 | 1.123 | 5.279 | <0.001 | <0.001 |
| CAND1 | Cullin-associated NEDD8-dissociated protein 1 | 1.378 | 4.779 | <0.001 | 0.001 |
| CAP1 | Adenylyl cyclase-associated protein 1 | 1.372 | 2.875 | 0.007 | 0.024 |
| CASP10 | Caspase-10:region 1 | -1.180 | -5.744 | <0.001 | <0.001 |
| CBR1 | Carbonyl reductase [NADPH] 1 | -1.005 | -6.088 | <0.001 | <0.001 |
| CCN4 | WNT1-inducible-signaling pathway protein 1 | 1.082 | 3.821 | 0.001 | 0.004 |
| CD3E | T-cell surface glycoprotein CD3 epsilon chain | 1.109 | 4.737 | <0.001 | 0.001 |
| CDC25A | M-phase inducer phosphatase 1 | -2.980 | -4.313 | <0.001 | 0.001 |
| CDC42BPA | Serine/threonine-protein kinase MRCK alpha | -1.317 | -3.851 | 0.001 | 0.003 |
| CDH23 | Cadherin-23 | 1.045 | 3.203 | 0.003 | 0.012 |
| CDH3 | Cadherin-3 | 1.727 | 4.356 | <0.001 | 0.001 |
| CDH6 | Cadherin-6 | 1.529 | 4.848 | <0.001 | 0.001 |
| CELA2A | Chymotrypsin-like elastase family member 2A | -1.012 | -4.227 | <0.001 | 0.002 |
| CETN2 | Centrin-2 | -1.009 | -5.106 | <0.001 | <0.001 |
| CFC1 | Cryptic protein | -1.323 | -3.708 | 0.001 | 0.004 |
| CFHR4 | Complement factor H-related protein 4 | 1.965 | 2.757 | 0.009 | 0.029 |
| CHAC1 | Glutathione-specific gamma-glutamylcyclotransferase 1 | -1.017 | -3.775 | 0.001 | 0.004 |
| CHGA | Chromogranin-A | 2.544 | 3.649 | 0.001 | 0.005 |
| CHKB | Choline/ethanolamine kinase | 3.072 | 4.939 | <0.001 | <0.001 |
| CHMP2B | Charged multivesicular body protein 2b | 1.329 | 5.131 | <0.001 | <0.001 |
| CLINT1 | Clathrin interactor 1 | -1.607 | -3.597 | 0.001 | 0.005 |
| CLPSL2 | Colipase-like protein 2 | 1.504 | 4.733 | <0.001 | 0.001 |
| CLXN | EF-hand calcium-binding domain-containing protein 1 | 1.244 | 4.543 | <0.001 | 0.001 |
| CNDP1 | Beta-Ala-His dipeptidase | 1.975 | 5.771 | <0.001 | <0.001 |
| CNRIP1 | CB1 cannabinoid receptor-interacting protein 1 | 1.070 | 3.593 | 0.001 | 0.006 |
| COL11A2 | Collagen alpha-2(XI) chain | 1.416 | 3.805 | 0.001 | 0.004 |
| COMMD10 | COMM domain-containing protein 10 | -1.087 | -3.367 | 0.002 | 0.009 |
| COMMD8 | COMM domain-containing protein 8 | -1.850 | -7.195 | <0.001 | <0.001 |
| COPS5 | COP9 signalosome complex subunit 5 | -1.688 | -4.285 | <0.001 | 0.001 |
| COQ9 | Ubiquinone biosynthesis protein COQ9; mitochondrial | 1.444 | 2.888 | 0.007 | 0.023 |
| CP | Ceruloplasmin | -1.460 | -6.205 | <0.001 | <0.001 |
| CPA2 | Carboxypeptidase A2 | 1.201 | 3.841 | 0.001 | 0.003 |
| CPLX3 | Complexin-3 | -1.077 | -5.263 | <0.001 | <0.001 |
| CPXM1 | Probable carboxypeptidase X1 | 1.468 | 3.424 | 0.002 | 0.008 |
| CRABP2 | Cellular retinoic acid-binding protein 2 | 1.833 | 6.818 | <0.001 | <0.001 |
| CREBBP | CREB-binding protein | -1.034 | -5.690 | <0.001 | <0.001 |
| CRELD1 | Cysteine-rich with EGF-like domain protein 1 | 1.320 | 4.580 | <0.001 | 0.001 |
| CRIP1 | Cysteine-rich protein 1 | 2.345 | 7.342 | <0.001 | <0.001 |
| CRKL | Crk-like protein | -1.015 | -4.982 | <0.001 | <0.001 |
| CSN2 | Beta-casein | 1.660 | 4.718 | <0.001 | 0.001 |
| CSNK2A1 | Casein kinase II subunit alpha | -1.526 | -6.076 | <0.001 | <0.001 |
| CTRB2 | Chymotrypsinogen B2 | -1.295 | -3.198 | 0.003 | 0.012 |
| CTSA | Lysosomal protective protein | 2.821 | 5.821 | <0.001 | <0.001 |
| DAPP1 | Dual adapter for phosphotyrosine and 3-phosphotyrosine and 3-phosphoinositide | -1.280 | -2.828 | 0.008 | 0.026 |
| DDRGK1 | DDRGK domain-containing protein 1 | 1.084 | 4.235 | <0.001 | 0.002 |
| DNAJC4 | DnaJ homolog subfamily C member 4:C-term | 1.595 | 4.898 | <0.001 | <0.001 |
| DNASE1L2 | Deoxyribonuclease-1-like 2 | -1.048 | -6.093 | <0.001 | <0.001 |
| DSG4 | Desmoglein-4 | 1.005 | 5.645 | <0.001 | <0.001 |
| DUS2 | tRNA-dihydrouridine(20) synthase [NAD(P)+]-like | 1.053 | 5.127 | <0.001 | <0.001 |
| EDN3 | Endothelin-3 | 1.033 | 4.903 | <0.001 | <0.001 |
| EFNB3 | Ephrin-B3:Cytoplasmic domain | 1.433 | 4.574 | <0.001 | 0.001 |

| | | | | | |
|---|---|---|---|---|---|
| EGF | Epidermal growth factor:Extracellular domain | 1.277 | 4.268 | <0.001 | 0.001 |
| EGLN2 | Egl nine homolog 2 | 1.182 | 4.905 | <0.001 | <0.001 |
| EHD3 | EH domain-containing protein 3 | -1.010 | -2.919 | 0.006 | 0.022 |
| EIF4A2 | Eukaryotic initiation factor 4A-II | 1.561 | 2.953 | 0.006 | 0.020 |
| EIF4G1 | Eukaryotic translation initiation factor 4 gamma 1 | -1.447 | -6.338 | <0.001 | <0.001 |
| EIF5 | Eukaryotic translation initiation factor 5 | -1.079 | -3.706 | 0.001 | 0.004 |
| ELK1 | ETS domain-containing protein Elk-1 | -1.074 | -6.610 | <0.001 | <0.001 |
| ELL2 | RNA polymerase II elongation factor ELL2 | -1.135 | -6.676 | <0.001 | <0.001 |
| EMC4 | ER membrane protein complex subunit 4 | 1.314 | 3.807 | 0.001 | 0.004 |
| ENO1 | Alpha-enolase | 2.227 | 5.228 | <0.001 | <0.001 |
| ENPEP | Glutamyl aminopeptidase | -2.065 | -5.295 | <0.001 | <0.001 |
| ETNK2 | Ethanolamine kinase 2 | -2.170 | -5.322 | <0.001 | <0.001 |
| FABP3 | Fatty acid-binding protein; heart | 1.897 | 6.333 | <0.001 | <0.001 |
| FAM110A | Protein FAM110A | -1.486 | -6.165 | <0.001 | <0.001 |
| FAM177A1 | Protein FAM177A1 | 2.177 | 5.480 | <0.001 | <0.001 |
| FAM210A | Protein FAM210A | 1.312 | 4.517 | <0.001 | 0.001 |
| FAM89B | Leucine repeat adapter protein 25 | 1.256 | 4.755 | <0.001 | 0.001 |
| FBLIM1 | Filamin-binding LIM protein 1 | 1.794 | 4.691 | <0.001 | 0.001 |
| FGD2 | FYVE; RhoGEF and PH domain-containing protein 2 | 1.033 | 3.971 | <0.001 | 0.003 |
| FGF8 | Fibroblast growth factor 8 isoform B | 1.149 | 4.419 | <0.001 | 0.001 |
| FGFBP1 | Fibroblast growth factor-binding protein 1 | 1.838 | 5.217 | <0.001 | <0.001 |
| FH | Fumarate hydratase; mitochondrial | -1.343 | -4.571 | <0.001 | 0.001 |
| FHOD1 | FH1/FH2 domain-containing protein 1 | -1.088 | -3.512 | 0.001 | 0.007 |
| FKBP1A | FK506 binding protein 1A | -2.039 | -6.634 | <0.001 | <0.001 |
| FLII | Protein flightless-1 homolog | -1.218 | -2.951 | 0.006 | 0.020 |
| FLNA | Filamin-A:Calponin Homology 1 | -1.450 | -5.999 | <0.001 | <0.001 |
| FLT3 | Receptor-type tyrosine-protein kinase FLT3 | 1.104 | 4.769 | <0.001 | 0.001 |
| FLT4 | Vascular endothelial growth factor receptor 3 | 1.484 | 4.662 | <0.001 | 0.001 |
| FTMT | Ferritin; mitochondrial | 1.570 | 4.279 | <0.001 | 0.001 |
| GAGE2B | G antigen 2 | 1.912 | 3.905 | <0.001 | 0.003 |
| GAL | Galanin | 1.079 | 4.557 | <0.001 | 0.001 |
| GALE | UDP-glucose 4-epimerase | 1.553 | 4.036 | <0.001 | 0.002 |
| GBP1 | Guanylate-binding protein 1 | 1.388 | 4.117 | <0.001 | 0.002 |
| GDI1 | Rab GDP dissociation inhibitor alpha | -1.096 | -7.420 | <0.001 | <0.001 |
| GEM | GTP-binding protein GEM | 1.219 | 3.274 | 0.002 | 0.011 |
| GEMIN7 | Gem-associated protein 7 | 1.390 | 3.976 | <0.001 | 0.003 |
| GIP | Gastric inhibitory polypeptide | 1.002 | 3.872 | <0.001 | 0.003 |
| GOSR2 | Golgi SNAP receptor complex member 2 | 1.483 | 4.352 | <0.001 | 0.001 |
| GOT1 | Aspartate aminotransferase; cytoplasmic | -1.199 | -6.013 | <0.001 | <0.001 |
| GPC6 | Glypican-6 | -1.424 | -4.637 | <0.001 | 0.001 |
| GPD1 | Glycerol-3-phosphate dehydrogenase [NAD(+)]; cytoplasmic | 2.888 | 6.164 | <0.001 | <0.001 |
| GSTM4 | Glutathione S-transferase Mu 4 | -1.026 | -3.214 | 0.003 | 0.012 |
| GULP1 | PTB domain-containing engulfment adapter protein 1 | 1.479 | 4.485 | <0.001 | 0.001 |
| H2BU1 | Histone H2B type 3-B | 1.319 | 2.745 | 0.010 | 0.030 |
| H3C1 | Histone H3.1 | -1.169 | -3.506 | 0.001 | 0.007 |
| HAAO | 3-hydroxyanthranilate 3;4-dioxygenase | 1.621 | 5.145 | <0.001 | <0.001 |
| HBE1 | Hemoglobin subunit epsilon | -1.190 | -4.928 | <0.001 | <0.001 |
| HCLS1 | Hematopoietic lineage cell-specific protein | 1.594 | 3.956 | <0.001 | 0.003 |
| HLA-E | HLA class I histocompatibility antigen; alpha chain E | 1.031 | 3.631 | 0.001 | 0.005 |
| HMGCS1 | Hydroxymethylglutaryl-CoA synthase; cytoplasmic | 2.136 | 5.805 | <0.001 | <0.001 |
| HNRNPAB | Heterogeneous nuclear ribonucleoprotein A/B | 1.537 | 5.520 | <0.001 | <0.001 |
| HNRNPD | Heterogeneous nuclear ribonucleoprotein D0 | 1.008 | 4.123 | <0.001 | 0.002 |
| HOMER1 | Homer protein homolog 1 | -1.106 | -3.437 | 0.002 | 0.008 |
| HSDL2 | Hydroxysteroid dehydrogenase-like protein 2 | 1.050 | 4.141 | <0.001 | 0.002 |
| HSPA1A | Heat shock 70 kDa protein 1A | -1.261 | -6.779 | <0.001 | <0.001 |
| HSPB6 | Heat shock protein beta-6 | 1.230 | 3.860 | <0.001 | 0.003 |
| HTR7 | 5-hydroxytryptamine receptor 7 | -1.050 | -3.387 | 0.002 | 0.008 |
| IAPP | Islet amyloid polypeptide | -1.397 | -4.617 | <0.001 | 0.001 |

| | | | | | |
|---|---|---|---|---|---|
| IBSP | Bone sialoprotein 2 | 1.449 | 3.027 | 0.005 | 0.017 |
| ICA1 | Islet cell autoantigen 1 | 1.477 | 5.340 | <0.001 | <0.001 |
| IDH1 | Isocitrate dehydrogenase [NADP] cytoplasmic | 2.514 | 8.756 | <0.001 | <0.001 |
| IFI16 | Gamma-interferon-inducible protein 16:Isoform 2; Hematopoietic expression; interferon-inducible nature; and nuclear localization 2 | 1.098 | 3.437 | 0.002 | 0.008 |
| IFNG | Interferon gamma | 1.245 | 5.145 | <0.001 | <0.001 |
| IFT22 | Intraflagellar transport protein 22 homolog | -1.283 | -5.852 | <0.001 | <0.001 |
| IGFBP1 | Insulin-like growth factor-binding protein 1 | -1.210 | -3.633 | 0.001 | 0.005 |
| IGFBP4 | Insulin-like growth factor-binding protein 4 | 1.145 | 3.704 | 0.001 | 0.004 |
| IGFBP6 | Insulin-like growth factor-binding protein 6 | 1.099 | 5.257 | <0.001 | <0.001 |
| IGFBP7 | Insulin-like growth factor-binding protein 7 | 1.189 | 2.970 | 0.005 | 0.020 |
| IGL\|IGHD\|IGK | Immunoglobulin D | 2.077 | 3.529 | 0.001 | 0.006 |
| IL2 | Interleukin-2 | -1.390 | -4.490 | <0.001 | 0.001 |
| IL36A | Interleukin-36 alpha | 2.767 | 4.915 | <0.001 | <0.001 |
| IMMP2L | Mitochondrial inner membrane protease subunit 2 | -1.644 | -4.840 | <0.001 | 0.001 |
| INHBC | Inhibin beta C chain | 2.130 | 5.571 | <0.001 | <0.001 |
| IQCF1 | IQ domain-containing protein F1 | 1.742 | 4.617 | <0.001 | 0.001 |
| IRF6 | Interferon regulatory factor 6 | -1.646 | -6.240 | <0.001 | <0.001 |
| ITGA2B\|ITGB3 | Integrin alpha-IIb: beta-3 complex | -1.585 | -6.238 | <0.001 | <0.001 |
| ITGB7 | Integrin beta-7 | 1.120 | 4.415 | <0.001 | 0.001 |
| ITIH4 | Inter-alpha-trypsin inhibitor heavy chain H4 | 2.345 | 3.623 | 0.001 | 0.005 |
| ITM2B | Integral membrane protein 2B | -1.240 | -3.923 | <0.001 | 0.003 |
| JPT1 | Hematological and neurological expressed 1 protein | 1.077 | 3.156 | 0.003 | 0.013 |
| KATNAL1 | Katanin p60 ATPase-containing subunit A-like 1 | -1.054 | -5.559 | <0.001 | <0.001 |
| KCNA10 | Potassium voltage-gated channel subfamily A member 10 | -1.218 | -6.640 | <0.001 | <0.001 |
| KCNAB3 | Voltage-gated potassium channel subunit beta-3 | 1.226 | 4.000 | <0.001 | 0.002 |
| KCNE5 | Potassium voltage-gated channel subfamily E regulatory beta subunit 5:Cytoplasmic domain | 1.137 | 5.369 | <0.001 | <0.001 |
| KDM4C | Lysine-specific demethylase 4C | -1.015 | -2.484 | 0.018 | 0.049 |
| KHSRP | Far upstream element-binding protein 2 | 1.060 | 5.175 | <0.001 | <0.001 |
| KLB | Beta-klotho | 1.023 | 4.972 | <0.001 | <0.001 |
| LAG3 | Lymphocyte activation gene 3 protein | 2.038 | 4.667 | <0.001 | 0.001 |
| LAT | Linker for activation of T-cells family member 1 | 1.226 | 2.824 | 0.008 | 0.026 |
| LDHA | L-lactate dehydrogenase A chain | 2.967 | 6.710 | <0.001 | <0.001 |
| LEAP2 | Liver-expressed antimicrobial peptide 2 | 1.323 | 2.924 | 0.006 | 0.021 |
| LGALS3BP | Galectin-3-binding protein | 1.471 | 4.622 | <0.001 | 0.001 |
| LGMN | Legumain | 1.541 | 6.501 | <0.001 | <0.001 |
| LIMK1 | LIM domain kinase 1 | 1.512 | 4.574 | <0.001 | 0.001 |
| LMAN2 | Vesicular integral-membrane protein VIP36 | 1.928 | 3.129 | 0.004 | 0.014 |
| LRRC25 | Leucine-rich repeat-containing protein 25:Cytoplasmic domain | 1.014 | 5.063 | <0.001 | <0.001 |
| LRRN1 | Leucine-rich repeat neuronal protein 1:Extracellular domain | -1.198 | -5.218 | <0.001 | <0.001 |
| LYRM1 | LYR motif-containing protein 1 | -1.047 | -5.582 | <0.001 | <0.001 |
| LZIC | Protein LZIC | -1.073 | -6.487 | <0.001 | <0.001 |
| MACROD2 | O-acetyl-ADP-ribose deacetylase MACROD2 | -2.029 | -5.382 | <0.001 | <0.001 |
| MAGEA3 | Melanoma-associated antigen 3 | -1.348 | -5.985 | <0.001 | <0.001 |
| MAGEA4 | Melanoma-associated antigen 4 | -1.522 | -3.725 | 0.001 | 0.004 |
| MAK16 | Protein MAK16 homolog | 1.471 | 4.601 | <0.001 | 0.001 |
| MANF | Mesencephalic astrocyte-derived neurotrophic factor | -1.023 | -2.875 | 0.007 | 0.024 |
| MAP3K11 | Mitogen-activated protein kinase kinase kinase 11 | -1.333 | -5.189 | <0.001 | <0.001 |
| MAP4K1 | Mitogen-activated protein kinase kinase kinase kinase 1 | -1.431 | -3.797 | 0.001 | 0.004 |
| MAPRE1 | Microtubule-associated protein RP/EB family member 1 | -1.431 | -3.858 | <0.001 | 0.003 |
| MAPRE3 | Microtubule-associated protein RP/EB family member 3 | 1.225 | 2.728 | 0.010 | 0.031 |
| MARCKSL1 | MARCKS-related protein | 1.602 | 5.482 | <0.001 | <0.001 |
| MAT1A | S-adenosylmethionine synthase isoform type-1 | -2.076 | -8.516 | <0.001 | <0.001 |
| MBD1 | Methyl-CpG-binding domain protein 1 | 2.198 | 5.146 | <0.001 | <0.001 |
| MENT | Protein MENT | 1.088 | 4.615 | <0.001 | 0.001 |
| METAP2 | Methionine aminopeptidase 2 | 1.677 | 4.896 | <0.001 | <0.001 |
| METRNL | Meteorin-like protein | 1.098 | 6.952 | <0.001 | <0.001 |
| MLN | Promotilin | 1.260 | 3.400 | 0.002 | 0.008 |

| | | | | | |
|---|---|---|---|---|---|
| MMP13 | Collagenase 3 | -1.386 | -6.187 | <0.001 | <0.001 |
| MOB1B | MOB kinase activator 1B | 1.302 | 4.636 | <0.001 | 0.001 |
| MORF4L2 | Mortality factor 4-like protein 2 | -1.089 | -5.953 | <0.001 | <0.001 |
| MPIG6B | Protein G6b:Isoform A; Extracellular domain | -1.002 | -3.047 | 0.004 | 0.017 |
| MPZ | Myelin protein P0 | 1.004 | 5.246 | <0.001 | <0.001 |
| MRPL21 | 39S ribosomal protein L21; mitochondrial | 1.663 | 4.694 | <0.001 | 0.001 |
| MRPL58 | Peptidyl-tRNA hydrolase ICT1; mitochondrial | 2.690 | 4.073 | <0.001 | 0.002 |
| MSN | Moesin | -1.116 | -4.610 | <0.001 | 0.001 |
| MST1 | Hepatocyte growth factor-like protein | 1.723 | 3.743 | 0.001 | 0.004 |
| MUSK | Muscle; skeletal receptor tyrosine-protein kinase | -1.441 | -3.682 | 0.001 | 0.005 |
| MYL6 | Myosin light polypeptide 6 | -1.201 | -3.712 | 0.001 | 0.004 |
| NAALADL1 | N-acetylated-alpha-linked acidic dipeptidase-like protein | 1.097 | 3.796 | 0.001 | 0.004 |
| NAB1 | NGFI-A-binding protein 1 | 2.089 | 3.981 | <0.001 | 0.003 |
| NARS1 | Asparaginyl-tRNA synthetase; cytoplasmic | -1.246 | -3.449 | 0.002 | 0.007 |
| NCF1 | Neutrophil cytosol factor 1 | 1.553 | 4.626 | <0.001 | 0.001 |
| NCF2 | Neutrophil cytosol factor 2 | -2.774 | -4.996 | <0.001 | <0.001 |
| NCF4 | Neutrophil cytosol factor 4 | 1.265 | 4.899 | <0.001 | <0.001 |
| NCKIPSD | NCK-interacting protein with SH3 domain | -1.137 | -2.829 | 0.008 | 0.026 |
| NCMAP | Noncompact myelin-associated protein | 2.065 | 5.411 | <0.001 | <0.001 |
| NFKBIE | I-kappa-B-epsilon | 1.468 | 2.937 | 0.006 | 0.021 |
| NLGN1 | Neuroligin-1 | 1.299 | 5.637 | <0.001 | <0.001 |
| NMES1 | Normal mucosa of esophagus-specific gene 1 protein | -1.305 | -5.300 | <0.001 | <0.001 |
| NOTUM | Palmitoleoyl-protein carboxylesterase NOTUM | 1.402 | 4.981 | <0.001 | <0.001 |
| NPLOC4 | Nuclear protein localization protein 4 homolog | -1.083 | -3.042 | 0.005 | 0.017 |
| NPS | Neuropeptide S | 1.013 | 4.620 | 0.000 | 0.001 |
| NPW | Neuropeptide W | 1.452 | 3.460 | 0.001 | 0.007 |
| NRAS | GTPase NRas | -1.051 | -4.226 | <0.001 | 0.002 |
| NRGN | Neurogranin | 1.947 | 4.174 | <0.001 | 0.002 |
| NSFL1C | NSFL1 cofactor p47 | -1.177 | -4.904 | <0.001 | <0.001 |
| NUDT16 | U8 snoRNA-decapping enzyme | 1.576 | 4.509 | <0.001 | 0.001 |
| NUP35 | Nucleoporin NUP53 | 1.009 | 3.079 | 0.004 | 0.016 |
| OASL | 2'-5'-oligoadenylate synthetase like protein | 1.166 | 4.733 | <0.001 | 0.001 |
| OLFM3 | Noelin-3:Isoform 2; C-term | 1.084 | 5.099 | <0.001 | <0.001 |
| OLFML3 | Olfactomedin-like protein 3 | 1.649 | 4.458 | <0.001 | 0.001 |
| OLR1 | Oxidized low-density lipoprotein receptor 1 | 1.220 | 4.952 | <0.001 | <0.001 |
| OPHN1 | Oligophrenin-1 | -1.010 | -2.617 | 0.013 | 0.038 |
| OPTC | Opticin | 1.201 | 4.653 | <0.001 | 0.001 |
| OTC | Ornithine carbamoyltransferase; mitochondrial | 1.047 | 3.391 | 0.002 | 0.008 |
| OTUD5 | OTU domain-containing protein 5 | 1.064 | 4.996 | <0.001 | <0.001 |
| PA2G4 | Proliferation-associated protein 2G4 | 2.609 | 4.685 | <0.001 | 0.001 |
| PADI4 | Protein-arginine deiminase type-4 | 2.859 | 4.127 | <0.001 | 0.002 |
| PAM | Peptidyl-glycine alpha-amidating monooxygenase | 1.855 | 4.010 | <0.001 | 0.002 |
| PARK7 | Protein DJ-1 | 1.675 | 3.787 | 0.001 | 0.004 |
| PARP16 | Mono [ADP-ribose] polymerase PARP16 | 1.590 | 4.639 | <0.001 | 0.001 |
| PCDH10 | Protocadherin-10:Cytoplasmic domain | 1.010 | 3.518 | 0.001 | 0.006 |
| PCDH8 | Protocadherin-8 | 1.227 | 5.445 | <0.001 | <0.001 |
| PCDHB2 | Protocadherin beta-2 | -1.210 | -4.303 | <0.001 | 0.001 |
| PCDHB7 | Protocadherin beta-7 | 1.081 | 4.853 | <0.001 | 0.001 |
| PCDHGC3 | Protocadherin gamma-C3 | 2.930 | 5.301 | <0.001 | <0.001 |
| PCK2 | Phosphoenolpyruvate carboxykinase [GTP]; mitochondrial | -1.528 | -4.103 | <0.001 | 0.002 |
| PCSK1N | ProSAAS | -2.263 | -4.834 | <0.001 | 0.001 |
| PCSK9 | Proprotein convertase subtilisin/kexin type 9 | 2.268 | 6.595 | <0.001 | <0.001 |
| PCYOX1 | Prenylcysteine oxidase 1 | 1.449 | 7.942 | <0.001 | <0.001 |
| PDE4C | cAMP-specific 3';5'-cyclic phosphodiesterase 4C | 1.151 | 3.510 | 0.001 | 0.007 |
| PDLIM1 | PDZ and LIM domain protein 1 | -1.224 | -3.405 | 0.002 | 0.008 |
| PDZD7 | PDZ domain-containing protein 7 | 1.027 | 5.227 | <0.001 | <0.001 |
| PENK | Proenkephalin-A | 2.709 | 5.173 | <0.001 | <0.001 |
| PFKFB1 | 6-phosphofructo-2-kinase/fructose-2;6-bisphosphatase 1 | -1.463 | -4.851 | <0.001 | 0.001 |

| | | | | | |
|---|---|---|---|---|---|
| PGD | 6-phosphogluconate dehydrogenase; decarboxylating | 2.618 | 5.950 | <0.001 | <0.001 |
| PGLYRP3 | Peptidoglycan recognition protein 3 | 1.094 | 5.405 | <0.001 | <0.001 |
| PGM2 | Phosphoglucomutase-2 | -1.290 | -4.050 | <0.001 | 0.002 |
| PGP | Glycerol-3-phosphate phosphatase | 1.258 | 7.139 | <0.001 | <0.001 |
| PHPT1 | 14 kDa phosphohistidine phosphatase | 1.541 | 3.561 | 0.001 | 0.006 |
| PIP4K2A | Phosphatidylinositol 5-phosphate 4-kinase type-2 alpha | -1.386 | -3.781 | 0.001 | 0.004 |
| PIP4K2B | Phosphatidylinositol 5-phosphate 4-kinase type-2 beta | -1.192 | -3.653 | 0.001 | 0.005 |
| PKD2 | Polycystin-2:Cytoplasmic domain 1 | 1.094 | 5.581 | <0.001 | <0.001 |
| PKM | Pyruvate kinase PKM | 1.534 | 4.402 | <0.001 | 0.001 |
| PLA2G4A | Cytosolic phospholipase A2 alpha | -1.907 | -6.516 | <0.001 | <0.001 |
| PLA2G7 | Platelet-activating factor acetylhydrolase | 1.028 | 5.498 | <0.001 | <0.001 |
| PLEK | Pleckstrin | -1.053 | -3.438 | 0.002 | 0.008 |
| PLVAP | Plasmalemma vesicle-associated protein | -1.190 | -3.231 | 0.003 | 0.012 |
| PLXDC1 | Plexin domain-containing protein 1 | 1.483 | 2.739 | 0.010 | 0.030 |
| PLXNA4 | Plexin-A4 | -1.065 | -4.180 | <0.001 | 0.002 |
| PMPCA | Mitochondrial-processing peptidase subunit alpha | 1.087 | 3.490 | 0.001 | 0.007 |
| POGLUT2 | KDEL motif-containing protein 1 | 1.089 | 4.608 | <0.001 | 0.001 |
| POMK | Protein O-mannose kinase | 1.092 | 2.661 | 0.012 | 0.035 |
| PON2 | Paraoxonase 2 | -1.183 | -5.803 | <0.001 | <0.001 |
| POP7 | Ribonuclease P protein subunit p20 | 2.053 | 4.682 | <0.001 | 0.001 |
| PPA1 | Inorganic pyrophosphatase | 1.356 | 3.905 | <0.001 | 0.003 |
| PPIF | Peptidyl-prolyl cis-trans isomerase F; mitochondrial | -1.287 | -3.077 | 0.004 | 0.016 |
| PPM1A | Protein phosphatase 1A | -1.066 | -6.908 | <0.001 | <0.001 |
| PPM1F | Protein phosphatase 1F | 1.544 | 6.683 | <0.001 | <0.001 |
| PPP1R9B | Neurabin-2 | -1.235 | -3.593 | 0.001 | 0.006 |
| PRCP | Lysosomal Pro-X carboxypeptidase | 2.375 | 5.616 | <0.001 | <0.001 |
| PRDX6 | Peroxiredoxin-6 | 1.663 | 3.165 | 0.003 | 0.013 |
| PRKRA | Interferon-inducible double-stranded RNA-dependent protein kinase activator A | 1.427 | 2.710 | 0.011 | 0.032 |
| PROC | Activated Protein C | -2.997 | -5.946 | <0.001 | <0.001 |
| PROC | Activated Protein C | 1.097 | 3.918 | <0.001 | 0.003 |
| PRPSAP2 | Phosphoribosyl pyrophosphate synthase-associated protein 2 | 1.257 | 3.765 | 0.001 | 0.004 |
| PSG2 | Pregnancy-specific beta-1-glycoprotein 2 | -1.607 | -5.223 | <0.001 | <0.001 |
| PSMA1 | Proteasome subunit alpha type-1 | -1.108 | -5.589 | <0.001 | <0.001 |
| PSMA5 | Proteasome subunit alpha type-5 | -1.284 | -5.280 | <0.001 | <0.001 |
| PSMA7 | Proteasome subunit alpha type-7 | -2.289 | -6.909 | <0.001 | <0.001 |
| PSMB4 | Proteasome subunit beta type-4 | -1.735 | -7.520 | <0.001 | <0.001 |
| PSMB9 | Proteasome subunit beta type-9 | -1.385 | -5.931 | <0.001 | <0.001 |
| PSMD11 | 26S proteasome non-ATPase regulatory subunit 11 | -1.046 | -3.193 | 0.003 | 0.012 |
| PTGR2 | Prostaglandin reductase 2 | -2.397 | -6.922 | <0.001 | <0.001 |
| PTH | Parathyroid hormone | 1.032 | 4.370 | <0.001 | 0.001 |
| PTPN11 | Tyrosine-protein phosphatase non-receptor type 11 | 1.276 | 3.075 | 0.004 | 0.016 |
| PTPN6 | Tyrosine-protein phosphatase non-receptor type 6 | -1.085 | -3.228 | 0.003 | 0.012 |
| PUS7 | Pseudouridylate synthase 7 homolog | -1.407 | -4.127 | <0.001 | 0.002 |
| PXDN | Peroxidasin homolog | 1.053 | 4.780 | <0.001 | 0.001 |
| PYGB | Glycogen phosphorylase; brain form | 1.185 | 4.338 | <0.001 | 0.001 |
| PYGL | Glycogen phosphorylase; liver form | 1.654 | 5.032 | <0.001 | <0.001 |
| PZP | Pregnancy zone protein | 2.132 | 4.114 | <0.001 | 0.002 |
| QPCTL | Glutaminyl-peptide cyclotransferase-like protein | 1.125 | 8.696 | <0.001 | <0.001 |
| RALA | Ras-related protein Ral-A | -1.130 | -5.699 | <0.001 | <0.001 |
| RBM41 | RNA-binding protein 41 | -1.282 | -5.654 | <0.001 | <0.001 |
| RCAN1 | Calcipressin-1 | 1.970 | 5.159 | <0.001 | <0.001 |
| REEP2 | Receptor expression-enhancing protein 2 | -1.234 | -4.412 | <0.001 | 0.001 |
| REG3A | Regenerating islet-derived protein 3-alpha | -1.266 | -4.349 | <0.001 | 0.001 |
| RGS10 | Regulator of G-protein signaling 10 | -1.026 | -3.338 | 0.002 | 0.009 |
| RNASE3 | Eosinophil cationic protein | 1.294 | 4.095 | <0.001 | 0.002 |
| RNPEP | Aminopeptidase B | -1.036 | -5.176 | <0.001 | <0.001 |
| RPL26L1 | 60S ribosomal protein L26-like 1 | 1.149 | 3.589 | 0.001 | 0.006 |
| RPP25 | Ribonuclease P protein subunit p25 | -2.214 | -6.060 | <0.001 | <0.001 |

| | | | | | |
|---|---|---|---|---|---|
| RUFY1 | RUN and FYVE domain-containing protein 1 | 1.297 | 4.570 | <0.001 | 0.001 |
| S100A4 | Protein S100-A4 | -1.272 | -5.992 | <0.001 | <0.001 |
| S100A5 | Protein S100-A5 | -1.422 | -7.845 | <0.001 | <0.001 |
| S100P | Protein S100-P | -1.004 | -6.370 | <0.001 | <0.001 |
| SAA1 | Serum amyloid A-1 protein | 1.775 | 3.580 | 0.001 | 0.006 |
| SCN4B | Sodium channel subunit beta-4 | 1.003 | 4.080 | <0.001 | 0.002 |
| SELENOS | Selenoprotein S | 1.221 | 4.215 | <0.001 | 0.002 |
| SEMA3G | Semaphorin-3G | 1.671 | 6.453 | <0.001 | <0.001 |
| SERPINA10 | Protein Z-dependent protease inhibitor | 1.295 | 3.137 | 0.004 | 0.014 |
| SERPINA11 | Serpin A11 | 1.073 | 3.877 | <0.001 | 0.003 |
| SERPINA5 | Plasma serine protease inhibitor | 2.945 | 3.835 | 0.001 | 0.003 |
| SERPINF2 | Alpha-2-antiplasmin | 2.327 | 5.832 | <0.001 | <0.001 |
| SERPING1 | Plasma protease C1 inhibitor | 1.945 | 3.371 | 0.002 | 0.009 |
| SEZ6L | Seizure 6-like protein | 1.000 | 2.927 | 0.006 | 0.021 |
| SH3BGRL2 | SH3 domain-binding glutamic acid-rich-like protein 2 | -1.347 | -3.445 | 0.002 | 0.007 |
| SHC1 | SHC-transforming protein 1:Src Homology domain | -1.281 | -4.196 | <0.001 | 0.002 |
| SIGLEC12 | Sialic acid-binding Ig-like lectin 12:Ig-like C2-type 2 domain; Isoform short | 1.830 | 3.950 | <0.001 | 0.003 |
| SLC22A16 | Solute carrier family 22 member 16 | 1.432 | 2.900 | 0.007 | 0.022 |
| SLC35G2 | Solute carrier family 35 member G2 | 1.799 | 3.445 | 0.002 | 0.007 |
| SLC5A8 | Sodium-coupled monocarboxylate transporter 1 | 1.063 | 3.754 | 0.001 | 0.004 |
| SLC6A14 | Sodium- and chloride-dependent neutral and basic amino acid transporter B(0+) | 1.271 | 4.011 | <0.001 | 0.002 |
| SNCA | Alpha-synuclein | 1.841 | 3.756 | 0.001 | 0.004 |
| SNCG | Gamma-synuclein | 1.523 | 3.288 | 0.002 | 0.010 |
| SNX3 | Sorting nexin-3 | 1.024 | 3.978 | <0.001 | 0.003 |
| SORD | Sorbitol dehydrogenase | 1.634 | 6.364 | <0.001 | <0.001 |
| SPARCL1 | SPARC-like protein 1 | -1.018 | -4.420 | <0.001 | 0.001 |
| SQSTM1 | Sequestosome-1 | 1.239 | 5.163 | <0.001 | <0.001 |
| SRI | Sorcin | -1.332 | -6.385 | <0.001 | <0.001 |
| STARD10 | START domain-containing protein 10 | 1.040 | 5.398 | <0.001 | <0.001 |
| STK10 | Serine/threonine kinase 10 | -1.195 | -5.302 | <0.001 | <0.001 |
| STK24 | Serine/threonine-protein kinase 24 | 1.698 | 4.202 | <0.001 | 0.002 |
| STMN4 | Stathmin-4 | 1.154 | 2.540 | 0.016 | 0.044 |
| STX17 | Syntaxin-17 | -1.195 | -4.978 | <0.001 | <0.001 |
| STX4 | Syntaxin-4 | -1.196 | -3.924 | <0.001 | 0.003 |
| STX8 | Syntaxin-8 | -1.125 | -3.780 | 0.001 | 0.004 |
| SUMO2 | Small ubiquitin-related modifier 2 | -1.782 | -6.801 | <0.001 | <0.001 |
| SUMO4 | Small ubiquitin-related modifier 4 | -1.007 | -5.066 | <0.001 | <0.001 |
| SURF1 | Surfeit locus protein 1 | -1.634 | -5.620 | <0.001 | <0.001 |
| SYTL4 | Synaptotagmin-like protein 4:Ca2+-dependent membrane-targeting module 1 | -1.619 | -3.627 | 0.001 | 0.005 |
| TAF8 | Transcription initiation factor TFIID subunit 8 | 1.233 | 4.800 | <0.001 | 0.001 |
| TAGLN2 | Transgelin-2 | -1.163 | -4.540 | <0.001 | 0.001 |
| TANK | TRAF family member-associated NF-kappa-B activator | -1.210 | -4.284 | <0.001 | 0.001 |
| TBC1D5 | TBC1 domain family member 5 | 1.209 | 5.200 | <0.001 | <0.001 |
| TBCE | Tubulin-specific chaperone E | -1.195 | -3.120 | 0.004 | 0.014 |
| TEX29 | Testis-expressed sequence 29 protein | -1.529 | -4.778 | <0.001 | 0.001 |
| THBS4 | Thrombospondin-4 | 1.139 | 5.764 | <0.001 | <0.001 |
| THRB | Thyroid hormone receptor beta | -1.468 | -6.319 | <0.001 | <0.001 |
| TIGIT | T-cell immunoreceptor with Ig and ITIM domains | 1.012 | 4.985 | <0.001 | <0.001 |
| TK2 | Thymidine kinase 2; mitochondrial | -1.234 | -4.250 | <0.001 | 0.002 |
| TKFC | Dihydroxyacetone kinase | 1.328 | 4.390 | <0.001 | 0.001 |
| TMEM225B | Transmembrane protein 225B | -1.129 | -5.597 | <0.001 | 0.000 |
| TMEM59L | Transmembrane protein 59-like | -1.309 | -5.963 | <0.001 | 0.000 |
| TMPRSS6 | Transmembrane protease serine 6 | 1.304 | 3.760 | 0.001 | 0.004 |
| TNFRSF14 | Tumor necrosis factor receptor superfamily member 14 | 1.315 | 4.797 | <0.001 | 0.001 |
| TP53I11 | Tumor protein p53-inducible protein 11 | 1.429 | 4.962 | <0.001 | <0.001 |
| TPD52L1 | Tumor protein D53 | 1.087 | 4.723 | <0.001 | 0.001 |

| | | | | | |
|---|---|---|---|---|---|
| TPD52L2 | Tumor protein D54 | 1.549 | 4.075 | <0.001 | 0.002 |
| TPM4 | Tropomyosin alpha-4 chain | -1.347 | -3.128 | 0.004 | 0.014 |
| TPST1 | Protein-tyrosine sulfotransferase 1 | 1.317 | 3.310 | 0.002 | 0.010 |
| TPT1 | Translationally-controlled tumor protein | -1.712 | -6.287 | <0.001 | <0.001 |
| TRABD2A | Metalloprotease TIKI1 | -1.044 | -3.175 | 0.003 | 0.013 |
| TRAPPC2 | Trafficking protein particle complex subunit 2 | -1.214 | -3.637 | 0.001 | 0.005 |
| TRMT112 | TRM112-like protein | 1.129 | 4.258 | <0.001 | 0.001 |
| TRMT6 | tRNA (adenine(58)-N(1))-methyltransferase non-catalytic subunit TRM6 | 1.379 | 4.982 | <0.001 | <0.001 |
| TRMT61B | tRNA (adenine(58)-N(1))-methyltransferase; mitochondrial | -1.027 | -3.378 | 0.002 | 0.009 |
| TSG101 | Tumor susceptibility gene 101 protein | -1.042 | -3.137 | 0.004 | 0.014 |
| TXNIP | Thioredoxin-interacting protein | 1.074 | 4.537 | <0.001 | 0.001 |
| TXNL1 | Thioredoxin-like protein 1 | -1.035 | -6.620 | <0.001 | <0.001 |
| TYMSOS | TYMS opposite strand protein | 1.366 | 4.816 | <0.001 | 0.001 |
| UBASH3B | Ubiquitin-associated and SH3 domain-containing protein B | 1.995 | 3.623 | 0.001 | 0.005 |
| UBE2D3 | Ubiquitin-conjugating enzyme E2 D3 | 1.572 | 4.278 | <0.001 | 0.001 |
| UBE2N\|UBE2V1 | UBE2N (Ubc13)/Uev1a Complex | -1.083 | -4.149 | <0.001 | 0.002 |
| USP14 | Ubiquitin carboxyl-terminal hydrolase 14 | 2.036 | 4.030 | <0.001 | 0.002 |
| USP5 | Ubiquitin carboxyl-terminal hydrolase 5 | -1.472 | -5.815 | <0.001 | 0.000 |
| USP8 | Ubiquitin carboxyl-terminal hydrolase 8 | -1.070 | -3.432 | 0.002 | 0.008 |
| VAT1 | Synaptic vesicle membrane protein VAT-1 homolog | 2.020 | 5.453 | <0.001 | <0.001 |
| VCX | Variable charge X-linked protein 1 | 1.713 | 4.198 | <0.001 | 0.002 |
| VEGFC | Vascular endothelial growth factor C | -1.005 | -4.650 | <0.001 | 0.001 |
| VTI1B | Vesicle transport through interaction with t-SNAREs homolog 1B | -1.195 | -3.496 | 0.001 | 0.007 |
| VWC2 | Brorin | 1.372 | 4.428 | <0.001 | 0.001 |
| WDR18 | WD repeat-containing protein 18 | 1.281 | 3.109 | 0.004 | 0.015 |
| WNK3 | Serine/threonine-protein kinase WNK3 | 1.413 | 3.287 | 0.002 | 0.010 |
| WNT10A | Protein Wnt-10a | 2.148 | 4.677 | <0.001 | 0.001 |
| WNT3A | Protein Wnt-3a | 1.834 | 4.090 | <0.001 | 0.002 |
| YWHAG | 14-3-3 protein gamma | -1.123 | -5.061 | <0.001 | <0.001 |
| ZC3H8 | Zinc finger CCCH domain-containing protein 8 | 1.178 | 4.735 | <0.001 | 0.001 |
| ZFAND2B | AN1-type zinc finger protein 2B | 1.020 | 4.039 | <0.001 | 0.002 |
| ZNF263 | Zinc finger protein 263 | 1.147 | 4.540 | <0.001 | 0.001 |

**Supplementary Table 5. Proteins specific to the recovery phase of Guillain-Barré syndrome (*n* = 32).** Phase-specific proteins were defined by consistent directionality of differential abundance across relevant contrasts. $Log_2$ fold-change ($log_2$ FC) indicates the magnitude of relative protein abundance. For both comparisons, positive values denote higher protein abundance in the recovery phase compared to healthy controls or the acute phase of Guillain-Barré syndrome (GBS), respectively. *P*-values were calculated using empirical Bayes moderated statistics and adjusted for multiple testing using the Benjamini-Hochberg false discovery rate (FDR) method. Proteins are listed in alphabetical order by gene symbol. The table includes the 10 proteins already shown in Table 3 of the main manuscript.

| Entrez Gene Symbol | Target Full Name | Recovery GBS vs controls | | Recovery GBS vs acute GBS | |
|---|---|---|---|---|---|
| | | $Log_2$ FC | FDR | $Log_2$ FC | FDR |
| APOC1 | Apolipoprotein C-I | 4.588 | <0.001 | 1.884 | <0.001 |
| APOA4 | Apolipoprotein A-IV | 3.211 | <0.001 | 2.021 | <0.001 |
| APOF | Apolipoprotein F | 3.401 | <0.001 | 1.804 | 0.001 |
| SERPINA5 | Plasma serine protease inhibitor | 2.945 | 0.003 | 1.564 | 0.027 |
| NCF2 | Neutrophil cytosol factor 2 | -2.774 | <0.001 | -1.589 | 0.006 |
| LDHA | L-lactate dehydrogenase A chain | 2.967 | <0.001 | 1.333 | 0.001 |
| PGD | 6-phosphogluconate dehydrogenase; decarboxylating | 2.618 | <0.001 | 1.520 | <0.001 |
| PA2G4 | Proliferation-associated protein 2G4 | 2.609 | 0.001 | 1.375 | <0.001 |
| CHGA | Chromogranin-A | 2.544 | 0.005 | 1.375 | 0.017 |
| PENK | Proenkephalin-A | 2.709 | <0.001 | 1.147 | 0.004 |
| PSMA7 | Proteasome subunit alpha type-7 | -2.289 | <0.001 | -1.253 | 0.001 |
| CRIP1 | Cysteine-rich protein 1 | 2.345 | <0.001 | 1.191 | <0.001 |
| ANXA7 | Annexin A7 | -2.187 | <0.001 | -1.282 | 0.001 |
| PRCP | Lysosomal Pro-X carboxypeptidase | 2.375 | <0.001 | 1.083 | 0.001 |
| USP14 | Ubiquitin carboxyl-terminal hydrolase 14 | 2.036 | 0.002 | 1.325 | 0.001 |
| CRABP2 | Cellular retinoic acid-binding protein 2 | 1.833 | <0.001 | 1.407 | <0.001 |
| HMGCS1 | Hydroxymethylglutaryl-CoA synthase; cytoplasmic | 2.136 | <0.001 | 1.047 | 0.001 |
| UBASH3B | Ubiquitin-associated and SH3 domain-containing protein B | 1.995 | 0.005 | 1.032 | 0.004 |
| SUMO2 | Small ubiquitin-related modifier 2 | -1.782 | <0.001 | -1.030 | 0.005 |
| CLINT1 | Clathrin interactor 1 | -1.607 | 0.005 | -1.147 | 0.033 |
| ANXA11 | Annexin A11 | -1.632 | <0.001 | -1.120 | 0.003 |
| COL11A2 | Collagen alpha-2(XI) chain | 1.416 | 0.004 | 1.304 | <0.001 |
| ADH1C | Alcohol dehydrogenase 1C | 1.590 | <0.001 | 1.037 | <0.001 |
| THBS4 | Thrombospondin-4 | 1.139 | <0.001 | 1.425 | <0.001 |
| ANTXR2 | Anthrax toxin receptor 2 | 1.358 | <0.001 | 1.181 | <0.001 |
| SH3BGRL2 | SH3 domain-binding glutamic acid-rich-like protein 2 | -1.347 | 0.007 | -1.165 | 0.027 |
| CHGA | Chromogranin-A | 1.356 | 0.042 | 1.127 | 0.011 |
| TPM4 | Tropomyosin alpha-4 chain | -1.347 | 0.014 | -1.132 | 0.030 |
| H3C1 | Histone H3.1 | -1.169 | 0.007 | -1.180 | <0.001 |
| TAGLN2 | Transgelin-2 | -1.163 | 0.001 | -1.085 | 0.005 |
| MANF | Mesencephalic astrocyte-derived neurotrophic factor | -1.023 | 0.024 | -1.138 | 0.007 |
| EHD3 | EH domain-containing protein 3 | -1.010 | 0.022 | -1.086 | 0.021 |

**Supplementary Table 6: Differential abundance of previously reported peripheral neuropathy biomarkers in the acute phase of Guillain-Barré syndrome compared to the recovery phase and healthy controls**. This table shows the relative abundance of 26 curated proteins which have been previously reported as possible biomarkers in neuropathies. $Log_2$ fold-change ($log_2FC$) indicates the magnitude of relative protein abundance. For both comparative columns, a positive value denotes higher protein abundance in the acute phase. *P*-values were calculated using empirical Bayes moderated statistics and adjusted for multiple testing using the Benjamini-Hochberg false discovery rate (FDR) method. Proteins are listed alphabetically by gene symbol.

| Entrez Gene Symbol | Target Full Name | Acute GBS vs Recovery GBS | | Acute GBS vs Healthy Controls | |
|---|---|---|---|---|---|
| | | $Log_2FC$ | FDR | $Log_2FC$ | FDR |
| BDNF | Brain-derived neurotrophic factor | -0.019 | 0.953 | -0.356 | 0.078 |
| CNTN1 | Contactin-1 | -0.332 | 0.018 | -0.642 | 0.004 |
| CNTN2 | Contactin-2 | -0.393 | <0.001 | -0.53 | 0.003 |
| CNTN3 | Contactin-3 | -0.177 | 0.123 | -0.256 | 0.179 |
| CNTN4 | Contactin-4 | -0.279 | 0.004 | -0.509 | 0.004 |
| CNTN5 | Contactin-5 | -0.002 | 0.988 | 0.042 | 0.699 |
| CNTN6 | Contactin-6 | -0.394 | 0.006 | -0.727 | 0.005 |
| DCTN2 | Dynactin 2 | 0.042 | 0.587 | -0.081 | 0.540 |
| ENO2 | Enolase 2 | 0.676 | 0.024 | 0.648 | 0.095 |
| GFAP | Glial Fibrillary Acidic Protein | -0.039 | 0.903 | 0.183 | 0.524 |
| GLRX | Glutaredoxin | -0.045 | 0.678 | 0.39 | 0.012 |
| IRAK4 | Interleukin-1 receptor-associated kinase 4 | 0.117 | 0.168 | 0.056 | 0.785 |
| MAPT | Microtubule associated protein tau | 0.09 | 0.510 | 0.049 | 0.809 |
| MMP9 | Matrix metalloproteinase-9 | 0.717 | 0.008 | 0.848 | 0.013 |
| NCAM1 | Neural Cell Adhesion Molecule 1 | -0.187 | 0.097 | -0.369 | 0.062 |
| NCAM2 | Neural Cell Adhesion Molecule 2 | -0.365 | 0.003 | -0.682 | 0.004 |
| NEFH | Neurofilament Heavy Chain | -0.018 | 0.961 | 0.587 | 0.044 |
| NEFL | Neurofilament Light Chain | 0.245 | 0.040 | 0.9 | 0.003 |
| NGF | Nerve Growth Factor | -0.018 | 0.839 | -0.292 | 0.180 |
| NGFR | Nerve Growth Factor Receptor | -0.339 | 0.081 | 0.009 | 0.979 |
| NT5C3A | Cytosolic 5'-nucleotidase 3 | 0.194 | 0.201 | 0.39 | 0.124 |
| PRX | Periaxin | 0.09 | 0.355 | 0.149 | 0.248 |
| S100A9 | Protein S100-A9 | 0.134 | 0.502 | 0.474 | 0.023 |
| SPP1 | Osteopontin | 0.173 | 0.003 | 0.419 | 0.014 |
| SUGT1 | Suppressor of G2 allele of skp1 | 0.513 | 0.059 | -0.21 | 0.670 |
| TMPRSS5 | Transmembrane Serine Protease 5 | -0.121 | 0.397 | 0.763 | 0.004 |

**Supplementary Table 7.** Significantly enriched biological pathways between the acute and recovery phases of Guillain-Barré syndrome. Gene-set enrichment analysis was performed using *Reactome* and *Gene Ontology Biological Process* (GOBP) terms. The normalized enrichment score (NES) reflects the degree to which a pathway is overrepresented. Positive values denote enrichment in the acute phase, whereas negative values denote enrichment in the recovery phase. Gene set size indicates the number of measured proteins mapped to each respective pathway. *P*-values were adjusted for multiple testing using the Benjamini-Hochberg false discovery rate method. For conciseness, only the top 50 pathways, sorted by highest absolute NES, are presented.

| Biological Pathway | Adjusted P-Value | Normalized Enrichment Score | Gene set size |
|---|---|---|---|
| Neuron Projection Guidance | < 0.001 | -2.58 | 149 |
| Axon Development | < 0.001 | -2.45 | 274 |
| Collagen Fibril Organization | < 0.001 | -2.43 | 36 |
| Neuron Recognition | < 0.001 | -2.38 | 33 |
| Assembly Of Collagen Fibrils And Other Multimeric Structures | < 0.001 | -2.30 | 30 |
| Collagen Chain Trimerization | < 0.001 | -2.30 | 20 |
| External Encapsulating Structure Organization | < 0.001 | -2.29 | 177 |
| Collagen Formation | < 0.001 | -2.22 | 47 |
| Collagen Biosynthesis And Modifying Enzymes | < 0.001 | -2.21 | 35 |
| Cell Morphogenesis Involved In Neuron Differentiation | < 0.001 | -2.20 | 287 |
| Cell Projection Morphogenesis | < 0.001 | -2.20 | 326 |
| Ecm Proteoglycans | < 0.001 | -2.20 | 46 |
| Artery Development | < 0.001 | -2.19 | 56 |
| Aorta Development | 0.002 | -2.19 | 31 |
| Regulation Of Axonogenesis | < 0.001 | -2.16 | 83 |
| Signaling By The B Cell Receptor Bcr | < 0.001 | 2.15 | 62 |
| Cell Cell Adhesion Via Plasma Membrane Adhesion Molecules | < 0.001 | -2.14 | 149 |
| Regulation Of Commissural Axon Pathfinding By Slit And Robo | 0.002 | -2.14 | 10 |
| Positive Regulation Of Synapse Assembly | < 0.001 | -2.12 | 53 |
| Uch Proteinases | < 0.001 | 2.12 | 37 |
| Epithelial Mesenchymal Transition | < 0.001 | -2.11 | 140 |
| Mrna Metabolic Process | < 0.001 | 2.11 | 233 |
| Diseases Of Glycosylation | < 0.001 | -2.11 | 70 |
| Downstream Signaling Events Of B Cell Receptor Bcr | < 0.001 | 2.09 | 40 |
| Global Genome Nucleotide Excision Repair Gg Ner | < 0.001 | 2.08 | 32 |
| Runx1 Regulates Transcription Of Genes Involved In Differentiation Of Hscs | < 0.001 | 2.08 | 34 |
| Gene And Protein Expression By Jak Stat Signaling After Interleukin 12 Stimulation | < 0.001 | 2.08 | 29 |
| Pulmonary Valve Development | 0.003 | -2.07 | 16 |
| Regulation Of Ras By Gaps | < 0.001 | 2.07 | 28 |
| Negative Chemotaxis | 0.003 | -2.06 | 39 |
| Positive Regulation Of Cell Junction Assembly | < 0.001 | -2.06 | 79 |
| Mrna Processing | < 0.001 | 2.05 | 152 |
| Keratan Sulfate Keratin Metabolism | 0.005 | -2.05 | 24 |
| Hedgehog Off State | < 0.001 | 2.05 | 35 |
| Prostate Gland Morphogenesis | 0.009 | -2.05 | 18 |
| Positive Regulation Of Proteolysis Involved In Protein Catabolic Process | < 0.001 | 2.05 | 61 |
| Pulmonary Valve Morphogenesis | 0.005 | -2.04 | 13 |
| Neuron Projection Regeneration | 0.002 | -2.04 | 43 |
| Embryonic Organ Morphogenesis | < 0.001 | -2.04 | 106 |
| Rna Splicing Via Transesterification Reactions | < 0.001 | 2.03 | 107 |
| Homophilic Cell Adhesion Via Plasma Membrane Adhesion Molecules | < 0.001 | -2.03 | 80 |
| Cardiac Septum Development | < 0.001 | -2.02 | 59 |
| Eye Morphogenesis | 0.001 | -2.02 | 73 |
| Artery Morphogenesis | 0.003 | -2.02 | 40 |
| Regulation Of Runx3 Expression And Activity | 0.002 | 2.02 | 29 |

| | | | |
|---|---|---|---|
| Hiv Infection | < 0.001 | 2.02 | 93 |
| Regulation Of Mrna Processing | 0.002 | 2.02 | 46 |
| Collagen Degradation | 0.003 | -2.02 | 35 |
| Response To Bmp | < 0.001 | -2.01 | 91 |
| Tgf Beta Receptor Signaling In Emt Epithelial To Mesenchymal Transition | 0.002 | 2.01 | 11 |

**Supplementary Table 8.** Significantly enriched biological pathways between the acute phase of Guillain-Barré syndrome and healthy controls. Gene-set enrichment analysis was performed using *Reactome* and *Gene Ontology Biological Process* (GOBP) terms. The normalized enrichment score reflects the degree to which a pathway is overrepresented. Positive values denote enrichment in the acute phase of Guillain-Barré syndrome, whereas negative values denote enrichment in healthy controls. Gene set size indicates the number of measured proteins mapped to each respective pathway. *P*-values were adjusted for multiple testing using the Benjamini-Hochberg false discovery rate method. For conciseness, only the top 50 pathways, sorted by highest absolute NES, are presented.

| Biological Pathway | Adjusted P-Value | Normalized Enrichment Score | Gene set size |
|---|---|---|---|
| Neuron Recognition | <0.001 | -2.62 | 33 |
| Positive Regulation Of Synapse Assembly | <0.001 | -2.41 | 53 |
| Neuron Projection Guidance | <0.001 | -2.35 | 149 |
| Axonal Fasciculation | 0.008 | -2.33 | 14 |
| Hedgehog Signaling | 0.007 | -2.25 | 23 |
| Sarcomere Organization | 0.001 | 2.24 | 25 |
| Synapse Assembly | <0.001 | -2.22 | 162 |
| Axon Development | <0.001 | -2.20 | 274 |
| Synaptic Membrane Adhesion | 0.005 | -2.19 | 26 |
| Ephrin Receptor Signaling Pathway | 0.003 | -2.15 | 41 |
| Positive Regulation Of Axonogenesis | 0.019 | -2.05 | 44 |
| Eph Ephrin Mediated Repulsion Of Cells | 0.047 | -2.03 | 38 |
| Regulation Of Axonogenesis | 0.002 | -2.01 | 83 |
| Cell Projection Morphogenesis | <0.001 | -2.00 | 326 |
| Positive Regulation Of Nervous System Development | <0.001 | -1.99 | 159 |
| Regulation Of Notch Signaling Pathway | 0.015 | -1.99 | 53 |
| Positive Regulation Of Cell Junction Assembly | 0.003 | -1.98 | 79 |
| Morphogenesis Of An Epithelial Sheet | 0.041 | -1.98 | 30 |
| Cell Cell Adhesion Via Plasma Membrane Adhesion Molecules | <0.001 | -1.97 | 149 |
| Cell Morphogenesis Involved In Neuron Differentiation | <0.001 | -1.97 | 287 |
| Myofibril Assembly | 0.016 | 1.95 | 37 |
| Mrna Splicing | 0.015 | 1.95 | 76 |
| Regulation Of Synapse Assembly | 0.002 | -1.94 | 113 |
| Processing Of Capped Intron Containing Pre Mrna | 0.010 | 1.93 | 89 |
| Integrated Stress Response Signaling | 0.041 | 1.89 | 23 |
| Regulation Of Cytoplasmic Pattern Recognition Receptor Signaling Pathway | 0.034 | 1.85 | 47 |
| Cell Recognition | 0.009 | -1.81 | 101 |
| Membraneless Organelle Assembly | 0.006 | 1.79 | 154 |
| Protein Rna Complex Organization | 0.028 | 1.78 | 77 |
| Regulation Of Nervous System Development | 0.000 | -1.78 | 237 |
| Defense Response To Gram Positive Bacterium | 0.028 | 1.77 | 86 |
| Mrna Processing | 0.010 | 1.77 | 152 |
| Rna Splicing Via Transesterification Reactions | 0.026 | 1.76 | 107 |
| Synapse Organization | <0.001 | -1.76 | 298 |
| Cell Junction Assembly | 0.001 | -1.74 | 258 |
| Monosaccharide Metabolic Process | 0.026 | 1.73 | 137 |
| Positive Regulation Of Neuron Projection Development | 0.016 | -1.72 | 93 |
| Central Nervous System Neuron Differentiation | 0.027 | -1.72 | 94 |
| Epithelial Mesenchymal Transition | 0.007 | -1.71 | 140 |
| Glucose Metabolic Process | 0.041 | 1.71 | 103 |
| Positive Regulation Of Cell Projection Organization | 0.006 | -1.70 | 177 |
| Mrna Metabolic Process | 0.006 | 1.69 | 233 |
| Ribonucleoprotein Complex Biogenesis | 0.041 | 1.68 | 124 |
| Positive Regulation Of Neurogenesis | 0.034 | -1.68 | 118 |
| Regulation Of Synapse Structure Or Activity | 0.005 | -1.68 | 182 |
| Rna Processing | 0.003 | 1.67 | 269 |
| Rna Splicing | 0.029 | 1.66 | 147 |

| | | | |
|---|---|---|---|
| Regulation Of Cell Junction Assembly | 0.029 | -1.64 | 164 |
| Regulation Of Neurogenesis | 0.015 | -1.62 | 182 |
| Cell Morphogenesis | <0.001 | -1.61 | 456 |

**Supplementary Table 9.** Significantly enriched biological pathways between the recovery phase of Guillain-Barré syndrome and healthy controls. Gene-set enrichment analysis was performed using *Reactome* and *Gene Ontology Biological Process* (GOBP) terms. The normalized enrichment score reflects the degree to which a pathway is overrepresented. Positive values denote enrichment in the recovery phase of Guillain-Barré syndrome, whereas negative values denote enrichment in healthy controls. Gene set size indicates the number of measured proteins mapped to each respective pathway. *P*-values were adjusted for multiple testing using the Benjamini-Hochberg false discovery rate method. For conciseness, only the top 50 pathways, sorted by highest absolute NES, are presented.

| Biological Pathway | Adjusted P-Value | Normalized Enrichment Score | Gene set size |
|---|---|---|---|
| Regulation Of Ras By Gaps | <0.001 | -2.58 | 28 |
| Dectin 1 Mediated Noncanonical Nf Kb Signaling | <0.001 | -2.55 | 29 |
| Degradation Of Dvl | <0.001 | -2.55 | 26 |
| Proteasome Assembly | <0.001 | -2.52 | 31 |
| Hh Mutants Abrogate Ligand Secretion | <0.001 | -2.51 | 26 |
| Uch Proteinases | <0.001 | -2.50 | 37 |
| Activation Of Nf Kappab In B Cells | <0.001 | -2.50 | 33 |
| The Role Of Gtse1 In G2 M Progression After G2 Checkpoint | <0.001 | -2.49 | 30 |
| Regulation Of Apoptosis | <0.001 | -2.49 | 22 |
| Downstream Signaling Events Of B Cell Receptor Bcr | <0.001 | -2.48 | 40 |
| Scf Beta Trcp Mediated Degradation Of Emi1 | <0.001 | -2.48 | 24 |
| Fbxl7 Down Regulates Aurka During Mitotic Entry And In Early Mitosis | <0.001 | -2.47 | 25 |
| Ubiquitin Dependent Degradation Of Cyclin D | <0.001 | -2.46 | 23 |
| Degradation Of Gli1 By The Proteasome | <0.001 | -2.46 | 28 |
| Stabilization Of P53 | <0.001 | -2.46 | 26 |
| Auf1 Hnrnp D0 Binds And Destabilizes Mrna | <0.001 | -2.45 | 27 |
| Orc1 Removal From Chromatin | <0.001 | -2.45 | 30 |
| Regulation Of Runx3 Expression And Activity | <0.001 | -2.44 | 29 |
| Asymmetric Localization Of Pcp Proteins | <0.001 | -2.43 | 32 |
| Negative Regulation Of Notch4 Signaling | <0.001 | -2.43 | 27 |
| Degradation Of Beta Catenin By The Destruction Complex | <0.001 | -2.43 | 34 |
| Fceri Mediated Nf Kb Activation | <0.001 | -2.42 | 39 |
| Downstream Tcr Signaling | <0.001 | -2.40 | 49 |
| Defective Cftr Causes Cystic Fibrosis | <0.001 | -2.39 | 27 |
| Cross Presentation Of Soluble Exogenous Antigens Endosomes | <0.001 | -2.38 | 27 |
| Vif Mediated Degradation Of Apobec3G | <0.001 | -2.37 | 23 |
| G1 S Dna Damage Checkpoints | <0.001 | -2.35 | 34 |
| Degradation Of Cry And Per Proteins | <0.001 | -2.35 | 25 |
| Antigen Processing Ub Atp Independent Proteasomal Degradation | <0.001 | -2.34 | 17 |
| Hedgehog Off State | <0.001 | -2.33 | 35 |
| Abc Transporter Disorders | <0.001 | -2.31 | 30 |
| Runx1 Regulates Transcription Of Genes Involved In Differentiation Of Hscs | <0.001 | -2.29 | 34 |
| Cdk Mediated Phosphorylation And Removal Of Cdc6 | <0.001 | -2.28 | 32 |
| Signaling By Notch4 | <0.001 | -2.28 | 40 |
| Scf Skp2 Mediated Degradation Of P27 P21 | <0.001 | -2.27 | 33 |
| Mitotic G2 G2 M Phases | <0.001 | -2.27 | 60 |
| Somitogenesis Somitogenesis | <0.001 | -2.26 | 24 |
| Pcp Ce Pathway | <0.001 | -2.24 | 48 |
| Abc Family Proteins Mediated Transport | <0.001 | -2.23 | 31 |
| Regulation Of Mrna Stability By Proteins That Bind Au Rich Elements | <0.001 | -2.22 | 45 |
| Hedgehog Ligand Biogenesis | 0.001 | -2.21 | 30 |
| Regulation Of Pten Stability And Activity | <0.001 | -2.21 | 35 |
| Positive Regulation Of Platelet Activation | 0.028 | -2.21 | 12 |
| Activation Of Apc C And Apc C Cdc20 Mediated Degradation Of Mitotic Proteins | <0.001 | -2.20 | 34 |
| Switching Of Origins To A Post Replicative State | <0.001 | -2.20 | 38 |
| Clec7A Dectin 1 Signaling | <0.001 | -2.17 | 53 |
| Metabolism Of Polyamines | 0.002 | -2.16 | 26 |

| | | | |
|---|---|---|---|
| Hedgehog On State | <0.001 | -2.16 | 39 |
| Regulation Of Runx2 Expression And Activity | 0.001 | -2.15 | 35 |
| Signaling By The B Cell Receptor Bcr | <0.001 | -2.15 | 62 |

**Supplementary Table 10. Baseline demographic characteristics of the Serum Amyloid A (SAA) validation cohort across all diagnostic groups.** Data are presented as mean ± standard deviation, and absolute frequency (percentage). Data regarding sex were unavailable for the acute disseminated encephalomyelitis (ADEM) cohort.

| Characteristics | Guillain-Barré syndrome | Healthy controls | Myasthenia gravis | ADEM |
|---|---|---|---|---|
| n | 22 | 22 | 22 | 14 |
| Age, mean ± SD | 56.2 ± 18.1 | 45.9 ± 17.0 | 57.6 ± 20.7 | 3.79 ± 2.62 |
| Sex, n (%) | | | | |
| Female | 10 (45.5) | 12 (54.5) | 10 (45.5) | NA |
| Male | 12 (54.5) | 10 (45.5) | 12 (54.5) | NA |

**Supplementary Table 11. Demographic and clinical characteristics of the high-sensitivity cardiac troponin T (hs-cTnT) validation cohort.** Data are presented as mean ± standard deviation, and absolute frequency (percentage).

| Characteristics | Guillain-Barré syndrome | Healthy controls | Myasthenia gravis | Multiple Sclerosis |
|---|---|---|---|---|
| n | 51 | 26 | 19 | 20 |
| Age, mean ± SD | 55.5 ± 18.3 | 48.9 ± 16.8 | 56.8 ± 20.7 | 45.2 ± 12.8 |
| Sex, n (%) | | | | |
| Female | 18 (34.7) | 14 (53.8) | 9 (47.4) | 15 (75) |
| Male | 33 (65.3) | 12 (46.2) | 10 (52.6) | 5 (25) |
