## Supplementary material for "Longitudinal proteomics defines stage-specific molecular signatures in Guillain-Barré syndrome": Mendelian Randomisation Full Methods and Results

### MENDELIAN RANDOMIZATION REPORT

#### Proteomic profiling of Guillain-Barré syndrome using aptamer-based technology: identifying and validating potential biomarkers

### eMETHODS

This study follows the guidelines for strengthening the reporting of MR studies (STROBE-MR).[1]

#### Data sources

We selected datasets that were either entirely or predominantly composed of individuals of European ancestry to minimize the risk of population stratification. As exposures, we used genome-wide association studies (GWAS) of plasma protein levels measured by the SomaScan platform, obtained from the publicly available deCODE genetics repository.[2] Of the 257 proteins, 188 have a GWAS summary in this repository, and only 82 showed cis-protein quantitative trait loci (cis-pQTLs). As an outcome, we used the GWAS for acute infective polyneuritis/Guillain-Barré syndrome in the UK Biobank European ancestry cohort (174 cases and 420,299 non-cases) from Pan-UKB repository.

#### Statistical analysis

##### Univariate MR analysis

For the MR analysis, we selected genome-wide significant variants ($p < 5 \times 10^{-8}$) that were clumped for linkage disequilibrium (LD) at $r^2 < 0.1$ within a ±500 kb window of each protein locus (*cis*-pQTL) as instrumental variables (IV). Variants close to the encoding gene are expected to have a greater biological effect on the protein.[3] We used the same surrounding region for further colocalization analysis (see below). When possible, we used proxy alleles if IV-selected exposures are not present in the outcome. After harmonizing the effect sizes for SNPs in exposures and outcomes to ensure the use of the same reference allele, we estimated the F-statistic ($F_{st}$) for each variant to measure the strength of the genetic instruments, discarding those with $F_{st} < 10$. Then, we performed a two-sample MR analysis using the inverse-variance weighted (IVW) method as the primary analysis. In the absence of horizontal pleiotropy, the IVW can provide an accurate estimate of causality.[4] We used fixedeffect IVW in the absence of heterogeneity or when clumping yielded 2-3 IVs, and random-effect IVW in the remaining situations.[5] When only one IV resulted from clumping, we used Wald's ratio method.

Additionally, we used robust MR methods, including weighted median (WM) and MR-Egger regression. WM provides a robust estimate of causality even in the presence of up to 50% invalid IV[6], and the MR-Egger method provides an accurate estimate of true causality in the presence of directional pleiotropy.[7] We also performed the MR using the robust adjusted profile score (MR-RAPS)[8] method. Using MR-RAPS ensures robustness against pleiotropy and weak instruments. Together, they provide complementary layers of protection against distinct sources of bias, enhancing the credibility of causal inference in our study. Results were considered consistent if all the methods indicated the same direction in the beta effect.

***Sensitivity analysis***

Finally, we performed sensitivity analyses to assess the robustness of the results. We evaluated heterogeneity among individual genetic variants using Cochran's Q test[9] and $I^2$[10], and assessed evidence of directional pleiotropy using the MR-Egger regression test for the intercept.[7] We calculated the weighted $I^2_{GX}$ statistic to provide an indicator of the expected relative bias of the MR-Egger causal estimate,[11] i.e., the actual variance of the SNP-exposure association. $I^2_{GX} < 0.9$ indicates a violation of the NO Measurement Error (NOME) assumption, which implies that the variance of the association between IVs and the exposure is not negligible. To identify and remove unbalanced pleiotropic IVs identified as outliers, we performed the MR-Pleiotropy Residual Sum and Outlier (MR-PRESSO) test[12]. Finally, we performed leave-one-out plots to evaluate whether individual IVs drove the overall effect estimate.

We considered true causal associations when we did not detect evidence of severe heterogeneity ($I^2 < 50\%$) and/or directional pleiotropy (*p*-value of the MR Egger intercept >

0.05) in consistent results, and IVW remained significant after applying the false discovery rate (FDR) correction for multiple testing.

##### *Colocalization*

Colocalization can provide complementary evidence supporting a shared mechanism linking exposure and outcome. To assess whether the same genetic variant was associated with both exposure and outcome, which can reinforce the causality association, we used the coloc R package to perform a colocalization test for two genetic traits.[13] Test follows a Bayesian approach to calculate the posterior probability (PP) that a genetic variant is both a protein quantitative trait locus (pQTL) and associated with ICH. We considered a 500 kb region surrounding each locus on both sides, and a locus was considered highly colocalized when the PP value was greater than 0.8. We used the Sum of Single Effects (SuSiE) framework to relax the single-causal variant assumption and fine-map multiple causal variants when H3 or H4 exceeded 0.8.

#### *Software*

We performed all analyses using R version 4.5.2[14] and RStudio version 2025.09.2-418.[15] We used the Two-sample Mendelian Randomization (TwoSampleMR v0.6.7)[16], MR_PRESSO[12], and MR-RAPS[8] R packages for MR analyses. We also used the coloc package version 5.2.3[13] for colocalization analysis.

#### *Data Availability*

The data supporting the findings of this study are available from the corresponding author upon reasonable request.

### eRESULTS

**Table 1**: Characteristics of the data sources included in the Mendelian randomization analysis

| Trait | ID | Cases | Controls | Sample size | Number of variants | Author | PMID[a] |
|---|---|---|---|---|---|---|---|
| **exposure** | | | | | | | |
| Short/branched chain specific acyl-CoA dehydrogenase; mitochondrial | ACADSB | | | 35,739.52 | 33,450,168 | Eldjarn GH (2023) | 37794188 |
| Acyl-CoA-binding domain-containing protein 6 | ACBD6 | | | 35,739.52 | 33,450,168 | Eldjarn GH (2023) | 37794188 |
| Tartrate-resistant acid phosphatase type 5 | ACP5 | | | 35,739.52 | 33,450,167 | Eldjarn GH (2023) | 37794188 |
| Aminoacylase-1 | ACY1 | | | 35,739.52 | 33,450,167 | Eldjarn GH (2023) | 37794188 |
| Advanced glycosylation end product-specific receptor; soluble | AGER | | | 35,779.07 | 33,450,173 | Eldjarn GH (2023) | 37794188 |
| Adenylate kinase isoenzyme 1 | AK1 | | | 35,739.52 | 33,450,167 | Eldjarn GH (2023) | 37794188 |
| Retinal dehydrogenase 1 | ALDH1A1 | | | 35,739.52 | 33,450,168 | Eldjarn GH (2023) | 37794188 |
| Apolipoprotein A-I | APOA1 | | | 35,739.52 | 33,450,167 | Eldjarn GH (2023) | 37794188 |
| Apolipoprotein B | APOB | | | 35,779.07 | 33,450,171 | Eldjarn GH (2023) | 37794188 |
| Apolipoprotein C-I | APOC1 | | | 35,739.52 | 33,450,167 | Eldjarn GH (2023) | 37794188 |
| Apolipoprotein E | APOE | | | 35,739.52 | 33,450,167 | Eldjarn GH (2023) | 37794188 |
| Beta-arrestin-1 | ARRB1 | | | 35,739.52 | 33,450,168 | Eldjarn GH (2023) | 37794188 |
| Argininosuccinate lyase | ASL | | | 35,739.52 | 33,450,167 | Eldjarn GH (2023) | 37794188 |
| Cyclic AMP-dependent transcription factor ATF-6 beta | ATF6B | | | 35,739.52 | 33,450,167 | Eldjarn GH (2023) | 37794188 |
| BPI fold-containing family B member 1 | BPIFB1 | | | 35,739.52 | 33,450,166 | Eldjarn GH (2023) | 37794188 |
| Complement C1r subcomponent | C1R | | | 35,739.52 | 33,450,168 | Eldjarn GH (2023) | 37794188 |
| Complement C4 | C4A_C4B | | | 35,739.52 | 33,450,164 | Eldjarn GH (2023) | 37794188 |
| Carbonic anhydrase-related protein 11 | CA11 | | | 35,739.52 | 33,450,168 | Eldjarn GH (2023) | 37794188 |
| WNT1-inducible-signaling pathway protein 1 | CCN4 | | | 35,739.52 | 33,450,164 | Eldjarn GH (2023) | 37794188 |
| Complement factor H-related protein 4 | CFHR4 | | | 35,779.07 | 33,450,169 | Eldjarn GH (2023) | 37794188 |

| Trait | ID | Cases | Controls | Sample size | Number of variants | Author | PMID[a] |
|---|---|---|---|---|---|---|---|
| Chondroadherin | CHAD | | | 35,739.52 | 33,450,167 | Eldjarn GH (2023) | 37794188 |
| Choline/ethanolamine kinase | CHKB | | | 35,739.52 | 33,450,168 | Eldjarn GH (2023) | 37794188 |
| Beta-Ala-His dipeptidase | CNDP1 | | | 35,739.52 | 33,450,167 | Eldjarn GH (2023) | 37794188 |
| Ceruloplasmin | CP | | | 35,779.07 | 33,450,173 | Eldjarn GH (2023) | 37794188 |
| Probable carboxypeptidase X1 | CPXM1 | | | 35,779.07 | 33,450,173 | Eldjarn GH (2023) | 37794188 |
| Cysteine-rich with EGF-like domain protein 1 | CRELD1 | | | 35,739.52 | 33,450,163 | Eldjarn GH (2023) | 37794188 |
| Cysteine-rich secretory protein LCCL domain-containing 2 | CRISPLD2 | | | 35,739.52 | 33,450,167 | Eldjarn GH (2023) | 37794188 |
| C-reactive protein | CRP | | | 35,739.52 | 33,450,168 | Eldjarn GH (2023) | 37794188 |
| Glutamyl aminopeptidase | ENPEP | | | 35,739.52 | 33,450,165 | Eldjarn GH (2023) | 37794188 |
| Protein FAM177A1 | FAM177A1 | | | 35,779.07 | 33,450,170 | Eldjarn GH (2023) | 37794188 |
| Protein FAM3B | FAM3B | | | 35,779.07 | 33,450,173 | Eldjarn GH (2023) | 37794188 |
| Low affinity immunoglobulin gamma Fc region receptor II-b | FCGR2B | | | 35,739.52 | 33,450,163 | Eldjarn GH (2023) | 37794188 |
| Fibroblast growth factor-binding protein 1 | FGFBP1 | | | 35,739.52 | 33,450,168 | Eldjarn GH (2023) | 37794188 |
| Vascular endothelial growth factor receptor 3 | FLT4 | | | 35,739.52 | 33,450,164 | Eldjarn GH (2023) | 37794188 |
| fibromodulin | FMOD | | | 35,779.07 | 33,450,173 | Eldjarn GH (2023) | 37794188 |
| Ferritin light chain | FTL | | | 35,739.52 | 33,450,168 | Eldjarn GH (2023) | 37794188 |
| Glycerol-3-phosphate dehydrogenase [NAD(+)]; cytoplasmic | GPD1 | | | 35,739.52 | 33,450,168 | Eldjarn GH (2023) | 37794188 |
| Glutathione S-transferase Mu 4 | GSTM4 | | | 35,739.52 | 33,450,167 | Eldjarn GH (2023) | 37794188 |
| Heterogeneous nuclear ribonucleoprotein A/B | HNRNPAB | | | 35,739.52 | 33,450,167 | Eldjarn GH (2023) | 37794188 |
| Bone sialoprotein 2 | IBSP | | | 35,739.52 | 33,450,167 | Eldjarn GH (2023) | 37794188 |
| Isocitrate dehydrogenase [NADP] cytoplasmic | IDH1 | | | 35,739.52 | 33,450,167 | Eldjarn GH (2023) | 37794188 |
| Insulin-like growth factor-binding protein 1 | IGFBP1 | | | 35,739.52 | 33,450,168 | Eldjarn GH (2023) | 37794188 |

| Trait | ID | Cases | Controls | Sample size | Number of variants | Author | PMID[a] |
|---|---|---|---|---|---|---|---|
| Immunoglobulin D | IGL_IGHD_IGK | | | 35,739.52 | 33,450,168 | Eldjarn GH (2023) | 37794188 |
| Inhibin beta C chain | INHBC | | | 35,779.07 | 33,450,173 | Eldjarn GH (2023) | 37794188 |
| Inter-alpha-trypsin inhibitor heavy chain H4 | ITIH4 | | | 35,779.07 | 33,450,173 | Eldjarn GH (2023) | 37794188 |
| Beta-klotho | KLB | | | 35,739.52 | 33,450,166 | Eldjarn GH (2023) | 37794188 |
| Lymphocyte activation gene 3 protein | LAG3 | | | 35,739.52 | 33,450,167 | Eldjarn GH (2023) | 37794188 |
| Cytosol aminopeptidase | LAP3 | | | 35,739.52 | 33,450,168 | Eldjarn GH (2023) | 37794188 |
| L-lactate dehydrogenase A chain | LDHA | | | 35,739.52 | 33,450,167 | Eldjarn GH (2023) | 37794188 |
| Galectin-3-binding protein | LGALS3BP | | | 35,739.52 | 33,450,167 | Eldjarn GH (2023) | 37794188 |
| Legumain | LGMN | | | 35,739.52 | 33,450,166 | Eldjarn GH (2023) | 37794188 |
| Collagenase 3 | MMP13 | | | 35,739.52 | 33,450,168 | Eldjarn GH (2023) | 37794188 |
| Neutrophil collagenase | MMP8 | | | 35,739.52 | 33,450,163 | Eldjarn GH (2023) | 37794188 |
| Hepatocyte growth factor-like protein | MST1 | | | 35,739.52 | 33,450,163 | Eldjarn GH (2023) | 37794188 |
| Muscle; skeletal receptor tyrosine-protein kinase | MUSK | | | 35,739.52 | 33,450,167 | Eldjarn GH (2023) | 37794188 |
| Interferon-induced GTP-binding protein Mx1 | MX1 | | | 35,779.07 | 33,450,174 | Eldjarn GH (2023) | 37794188 |
| Sialic acid synthase | NANS | | | 35,739.52 | 33,450,168 | Eldjarn GH (2023) | 37794188 |
| Neutrophil cytosol factor 1 | NCF1 | | | 35,739.52 | 33,450,166 | Eldjarn GH (2023) | 37794188 |
| Olfactomedin-like protein 3 | OLFML3 | | | 35,739.52 | 33,450,167 | Eldjarn GH (2023) | 37794188 |
| Peptidyl-glycine alpha-amidating monooxygenase | PAM | | | 35,779.07 | 33,450,169 | Eldjarn GH (2023) | 37794188 |
| Proprotein convertase subtilisin/kexin type 9 | PCSK9 | | | 35,739.52 | 33,450,167 | Eldjarn GH (2023) | 37794188 |
| Proenkephalin-A | PENK | | | 35,739.52 | 33,450,166 | Eldjarn GH (2023) | 37794188 |
| Peptidoglycan recognition protein 1 | PGLYRP1 | | | 35,739.52 | 33,450,167 | Eldjarn GH (2023) | 37794188 |
| Pyruvate kinase PKM | PKM | | | 35,739.52 | 33,450,168 | Eldjarn GH (2023) | 37794188 |

| Trait | ID | Cases | Controls | Sample size | Number of variants | Author | PMID[a] |
|---|---|---|---|---|---|---|---|
| Inorganic pyrophosphatase | PPA1 | | | 35,739.52 | 33,450,167 | Eldjarn GH (2023) | 37794188 |
| Peroxiredoxin-6 | PRDX6 | | | 35,739.52 | 33,450,167 | Eldjarn GH (2023) | 37794188 |
| Myeloblastin | PRTN3 | | | 35,739.52 | 33,450,167 | Eldjarn GH (2023) | 37794188 |
| Glycogen phosphorylase; liver form | PYGL | | | 35,739.52 | 33,450,167 | Eldjarn GH (2023) | 37794188 |
| Calcipressin-1 | RCAN1 | | | 35,739.52 | 33,450,167 | Eldjarn GH (2023) | 37794188 |
| Regenerating islet-derived protein 3-alpha | REG3A | | | 35,739.52 | 33,450,167 | Eldjarn GH (2023) | 37794188 |
| Eosinophil cationic protein | RNASE3 | | | 35,739.52 | 33,450,164 | Eldjarn GH (2023) | 37794188 |
| Inactive tyrosine-protein kinase transmembrane receptor ROR1 | ROR1 | | | 35,739.52 | 33,450,167 | Eldjarn GH (2023) | 37794188 |
| Serum amyloid A-1 protein | SAA1 | | | 35,739.52 | 33,450,168 | Eldjarn GH (2023) | 37794188 |
| Semaphorin-3G | SEMA3G | | | 35,739.52 | 33,450,167 | Eldjarn GH (2023) | 37794188 |
| Protein Z-dependent protease inhibitor | SERPINA10 | | | 35,739.52 | 33,450,163 | Eldjarn GH (2023) | 37794188 |
| Alpha-2-antiplasmin | SERPINF2 | | | 35,739.52 | 33,450,167 | Eldjarn GH (2023) | 37794188 |
| Plasma protease C1 inhibitor | SERPING1 | | | 35,739.52 | 33,450,167 | Eldjarn GH (2023) | 37794188 |
| Pulmonary surfactant-associated protein D | SFTPD | | | 35,739.52 | 33,450,165 | Eldjarn GH (2023) | 37794188 |
| Acid sphingomyelinase-like phosphodiesterase 3a | SMPDL3A | | | 35,779.07 | 33,450,173 | Eldjarn GH (2023) | 37794188 |
| Alpha-synuclein | SNCA | | | 35,739.52 | 33,450,167 | Eldjarn GH (2023) | 37794188 |
| Sorbitol dehydrogenase | SORD | | | 35,739.52 | 33,450,167 | Eldjarn GH (2023) | 37794188 |
| Synaptic vesicle membrane protein VAT-1 homolog | VAT1 | | | 35,739.52 | 33,450,167 | Eldjarn GH (2023) | 37794188 |
| **outcome** | | | | | | | |
| Acute infective polyneuritis/Guillain-Barre syndrome | GBS | 174 | 420,299 | 420,473.00 | 28,987,534 | UKBB (NA) | |

[a] identification number of the publication in the PubMed database.

**Table 2**: Summary of the instrumental variables used in the MR with *cis*-pQTL

| Exposure[a] | IVs[b] | $R^2$ | $F_{st}$ |
|---|---|---|---|
| **Acute infective polyneuritis/Guillain-Barre syndrome** | | | |
| ACADSB | 6 | 0.011 | 69.4 |
| ACBD6 | 1 | 0.001 | 49.6 |
| ACP5 | 16 | 0.075 | 181.7 |
| ACY1 | 4 | 0.029 | 271.9 |
| AGER | 19 | 0.088 | 182.7 |
| APOC1 | 47 | 0.087 | 72.8 |
| AK1 | 1 | 0.001 | 44.5 |
| ALDH1A1 | 6 | 0.008 | 48.2 |
| CHAD | 1 | 0.001 | 35.6 |
| APOA1 | 14 | 0.045 | 119.7 |
| APOB | 1 | 0.002 | 67.3 |
| APOE | 36 | 0.107 | 119.2 |
| FTL | 1 | 0.001 | 45.6 |
| ARRB1 | 1 | 0.003 | 97.8 |
| ASL | 5 | 0.013 | 95.5 |
| LDHA | 2 | 0.004 | 73.2 |
| ATF6B | 22 | 0.053 | 91.1 |
| PENK | 70 | 0.367 | 296.6 |
| BPIFB1 | 55 | 0.255 | 223.8 |
| SAA1 | 43 | 0.173 | 175.0 |
| C1R | 4 | 0.006 | 52.3 |
| C4A_C4B | 52 | 0.532 | 786.3 |
| CA11 | 4 | 0.012 | 113.5 |
| CCN4 | 109 | 0.413 | 231.1 |
| CFHR4 | 51 | 0.625 | 1170.7 |
| CHKB | 18 | 0.037 | 77.2 |
| CNDP1 | 62 | 0.200 | 145.1 |
| CP | 2 | 0.004 | 83.7 |
| CPXM1 | 66 | 0.225 | 158.4 |
| CRELD1 | 101 | 0.603 | 539.3 |
| CRISPLD2 | 48 | 0.142 | 123.6 |
| CRP | 13 | 0.035 | 100.8 |
| ENPEP | 75 | 0.304 | 209.0 |

| Exposure[a] | IVs[b] | $R^2$ | $F_{st}$ |
|---|---|---|---|
| FAM177A1 | 47 | 0.310 | 343.2 |
| FAM3B | 18 | 0.056 | 117.6 |
| FCGR2B | 102 | 0.908 | 3475.3 |
| FGFBP1 | 4 | 0.007 | 61.5 |
| FLT4 | 10 | 0.023 | 82.9 |
| FMOD | 16 | 0.027 | 61.2 |
| GPD1 | 1 | 0.001 | 46.0 |
| GSTM4 | 95 | 0.529 | 424.2 |
| HNRNPAB | 2 | 0.003 | 58.6 |
| IBSP | 6 | 0.009 | 55.9 |
| IDH1 | 8 | 0.035 | 161.0 |
| IGFBP1 | 3 | 0.005 | 60.3 |
| IGL_IGHD_IGK | 2 | 0.005 | 103.4 |
| INHBC | 5 | 0.008 | 58.0 |
| ITIH4 | 8 | 0.013 | 61.0 |
| KLB | 68 | 0.480 | 486.0 |
| LAG3 | 7 | 0.015 | 78.6 |
| LAP3 | 5 | 0.008 | 59.6 |
| LGALS3BP | 20 | 0.058 | 111.2 |
| LGMN | 24 | 0.053 | 83.8 |
| MMP13 | 1 | 0.001 | 53.9 |
| MMP8 | 18 | 0.050 | 105.7 |
| MST1 | 28 | 0.417 | 916.0 |
| MUSK | 3 | 0.004 | 51.0 |
| MX1 | 44 | 0.109 | 100.2 |
| NANS | 7 | 0.011 | 55.7 |
| NCF1 | 17 | 0.071 | 160.6 |
| OLFML3 | 4 | 0.007 | 59.8 |
| PAM | 29 | 0.270 | 459.2 |
| PCSK9 | 22 | 0.058 | 99.6 |
| PGLYRP1 | 12 | 0.028 | 85.8 |
| PKM | 1 | 0.003 | 98.7 |
| PPA1 | 19 | 0.046 | 92.0 |
| PRDX6 | 3 | 0.006 | 68.9 |
| PRTN3 | 74 | 0.193 | 115.5 |

| Exposure[a] | IVs[b] | $R^2$ | $F_{st}$ |
|---|---|---|---|
| PYGL | 3 | 0.010 | 116.5 |
| RCAN1 | 1 | 0.002 | 61.6 |
| REG3A | 38 | 0.101 | 106.3 |
| RNASE3 | 38 | 0.132 | 143.3 |
| ROR1 | 29 | 0.093 | 127.7 |
| SEMA3G | 17 | 0.056 | 126.1 |
| SERPINA10 | 112 | 0.460 | 272.0 |
| SERPINF2 | 15 | 0.030 | 72.8 |
| SERPING1 | 53 | 0.295 | 282.9 |
| SFTPD | 54 | 0.359 | 372.7 |
| SMPDL3A | 12 | 0.045 | 142.5 |
| SNCA | 1 | 0.001 | 40.0 |
| SORD | 19 | 0.050 | 99.4 |
| VAT1 | 3 | 0.004 | 51.8 |

[a] Exposure abbreviations are defined in **Table 1**. [b] Number of instrumental variables (IV).

**Table 3**: Result of the Mendelian randomization using cis-pQTL

| Exposure[a] | Method[b] | IV[b] | Beta | SE | *p*-value |
|---|---|---|---|---|---|
| **Acute infective polyneuritis/Guillain-Barre syndrome** | | | | | |
| ACADSB | IVW (fe) | 6 | 0.424 | 0.704 | 0.547 |
| | MR Egger | 6 | 0.155 | 1.606 | 0.928 |
| | WM | 6 | 0.316 | 0.845 | 0.708 |
| | MR-PRESSO _raw | 6 | 0.424 | 0.656 | 0.547 |
| | Outlier_corrected | 6 | | | |
| | MR-RAPS | 6 | 0.429 | 0.716 | 0.549 |
| ACBD6 | Wald ratio | 1 | -0.285 | 1.681 | 0.865 |
| | MR-RAPS | 1 | -0.285 | 1.746 | 0.870 |
| ACP5 | IVW (fe) | 16 | -0.350 | 0.258 | 0.174 |
| | MR Egger | 16 | -0.908 | 0.564 | 0.130 |
| | WM | 16 | -0.459 | 0.348 | 0.187 |
| | MR-PRESSO _raw | 16 | -0.350 | 0.237 | 0.160 |
| | Outlier_corrected | 16 | | | |
| | MR-RAPS | 16 | -0.352 | 0.259 | 0.175 |
| ACY1 | IVW (fe) | 4 | -0.119 | 0.434 | 0.784 |
| | MR Egger | 4 | -0.304 | 0.596 | 0.661 |
| | WM | 4 | 0.220 | 0.508 | 0.664 |
| | MR-PRESSO _raw | 4 | -0.119 | 0.307 | 0.725 |
| | Outlier_corrected | 4 | | | |
| | MR-RAPS | 4 | -0.119 | 0.436 | 0.785 |
| AGER | IVW (fe) | 19 | -0.325 | 0.197 | 0.098 |
| | MR Egger | 19 | -0.157 | 0.309 | 0.617 |
| | WM | 19 | -0.133 | 0.254 | 0.599 |
| | MR-PRESSO _raw | 19 | -0.325 | 0.139 | $3.1 \times 10^{-2}$ |
| | Outlier_corrected | 19 | | | |
| | MR-RAPS | 19 | -0.326 | 0.198 | 0.100 |
| AK1 | Wald ratio | 1 | 0.810 | 2.190 | 0.711 |
| | MR-RAPS | 1 | 0.810 | 2.283 | 0.723 |
| ALDH1A1 | IVW (fe) | 6 | 1.395 | 0.786 | 0.076 |
| | MR Egger | 6 | -0.440 | 2.244 | 0.854 |
| | WM | 6 | 0.851 | 0.968 | 0.379 |
| | MR-PRESSO _raw | 6 | 1.395 | 0.452 | $2.7 \times 10^{-2}$ |

| Exposure[a] | Method[b] | IV[b] | Beta | SE | *p*-value |
|---|---|---|---|---|---|
| | Outlier_corrected | 6 | | | |
| | MR-RAPS | 6 | 1.404 | 0.814 | 0.085 |
| APOA1 | IVW (fe) | 14 | 0.193 | 0.323 | 0.551 |
| | MR Egger | 14 | 0.556 | 0.504 | 0.291 |
| | WM | 14 | 0.108 | 0.435 | 0.805 |
| | MR-PRESSO _raw | 14 | 0.193 | 0.196 | 0.344 |
| | Outlier_corrected | 14 | | | |
| | MR-RAPS | 14 | 0.193 | 0.327 | 0.554 |
| APOB | Wald ratio | 1 | -0.354 | 1.589 | 0.824 |
| | MR-RAPS | 1 | -0.354 | 1.635 | 0.828 |
| APOC1 | IVW (fe) | 47 | -0.164 | 0.243 | 0.499 |
| | MR Egger | 47 | 0.231 | 0.496 | 0.644 |
| | WM | 47 | 0.046 | 0.348 | 0.895 |
| | MR-PRESSO _raw | 47 | -0.164 | 0.213 | 0.445 |
| | Outlier_corrected | 47 | | | |
| | MR-RAPS | 47 | -0.166 | 0.247 | 0.501 |
| APOE | IVW (fe) | 36 | -0.142 | 0.225 | 0.528 |
| | MR Egger | 36 | -0.672 | 0.473 | 0.164 |
| | WM | 36 | -0.344 | 0.306 | 0.260 |
| | MR-PRESSO _raw | 36 | -0.142 | 0.216 | 0.514 |
| | Outlier_corrected | 36 | | | |
| | MR-RAPS | 36 | -0.143 | 0.228 | 0.529 |
| ARRB1 | Wald ratio | 1 | 0.647 | 1.459 | 0.657 |
| | MR-RAPS | 1 | 0.647 | 1.488 | 0.663 |
| ASL | IVW (fe) | 5 | 0.095 | 0.627 | 0.879 |
| | MR Egger | 5 | -0.856 | 2.634 | 0.767 |
| | WM | 5 | 0.138 | 0.706 | 0.845 |
| | MR-PRESSO _raw | 5 | 0.095 | 0.345 | 0.796 |
| | Outlier_corrected | 5 | | | |
| | MR-RAPS | 5 | 0.096 | 0.635 | 0.880 |
| ATF6B | IVW (fe) | 22 | -0.096 | 0.316 | 0.760 |
| | MR Egger | 22 | -0.481 | 0.784 | 0.546 |
| | WM | 22 | 0.198 | 0.437 | 0.651 |
| | MR-PRESSO _raw | 22 | -0.096 | 0.293 | 0.746 |

| Exposure[a] | Method[b] | IV[b] | Beta | SE | *p*-value |
|---|---|---|---|---|---|
| | Outlier_corrected | 22 | | | |
| | MR-RAPS | 22 | -0.097 | 0.319 | 0.761 |
| BPIFB1 | IVW (fe) | 55 | 0.081 | 0.140 | 0.564 |
| | MR Egger | 55 | -0.056 | 0.273 | 0.837 |
| | WM | 55 | 0.139 | 0.227 | 0.539 |
| | MR-PRESSO _raw | 55 | 0.081 | 0.127 | 0.526 |
| | Outlier_corrected | 55 | | | |
| | MR-RAPS | 55 | 0.081 | 0.141 | 0.564 |
| C1R | IVW (fe) | 4 | -0.145 | 1.016 | 0.887 |
| | MR Egger | 4 | 1.997 | 1.925 | 0.408 |
| | WM | 4 | 0.420 | 1.228 | 0.732 |
| | MR-PRESSO _raw | 4 | -0.145 | 0.780 | 0.865 |
| | Outlier_corrected | 4 | | | |
| | MR-RAPS | 4 | -0.146 | 1.040 | 0.888 |
| C4A_C4B | IVW (fe) | 52 | 0.055 | 0.089 | 0.534 |
| | MR Egger | 52 | -0.214 | 0.172 | 0.218 |
| | WM | 52 | -0.017 | 0.128 | 0.894 |
| | MR-PRESSO _raw | 52 | 0.055 | 0.077 | 0.478 |
| | Outlier_corrected | 52 | | | |
| | MR-RAPS | 52 | 0.055 | 0.089 | 0.534 |
| CA11 | IVW (fe) | 4 | -1.347 | 0.711 | 0.058 |
| | MR Egger | 4 | -0.312 | 1.496 | 0.854 |
| | WM | 4 | -1.321 | 0.768 | 0.086 |
| | MR-PRESSO _raw | 4 | -1.347 | 0.472 | 0.065 |
| | Outlier_corrected | 4 | | | |
| | MR-RAPS | 4 | -1.350 | 0.721 | 0.061 |
| CCN4 | IVW (mre) | 109 | -0.011 | 0.095 | 0.910 |
| | MR Egger | 109 | 0.116 | 0.175 | 0.509 |
| | WM | 109 | 0.119 | 0.152 | 0.434 |
| | MR-PRESSO _raw | 109 | -0.011 | 0.095 | 0.910 |
| | Outlier_corrected | 109 | | | |
| | MR-RAPS | 109 | -0.011 | 0.094 | 0.908 |
| CFHR4 | IVW (fe) | 51 | -0.215 | 0.074 | $3.7 \times 10^{-3}$ |
| | MR Egger | 51 | -0.265 | 0.142 | 0.068 |

| Exposure[a] | Method[b] | IV[b] | Beta | SE | *p*-value |
|---|---|---|---|---|---|
| | WM | 51 | -0.274 | 0.109 | $1.2 \times 10^{-2}$ |
| | MR-PRESSO _raw | 51 | -0.215 | 0.063 | $1.2 \times 10^{-3}$ |
| | Outlier_corrected | 51 | | | |
| | MR-RAPS | 51 | -0.215 | 0.074 | $3.8 \times 10^{-3}$ |
| CHAD | Wald ratio | 1 | -0.934 | 2.450 | 0.703 |
| | MR-RAPS | 1 | -0.934 | 2.586 | 0.718 |
| CHKB | IVW (fe) | 18 | -0.181 | 0.404 | 0.654 |
| | MR Egger | 18 | -0.012 | 0.716 | 0.987 |
| | WM | 18 | 0.187 | 0.535 | 0.726 |
| | MR-PRESSO _raw | 18 | -0.181 | 0.355 | 0.616 |
| | Outlier_corrected | 18 | | | |
| | MR-RAPS | 18 | -0.183 | 0.411 | 0.656 |
| CNDP1 | IVW (fe) | 62 | -0.505 | 0.152 | $9.1 \times 10^{-4}$ |
| | MR Egger | 62 | -0.850 | 0.291 | $4.9 \times 10^{-3}$ |
| | WM | 62 | -0.675 | 0.233 | $3.9 \times 10^{-3}$ |
| | MR-PRESSO _raw | 62 | -0.505 | 0.147 | $1.1 \times 10^{-3}$ |
| | Outlier_corrected | 62 | | | |
| | MR-RAPS | 62 | -0.508 | 0.154 | $9.5 \times 10^{-4}$ |
| CP | IVW (fe) | 2 | 0.440 | 0.838 | 0.600 |
| | MR-RAPS | 2 | 0.442 | 0.853 | 0.604 |
| CPXM1 | IVW (fe) | 66 | 0.261 | 0.147 | 0.075 |
| | MR Egger | 66 | 0.204 | 0.244 | 0.406 |
| | WM | 66 | 0.229 | 0.220 | 0.299 |
| | MR-PRESSO _raw | 66 | 0.261 | 0.130 | $4.9 \times 10^{-2}$ |
| | Outlier_corrected | 66 | | | |
| | MR-RAPS | 66 | 0.263 | 0.148 | 0.076 |
| CRELD1 | IVW (fe) | 101 | 0.067 | 0.086 | 0.433 |
| | MR Egger | 101 | 0.063 | 0.136 | 0.642 |
| | WM | 101 | 0.071 | 0.145 | 0.623 |
| | MR-PRESSO _raw | 101 | 0.067 | 0.083 | 0.417 |
| | Outlier_corrected | 101 | | | |
| | MR-RAPS | 101 | 0.068 | 0.086 | 0.433 |
| CRISPLD2 | IVW (fe) | 48 | -0.750 | 0.183 | $4.0 \times 10^{-5}$ |
| | MR Egger | 48 | -0.762 | 0.308 | $1.7 \times 10^{-2}$ |

| Exposure[a] | Method[b] | IV[b] | Beta | SE | *p*-value |
|---|---|---|---|---|---|
| | WM | 48 | -0.762 | 0.276 | $5.7 \times 10^{-3}$ |
| | MR-PRESSO _raw | 48 | -0.750 | 0.182 | $1.5 \times 10^{-4}$ |
| | Outlier_corrected | 48 | | | |
| | MR-RAPS | 48 | -0.758 | 0.185 | $4.1 \times 10^{-5}$ |
| CRP | IVW (fe) | 13 | 0.218 | 0.379 | 0.565 |
| | MR Egger | 13 | -0.048 | 0.832 | 0.955 |
| | WM | 13 | 0.100 | 0.496 | 0.840 |
| | MR-PRESSO _raw | 13 | 0.218 | 0.344 | 0.538 |
| | Outlier_corrected | 13 | | | |
| | MR-RAPS | 13 | 0.220 | 0.383 | 0.566 |
| ENPEP | IVW (mre) | 75 | -0.054 | 0.130 | 0.676 |
| | MR Egger | 75 | 0.083 | 0.231 | 0.720 |
| | WM | 75 | -0.184 | 0.208 | 0.376 |
| | MR-PRESSO _raw | 75 | -0.054 | 0.130 | 0.677 |
| | Outlier_corrected | 75 | | | |
| | MR-RAPS | 75 | -0.055 | 0.128 | 0.668 |
| FAM177A1 | IVW (fe) | 47 | -0.099 | 0.110 | 0.369 |
| | MR Egger | 47 | -0.082 | 0.176 | 0.645 |
| | WM | 47 | -0.014 | 0.170 | 0.935 |
| | MR-PRESSO _raw | 47 | -0.099 | 0.100 | 0.330 |
| | Outlier_corrected | 47 | | | |
| | MR-RAPS | 47 | -0.099 | 0.110 | 0.370 |
| FAM3B | IVW (fe) | 18 | 0.052 | 0.301 | 0.862 |
| | MR Egger | 18 | 0.038 | 0.476 | 0.937 |
| | WM | 18 | -0.205 | 0.407 | 0.615 |
| | MR-PRESSO _raw | 18 | 0.052 | 0.276 | 0.852 |
| | Outlier_corrected | 18 | | | |
| | MR-RAPS | 18 | 0.053 | 0.304 | 0.863 |
| FCGR2B | IVW (fe) | 102 | 0.116 | 0.052 | $2.5 \times 10^{-2}$ |
| | MR Egger | 102 | 0.112 | 0.096 | 0.250 |
| | WM | 102 | 0.164 | 0.087 | 0.059 |
| | MR-PRESSO _raw | 102 | 0.116 | 0.051 | $2.5 \times 10^{-2}$ |
| | Outlier_corrected | 102 | | | |
| | MR-RAPS | 102 | 0.116 | 0.052 | $2.5 \times 10^{-2}$ |

| Exposure[a] | Method[b] | IV[b] | Beta | SE | *p*-value |
|---|---|---|---|---|---|
| FGFBP1 | IVW (fe) | 4 | 1.280 | 0.875 | 0.143 |
| | MR Egger | 4 | -0.942 | 2.154 | 0.704 |
| | WM | 4 | 0.803 | 1.092 | 0.462 |
| | MR-PRESSO _raw | 4 | 1.280 | 0.641 | 0.140 |
| | Outlier_corrected | 4 | | | |
| | MR-RAPS | 4 | 1.289 | 0.897 | 0.151 |
| FLT4 | IVW (fe) | 10 | 0.818 | 0.517 | 0.113 |
| | MR Egger | 10 | 0.154 | 1.282 | 0.907 |
| | WM | 10 | 0.757 | 0.664 | 0.254 |
| | MR-PRESSO _raw | 10 | 0.818 | 0.422 | 0.084 |
| | Outlier_corrected | 10 | | | |
| | MR-RAPS | 10 | 0.824 | 0.525 | 0.117 |
| FMOD | IVW (mre) | 16 | -0.318 | 0.448 | 0.478 |
| | MR Egger | 16 | 0.307 | 0.756 | 0.690 |
| | WM | 16 | 0.548 | 0.580 | 0.345 |
| | MR-PRESSO _raw | 16 | -0.318 | 0.448 | 0.489 |
| | Outlier_corrected | 16 | | | |
| | MR-RAPS | 16 | -0.323 | 0.414 | 0.434 |
| FTL | Wald ratio | 1 | 0.596 | 2.038 | 0.770 |
| | MR-RAPS | 1 | 0.596 | 2.126 | 0.779 |
| GPD1 | Wald ratio | 1 | 0.533 | 1.258 | 0.672 |
| | MR-RAPS | 1 | 0.533 | 1.305 | 0.683 |
| GSTM4 | IVW (mre) | 95 | -0.239 | 0.083 | $4.1 \times 10^{-3}$ |
| | MR Egger | 95 | -0.217 | 0.137 | 0.115 |
| | WM | 95 | -0.140 | 0.122 | 0.248 |
| | MR-PRESSO _raw | 95 | -0.239 | 0.083 | $5.1 \times 10^{-3}$ |
| | Outlier_corrected | 95 | | | |
| | MR-RAPS | 95 | -0.240 | 0.077 | $1.7 \times 10^{-3}$ |
| HNRNPAB | IVW (fe) | 2 | 0.680 | 1.414 | 0.631 |
| | MR-RAPS | 2 | 0.683 | 1.450 | 0.638 |
| IBSP | IVW (mre) | 6 | -0.308 | 0.877 | 0.726 |
| | MR Egger | 6 | 1.053 | 3.013 | 0.744 |
| | WM | 6 | -0.077 | 0.947 | 0.935 |
| | MR-PRESSO _raw | 6 | -0.308 | 0.877 | 0.740 |

| Exposure[a] | Method[b] | IV[b] | Beta | SE | *p*-value |
|---|---|---|---|---|---|
| | Outlier_corrected | 6 | | | |
| | MR-RAPS | 6 | -0.313 | 0.769 | 0.684 |
| IDH1 | IVW (fe) | 8 | -0.491 | 0.328 | 0.135 |
| | MR Egger | 8 | -0.448 | 0.422 | 0.329 |
| | WM | 8 | -0.377 | 0.376 | 0.316 |
| | MR-PRESSO _raw | 8 | -0.491 | 0.246 | 0.086 |
| | Outlier_corrected | 8 | | | |
| | MR-RAPS | 8 | -0.492 | 0.331 | 0.137 |
| IGFBP1 | IVW (fe) | 3 | -0.957 | 1.047 | 0.361 |
| | MR Egger | 3 | 27.984 | 24.360 | 0.456 |
| | WM | 3 | -0.833 | 1.201 | 0.488 |
| | MR-RAPS | 3 | -0.967 | 1.071 | 0.367 |
| IGL_IGHD_IGK | IVW (fe) | 2 | 0.476 | 1.089 | 0.662 |
| | MR-RAPS | 2 | 0.476 | 1.108 | 0.667 |
| INHBC | IVW (fe) | 5 | -0.795 | 0.762 | 0.297 |
| | MR Egger | 5 | 1.327 | 2.180 | 0.586 |
| | WM | 5 | -0.965 | 0.872 | 0.269 |
| | MR-PRESSO _raw | 5 | -0.795 | 0.482 | 0.174 |
| | Outlier_corrected | 5 | | | |
| | MR-RAPS | 5 | -0.803 | 0.783 | 0.305 |
| ITIH4 | IVW (mre) | 8 | -1.013 | 0.749 | 0.176 |
| | MR Egger | 8 | -5.487 | 2.029 | $3.5 \times 10^{-2}$ |
| | WM | 8 | -1.873 | 0.864 | $3.0 \times 10^{-2}$ |
| | MR-PRESSO _raw | 8 | -1.013 | 0.749 | 0.218 |
| | Outlier_corrected | 8 | | | |
| | MR-RAPS | 8 | -1.030 | 0.640 | 0.107 |
| KLB | IVW (fe) | 68 | 0.129 | 0.091 | 0.157 |
| | MR Egger | 68 | 0.227 | 0.171 | 0.190 |
| | WM | 68 | 0.029 | 0.141 | 0.836 |
| | MR-PRESSO _raw | 68 | 0.129 | 0.084 | 0.131 |
| | Outlier_corrected | 68 | | | |
| | MR-RAPS | 68 | 0.129 | 0.092 | 0.157 |
| LAG3 | IVW (fe) | 7 | 0.010 | 0.544 | 0.985 |
| | MR Egger | 7 | 0.505 | 1.442 | 0.740 |

| Exposure[a] | Method[b] | IV[b] | Beta | SE | *p*-value |
|---|---|---|---|---|---|
| | WM | 7 | 0.037 | 0.648 | 0.954 |
| | MR-PRESSO _raw | 7 | 0.010 | 0.240 | 0.968 |
| | Outlier_corrected | 7 | | | |
| | MR-RAPS | 7 | 0.010 | 0.555 | 0.986 |
| LAP3 | IVW (fe) | 5 | -0.655 | 0.814 | 0.422 |
| | MR Egger | 5 | -1.083 | 1.550 | 0.535 |
| | WM | 5 | -0.473 | 0.922 | 0.608 |
| | MR-PRESSO _raw | 5 | -0.655 | 0.347 | 0.133 |
| | Outlier_corrected | 5 | | | |
| | MR-RAPS | 5 | -0.656 | 0.836 | 0.433 |
| LDHA | IVW (fe) | 2 | -1.043 | 1.580 | 0.509 |
| | MR-RAPS | 2 | -1.048 | 1.622 | 0.518 |
| LGALS3BP | IVW (fe) | 20 | -0.101 | 0.279 | 0.716 |
| | MR Egger | 20 | 0.649 | 0.478 | 0.192 |
| | WM | 20 | -0.063 | 0.407 | 0.877 |
| | MR-PRESSO _raw | 20 | -0.101 | 0.268 | 0.710 |
| | Outlier_corrected | 20 | | | |
| | MR-RAPS | 20 | -0.102 | 0.282 | 0.716 |
| LGMN | IVW (fe) | 24 | 0.499 | 0.304 | 0.100 |
| | MR Egger | 24 | 0.725 | 0.577 | 0.222 |
| | WM | 24 | 0.535 | 0.434 | 0.218 |
| | MR-PRESSO _raw | 24 | 0.499 | 0.273 | 0.080 |
| | Outlier_corrected | 24 | | | |
| | MR-RAPS | 24 | 0.503 | 0.308 | 0.103 |
| MMP13 | Wald ratio | 1 | -0.989 | 1.918 | 0.606 |
| | MR-RAPS | 1 | -0.989 | 1.985 | 0.618 |
| MMP8 | IVW (fe) | 18 | 0.643 | 0.302 | $3.3 \times 10^{-2}$ |
| | MR Egger | 18 | 0.500 | 0.443 | 0.276 |
| | WM | 18 | 0.820 | 0.447 | 0.067 |
| | MR-PRESSO _raw | 18 | 0.643 | 0.276 | $3.2 \times 10^{-2}$ |
| | Outlier_corrected | 18 | | | |
| | MR-RAPS | 18 | 0.649 | 0.305 | $3.4 \times 10^{-2}$ |
| MST1 | IVW (fe) | 28 | -0.094 | 0.077 | 0.223 |
| | MR Egger | 28 | 0.067 | 0.187 | 0.722 |

| Exposure[a] | Method[b] | IV[b] | Beta | SE | *p*-value |
|---|---|---|---|---|---|
| | WM | 28 | -0.047 | 0.109 | 0.665 |
| | MR-PRESSO _raw | 28 | -0.094 | 0.075 | 0.217 |
| | Outlier_corrected | 28 | | | |
| | MR-RAPS | 28 | -0.094 | 0.077 | 0.223 |
| MUSK | IVW (fe) | 3 | 1.032 | 1.166 | 0.376 |
| | MR Egger | 3 | 1.794 | 3.460 | 0.696 |
| | WM | 3 | 1.249 | 1.282 | 0.330 |
| | MR-RAPS | 3 | 1.033 | 1.203 | 0.391 |
| MX1 | IVW (fe) | 44 | 0.187 | 0.219 | 0.393 |
| | MR Egger | 44 | 0.735 | 0.443 | 0.105 |
| | WM | 44 | 0.153 | 0.312 | 0.625 |
| | MR-PRESSO _raw | 44 | 0.187 | 0.182 | 0.309 |
| | Outlier_corrected | 44 | | | |
| | MR-RAPS | 44 | 0.188 | 0.222 | 0.396 |
| NANS | IVW (fe) | 7 | -0.831 | 0.707 | 0.240 |
| | MR Egger | 7 | -1.314 | 2.021 | 0.544 |
| | WM | 7 | -0.776 | 0.846 | 0.359 |
| | MR-PRESSO _raw | 7 | -0.831 | 0.281 | $2.5 \times 10^{-2}$ |
| | Outlier_corrected | 7 | | | |
| | MR-RAPS | 7 | -0.833 | 0.727 | 0.252 |
| NCF1 | IVW (fe) | 17 | -0.106 | 0.195 | 0.587 |
| | MR Egger | 17 | 0.210 | 0.323 | 0.526 |
| | WM | 17 | 0.013 | 0.244 | 0.957 |
| | MR-PRESSO _raw | 17 | -0.106 | 0.169 | 0.539 |
| | Outlier_corrected | 17 | | | |
| | MR-RAPS | 17 | -0.106 | 0.197 | 0.590 |
| OLFML3 | IVW (fe) | 4 | 0.952 | 0.835 | 0.254 |
| | MR Egger | 4 | 0.472 | 3.961 | 0.916 |
| | WM | 4 | 0.820 | 0.974 | 0.400 |
| | MR-PRESSO _raw | 4 | 0.952 | 0.257 | $3.4 \times 10^{-2}$ |
| | Outlier_corrected | 4 | | | |
| | MR-RAPS | 4 | 0.953 | 0.859 | 0.267 |
| PAM | IVW (mre) | 29 | 0.078 | 0.126 | 0.537 |
| | MR Egger | 29 | 0.161 | 0.251 | 0.526 |

| Exposure[a] | Method[b] | IV[b] | Beta | SE | *p*-value |
|---|---|---|---|---|---|
| | WM | 29 | 0.224 | 0.173 | 0.196 |
| | MR-PRESSO _raw | 29 | 0.078 | 0.126 | 0.542 |
| | Outlier_corrected | 29 | | | |
| | MR-RAPS | 29 | 0.078 | 0.118 | 0.508 |
| PCSK9 | IVW (fe) | 22 | -0.440 | 0.279 | 0.115 |
| | MR Egger | 22 | -0.603 | 0.430 | 0.176 |
| | WM | 22 | -0.517 | 0.388 | 0.183 |
| | MR-PRESSO _raw | 22 | -0.440 | 0.174 | $1.9 \times 10^{-2}$ |
| | Outlier_corrected | 22 | | | |
| | MR-RAPS | 22 | -0.441 | 0.283 | 0.119 |
| PENK | IVW (mre) | 70 | -0.256 | 0.114 | $2.5 \times 10^{-2}$ |
| | MR Egger | 70 | 0.210 | 0.228 | 0.361 |
| | WM | 70 | -0.440 | 0.174 | $1.1 \times 10^{-2}$ |
| | MR-PRESSO _raw | 70 | -0.256 | 0.114 | $2.8 \times 10^{-2}$ |
| | Outlier_corrected | 70 | | | |
| | MR-RAPS | 70 | -0.257 | 0.107 | $1.6 \times 10^{-2}$ |
| PGLYRP1 | IVW (fe) | 12 | 0.764 | 0.419 | 0.068 |
| | MR Egger | 12 | -0.173 | 0.952 | 0.859 |
| | WM | 12 | 0.858 | 0.530 | 0.105 |
| | MR-PRESSO _raw | 12 | 0.764 | 0.376 | 0.067 |
| | Outlier_corrected | 12 | | | |
| | MR-RAPS | 12 | 0.771 | 0.426 | 0.070 |
| PKM | Wald ratio | 1 | 0.353 | 1.587 | 0.824 |
| | MR-RAPS | 1 | 0.353 | 1.616 | 0.827 |
| PPA1 | IVW (mre) | 19 | 0.152 | 0.363 | 0.675 |
| | MR Egger | 19 | -0.639 | 0.833 | 0.453 |
| | WM | 19 | 0.269 | 0.515 | 0.601 |
| | MR-PRESSO _raw | 19 | 0.152 | 0.363 | 0.680 |
| | Outlier_corrected | 19 | | | |
| | MR-RAPS | 19 | 0.154 | 0.355 | 0.665 |
| PRDX6 | IVW (fe) | 3 | 0.416 | 0.890 | 0.640 |
| | MR Egger | 3 | -0.579 | 3.303 | 0.889 |
| | WM | 3 | 0.280 | 1.001 | 0.780 |
| | MR-RAPS | 3 | 0.416 | 0.909 | 0.647 |

| Exposure[a] | Method[b] | IV[b] | Beta | SE | *p*-value |
|---|---|---|---|---|---|
| PRTN3 | IVW (fe) | 74 | 0.223 | 0.143 | 0.119 |
| | MR Egger | 74 | 0.046 | 0.269 | 0.864 |
| | WM | 74 | 0.188 | 0.247 | 0.448 |
| | MR-PRESSO _raw | 74 | 0.223 | 0.138 | 0.110 |
| | Outlier_corrected | 74 | | | |
| | MR-RAPS | 74 | 0.225 | 0.145 | 0.120 |
| PYGL | IVW (fe) | 3 | 1.395 | 0.759 | 0.066 |
| | MR Egger | 3 | -1.249 | 1.815 | 0.616 |
| | WM | 3 | 1.589 | 0.876 | 0.070 |
| | MR-RAPS | 3 | 1.407 | 0.769 | 0.068 |
| RCAN1 | Wald ratio | 1 | -0.495 | 1.784 | 0.781 |
| | MR-RAPS | 1 | -0.495 | 1.836 | 0.787 |
| REG3A | IVW (fe) | 38 | -0.125 | 0.213 | 0.558 |
| | MR Egger | 38 | -0.142 | 0.412 | 0.733 |
| | WM | 38 | -0.317 | 0.316 | 0.316 |
| | MR-PRESSO _raw | 38 | -0.125 | 0.207 | 0.549 |
| | Outlier_corrected | 38 | | | |
| | MR-RAPS | 38 | -0.126 | 0.216 | 0.559 |
| RNASE3 | IVW (fe) | 38 | 0.125 | 0.175 | 0.474 |
| | MR Egger | 38 | 0.433 | 0.258 | 0.102 |
| | WM | 38 | 0.214 | 0.271 | 0.429 |
| | MR-PRESSO _raw | 38 | 0.125 | 0.151 | 0.412 |
| | Outlier_corrected | 38 | | | |
| | MR-RAPS | 38 | 0.126 | 0.176 | 0.476 |
| ROR1 | IVW (mre) | 29 | -0.188 | 0.245 | 0.441 |
| | MR Egger | 29 | -0.945 | 0.444 | $4.2 \times 10^{-2}$ |
| | WM | 29 | -0.330 | 0.341 | 0.334 |
| | MR-PRESSO _raw | 29 | -0.188 | 0.245 | 0.448 |
| | Outlier_corrected | 29 | | | |
| | MR-RAPS | 29 | -0.190 | 0.236 | 0.420 |
| SAA1 | IVW (fe) | 43 | -0.099 | 0.160 | 0.536 |
| | MR Egger | 43 | -0.344 | 0.251 | 0.179 |
| | WM | 43 | -0.363 | 0.235 | 0.122 |
| | MR-PRESSO _raw | 43 | -0.099 | 0.144 | 0.496 |

| Exposure[a] | Method[b] | IV[b] | Beta | SE | *p*-value |
|---|---|---|---|---|---|
| | Outlier_corrected | 43 | | | |
| | MR-RAPS | 43 | -0.099 | 0.161 | 0.537 |
| SEMA3G | IVW (mre) | 17 | 0.131 | 0.333 | 0.694 |
| | MR Egger | 17 | -0.624 | 0.662 | 0.361 |
| | WM | 17 | -0.025 | 0.420 | 0.952 |
| | MR-PRESSO _raw | 17 | 0.131 | 0.333 | 0.699 |
| | Outlier_corrected | 17 | | | |
| | MR-RAPS | 17 | 0.132 | 0.299 | 0.658 |
| SERPINA10 | IVW (mre) | 112 | -0.045 | 0.099 | 0.649 |
| | MR Egger | 112 | -0.034 | 0.159 | 0.831 |
| | WM | 112 | 0.040 | 0.167 | 0.809 |
| | MR-PRESSO _raw | 112 | -0.045 | 0.099 | 0.650 |
| | Outlier_corrected | 112 | | | |
| | MR-RAPS | 112 | -0.045 | 0.099 | 0.648 |
| SERPINF2 | IVW (mre) | 15 | 0.777 | 0.419 | 0.063 |
| | MR Egger | 15 | -0.203 | 0.906 | 0.826 |
| | WM | 15 | 0.908 | 0.552 | 0.100 |
| | MR-PRESSO _raw | 15 | 0.777 | 0.419 | 0.085 |
| | Outlier_corrected | 15 | | | |
| | MR-RAPS | 15 | 0.787 | 0.421 | 0.062 |
| SERPING1 | IVW (fe) | 53 | -0.497 | 0.118 | $2.6 \times 10^{-5}$ |
| | MR Egger | 53 | -0.611 | 0.237 | $1.3 \times 10^{-2}$ |
| | WM | 53 | -0.466 | 0.181 | $1.0 \times 10^{-2}$ |
| | MR-PRESSO _raw | 53 | -0.497 | 0.105 | $1.9 \times 10^{-5}$ |
| | Outlier_corrected | 53 | | | |
| | MR-RAPS | 53 | -0.499 | 0.119 | $2.7 \times 10^{-5}$ |
| SFTPD | IVW (fe) | 54 | 0.121 | 0.113 | 0.284 |
| | MR Egger | 54 | 0.072 | 0.215 | 0.738 |
| | WM | 54 | 0.113 | 0.179 | 0.530 |
| | MR-PRESSO _raw | 54 | 0.121 | 0.112 | 0.283 |
| | Outlier_corrected | 54 | | | |
| | MR-RAPS | 54 | 0.122 | 0.114 | 0.284 |
| SMPDL3A | IVW (fe) | 12 | -0.345 | 0.351 | 0.326 |
| | MR Egger | 12 | -0.220 | 0.658 | 0.745 |

| Exposure[a] | Method[b] | IV[b] | Beta | SE | *p*-value |
|---|---|---|---|---|---|
| | WM | 12 | -0.253 | 0.436 | 0.562 |
| | MR-PRESSO _raw | 12 | -0.345 | 0.348 | 0.343 |
| | Outlier_corrected | 12 | | | |
| | MR-RAPS | 12 | -0.347 | 0.354 | 0.327 |
| SNCA | Wald ratio | 1 | -1.513 | 2.280 | 0.507 |
| | MR-RAPS | 1 | -1.513 | 2.396 | 0.528 |
| SORD | IVW (fe) | 19 | 0.673 | 0.327 | $3.9 \times 10^{-2}$ |
| | MR Egger | 19 | 0.462 | 0.580 | 0.437 |
| | WM | 19 | 0.399 | 0.442 | 0.366 |
| | MR-PRESSO _raw | 19 | 0.673 | 0.202 | $3.7 \times 10^{-3}$ |
| | Outlier_corrected | 19 | | | |
| | MR-RAPS | 19 | 0.675 | 0.332 | $4.2 \times 10^{-2}$ |
| VAT1 | IVW (fe) | 3 | 0.098 | 1.289 | 0.939 |
| | MR Egger | 3 | 7.042 | 5.804 | 0.439 |
| | WM | 3 | 0.018 | 1.408 | 0.990 |
| | MR-RAPS | 3 | 0.100 | 1.321 | 0.940 |

[a] Exposure abbreviations are defined in **eTable 1**.[b] Mendelian randomization method: IVW (mre), inverse variance weighted (IVW), multiplicative random effects; WM, Weighted median; MR-RAPS, Mendelian randomization using the robust adjusted profile score; MRlap, Mendelian randomization using the latent heritable confounders adjustment.[b] Number of instrumental variables.

**Table 4**: Primary analysis of the Mendelian randomization using cis-pQTL

| | | | | Inverse variance weighted | | | | | Intercept | Association | | |
|---|---|---|---|---|---|---|---|---|---|---|---|---|
| Exposure[a] | IVs[b] | Method | Beta | OR (95% CI) | *p*-value | *p*-FDR[c] | $I^2$ | $I^2_{GX}$ | *p*-value[d] | Consistent[e] | Nominal[f] | Classification[g] |
| **Acute infective polyneuritis/Guillain-Barre syndrome** | | | | | | | | | | | | |
| ACADSB | 6 | IVW (fe) | 0.424 | 1.53 (0.38 - 6.07) | 0.547 | 0.862 | 0.0% | 94.1% | 0.860 | | | Not_associated |
| ACBD6 | 1 | Wald ratio | -0.285 | | 0.865 | 0.920 | | | | yes | | Not_associated |
| ACP5 | 16 | IVW (fe) | -0.350 | 0.70 (0.43 - 1.17) | 0.174 | 0.578 | 0.0% | 97.4% | 0.285 | | | Not_associated |
| ACY1 | 4 | IVW (fe) | -0.119 | 0.89 (0.38 - 2.08) | 0.784 | 0.881 | 0.0% | 99.4% | 0.695 | | | Not_associated |
| AGER | 19 | IVW (fe) | -0.325 | 0.72 (0.49 - 1.06) | 0.098 | 0.485 | 0.0% | 98.7% | 0.489 | yes | MRPRESSO | Not_associated |
| AK1 | 1 | Wald ratio | 0.810 | | 0.711 | 0.862 | | 100.0% | | yes | | Not_associated |
| ALDH1A1 | 6 | IVW (fe) | 1.395 | 4.04 (0.86 - 18.85) | 0.076 | 0.415 | 0.0% | 86.9% | 0.432 | yes | MRPRESSO | Not_associated |
| APOA1 | 14 | IVW (fe) | 0.193 | 1.21 (0.64 - 2.28) | 0.551 | 0.862 | 0.0% | 98.0% | 0.365 | | | Not_associated |
| APOB | 1 | Wald ratio | -0.354 | | 0.824 | 0.901 | | | | yes | | Not_associated |
| APOC1 | 47 | IVW (fe) | -0.164 | 0.85 (0.53 - 1.37) | 0.499 | 0.862 | 0.0% | 93.4% | 0.365 | | | Not_associated |

| | | Inverse variance weighted | | | | | | | Intercept | | Association | |
|---|---|---|---|---|---|---|---|---|---|---|---|---|
| Exposure[a] | IVs[b] | Method | Beta | OR (95% CI) | *p*-value | *p*-FDR[c] | $I^2$ | $I^2_{GX}$ | *p*-value[d] | Consistent[e] | Nominal[f] | Classification[g] |
| APOE | 36 | IVW (fe) | -0.142 | 0.87 (0.56 - 1.35) | 0.528 | 0.862 | 0.0% | 95.9% | 0.211 | | | Not_associated |
| ARRB1 | 1 | Wald ratio | 0.647 | | 0.657 | 0.862 | | | | yes | | Not_associated |
| ASL | 5 | IVW (fe) | 0.095 | 1.10 (0.32 - 3.76) | 0.879 | 0.920 | 0.0% | 86.3% | 0.735 | | | Not_associated |
| ATF6B | 22 | IVW (fe) | -0.096 | 0.91 (0.49 - 1.69) | 0.760 | 0.881 | 0.0% | 93.8% | 0.597 | | | Not_associated |
| BPIFB1 | 55 | IVW (fe) | 0.081 | 1.08 (0.82 - 1.43) | 0.564 | 0.862 | 0.0% | 98.4% | 0.559 | | | Not_associated |
| C1R | 4 | IVW (fe) | -0.145 | 0.87 (0.12 - 6.34) | 0.887 | 0.920 | 0.0% | 94.5% | 0.320 | | | Not_associated |
| C4A_C4B | 52 | IVW (fe) | 0.055 | 1.06 (0.89 - 1.26) | 0.534 | 0.862 | 0.0% | 99.1% | 0.073 | | | Not_associated |
| CA11 | 4 | IVW (fe) | -1.347 | 0.26 (0.06 - 1.05) | 0.058 | 0.415 | 0.0% | 97.4% | 0.514 | | | Not_associated |
| CCN4 | 109 | IVW (mre) | -0.011 | 0.99 (0.82 - 1.19) | 0.910 | 0.932 | 3.4% | 98.6% | 0.390 | | | Not_associated |
| CFHR4 | 51 | IVW (fe) | -0.215 | 0.81 (0.70 - 0.93) | $3.7 \times 10^{-3}$ | 0.067 | 0.0% | 99.8% | 0.683 | yes | IVW;WM;MRPRESSO;RAPS | Causal_robust |
| CHAD | 1 | Wald ratio | -0.934 | | 0.703 | 0.862 | | 100.0% | | yes | | Not_associated |

| | | | Inverse variance weighted | | | | | | Intercept | Association | | |
|---|---|---|---|---|---|---|---|---|---|---|---|---|
| Exposure[a] | IVs[b] | Method | Beta | OR (95% CI) | *p*-value | *p*-FDR[c] | $I^2$ | $I^2_{GX}$ | *p*-value[d] | Consistent[e] | Nominal[f] | Classification[g] |
| CHKB | 18 | IVW (fe) | -0.181 | 0.83 (0.38 - 1.84) | 0.654 | 0.862 | 0.0% | 96.4% | 0.778 | | | Not_associated |
| CNDP1 | 62 | IVW (fe) | -0.505 | 0.60 (0.45 - 0.81) | $9.1 \times 10^{-4}$ | $2.5 \times 10^{-2}$ | 0.0% | 97.0% | 0.168 | yes | IVW;WM;MRPRESSO;RAPS | Causal_robust |
| CP | 2 | IVW (fe) | 0.440 | | 0.600 | 0.862 | 0.0% | 96.1% | | yes | | Not_associated |
| CPXM1 | 66 | IVW (fe) | 0.261 | 1.30 (0.97 - 1.73) | 0.075 | 0.415 | 0.0% | 97.9% | 0.772 | yes | MRPRESSO | Not_associated |
| CRELD1 | 101 | IVW (fe) | 0.067 | 1.07 (0.90 - 1.27) | 0.433 | 0.862 | 0.0% | 99.4% | 0.971 | | | Not_associated |
| CRISPLD2 | 48 | IVW (fe) | -0.750 | 0.47 (0.33 - 0.68) | $4.0 \times 10^{-5}$ | $1.6 \times 10^{-3}$ | 0.0% | 97.4% | 0.964 | yes | IVW;WM;MRPRESSO;RAPS | Causal_robust |
| CRP | 13 | IVW (fe) | 0.218 | 1.24 (0.59 - 2.61) | 0.565 | 0.862 | 0.0% | 95.4% | 0.726 | | | Not_associated |
| ENPEP | 75 | IVW (mre) | -0.054 | 0.95 (0.73 - 1.22) | 0.676 | 0.862 | 4.6% | 98.0% | 0.473 | | | Not_associated |
| FAM177A1 | 47 | IVW (fe) | -0.099 | 0.91 (0.73 - 1.12) | 0.369 | 0.862 | 0.0% | 99.3% | 0.903 | | | Not_associated |
| FAM3B | 18 | IVW (fe) | 0.052 | 1.05 (0.58 - 1.90) | 0.862 | 0.920 | 0.0% | 97.9% | 0.971 | | | Not_associated |
| FCGR2B | 102 | IVW (fe) | 0.116 | 1.12 (1.01 - 1.24) | $2.5 \times 10^{-2}$ | 0.293 | 0.0% | 99.8% | 0.959 | yes | IVW;MRPRESSO;RAPS | Causal_robust |

| | | | Inverse variance weighted | | | | | | Intercept | | Association | |
|---|---|---|---|---|---|---|---|---|---|---|---|---|
| **Exposure**[a] | **IVs**[b] | **Method** | **Beta** | **OR (95% CI)** | ***p*-value** | ***p*-FDR**[c] | **$I^2$** | **$I^2_{GX}$** | ***p*-value**[d] | **Consistent**[e] | **Nominal**[f] | **Classification**[g] |
| FGFBP1 | 4 | IVW (fe) | 1.280 | 3.60 (0.65 - 19.98) | 0.143 | 0.535 | 0.0% | 92.4% | 0.376 | | | Not_associated |
| FLT4 | 10 | IVW (fe) | 0.818 | 2.27 (0.82 - 6.24) | 0.113 | 0.489 | 0.0% | 93.3% | 0.586 | | | Not_associated |
| FMOD | 16 | IVW (mre) | -0.318 | 0.73 (0.30 - 1.75) | 0.478 | 0.862 | 17.6% | 95.4% | 0.322 | | | Not_associated |
| FTL | 1 | Wald ratio | 0.596 | | 0.770 | 0.881 | | | | yes | | Not_associated |
| GPD1 | 1 | Wald ratio | 0.533 | | 0.672 | 0.862 | | | | yes | | Not_associated |
| GSTM4 | 95 | IVW (mre) | -0.239 | 0.79 (0.67 - 0.93) | $4.1 \times 10^{-3}$ | 0.067 | 15.8% | 99.4% | 0.842 | yes | IVW;MRPRESSO;RAPS | Causal_robust |
| HNRNPAB | 2 | IVW (fe) | 0.680 | | 0.631 | 0.862 | 0.0% | 76.0% | | yes | | Not_associated |
| IBSP | 6 | IVW (mre) | -0.308 | 0.74 (0.13 - 4.10) | 0.726 | 0.862 | 25.6% | 86.0% | 0.659 | | | Not_associated |
| IDH1 | 8 | IVW (fe) | -0.491 | 0.61 (0.32 - 1.16) | 0.135 | 0.526 | 0.0% | 99.0% | 0.876 | | | Not_associated |
| IGFBP1 | 3 | IVW (fe) | -0.957 | | 0.361 | 0.862 | 0.0% | 0.0% | 0.445 | yes | | Not_associated |
| IGL_IGHD_IGK | 2 | IVW (fe) | 0.476 | | 0.662 | 0.862 | 0.0% | 0.0% | | yes | | Not_associated |
| INHBC | 5 | IVW (fe) | -0.795 | 0.45 (0.10 - 2.01) | 0.297 | 0.811 | 0.0% | 86.1% | 0.375 | | | Not_associated |

| | | Inverse variance weighted | | | | | | | Intercept | Association | | |
|---|---|---|---|---|---|---|---|---|---|---|---|---|
| Exposure[a] | IVs[b] | Method | Beta | OR (95% CI) | *p*-value | *p*-FDR[c] | $I^2$ | $I^2_{GX}$ | *p*-value[d] | Consistent[e] | Nominal[f] | Classification[g] |
| ITIH4 | 8 | IVW (mre) | -1.013 | 0.36 (0.08 - 1.58) | 0.176 | 0.578 | 29.1% | 88.5% | 0.059 | yes | WM | Not_associated |
| KLB | 68 | IVW (fe) | 0.129 | 1.14 (0.95 - 1.36) | 0.157 | 0.560 | 0.0% | 99.1% | 0.503 | | | Not_associated |
| LAG3 | 7 | IVW (fe) | 0.010 | 1.01 (0.35 - 2.93) | 0.985 | 0.985 | 0.0% | 90.7% | 0.726 | | | Not_associated |
| LAP3 | 5 | IVW (fe) | -0.655 | 0.52 (0.11 - 2.56) | 0.422 | 0.862 | 0.0% | 95.3% | 0.766 | | | Not_associated |
| LDHA | 2 | IVW (fe) | -1.043 | | 0.509 | 0.862 | 0.0% | 97.5% | | yes | | Not_associated |
| LGALS3BP | 20 | IVW (fe) | -0.101 | 0.90 (0.52 - 1.56) | 0.716 | 0.862 | 0.0% | 97.2% | 0.070 | | | Not_associated |
| LGMN | 24 | IVW (fe) | 0.499 | 1.65 (0.91 - 2.98) | 0.100 | 0.485 | 0.0% | 95.4% | 0.649 | | | Not_associated |
| MMP13 | 1 | Wald ratio | -0.989 | | 0.606 | 0.862 | | | | yes | | Not_associated |
| MMP8 | 18 | IVW (fe) | 0.643 | 1.90 (1.05 - 3.44) | $3.3 \times 10^{-2}$ | 0.339 | 0.0% | 97.9% | 0.665 | yes | IVW;MRPRESSO;RAPS | Causal_likely |
| MST1 | 28 | IVW (fe) | -0.094 | 0.91 (0.78 - 1.06) | 0.223 | 0.702 | 0.0% | 99.9% | 0.351 | | | Not_associated |
| MUSK | 3 | IVW (fe) | 1.032 | | 0.376 | 0.862 | 0.0% | 88.0% | 0.854 | yes | | Not_associated |

| | | | | Inverse variance weighted | | | | | Intercept | | Association | |
|---|---|---|---|---|---|---|---|---|---|---|---|---|
| Exposure[a] | IVs[b] | Method | Beta | OR (95% CI) | *p*-value | *p*-FDR[c] | $I^2$ | $I^2_{GX}$ | *p*-value[d] | Consistent[e] | Nominal[f] | Classification[g] |
| MX1 | 44 | IVW (fe) | 0.187 | 1.21 (0.79 - 1.85) | 0.393 | 0.862 | 0.0% | 95.2% | 0.163 | | | Not_associated |
| NANS | 7 | IVW (fe) | -0.831 | 0.44 (0.11 - 1.74) | 0.240 | 0.728 | 0.0% | 87.7% | 0.809 | yes | MRPRESSO | Not_associated |
| NCF1 | 17 | IVW (fe) | -0.106 | 0.90 (0.61 - 1.32) | 0.587 | 0.862 | 0.0% | 97.8% | 0.240 | | | Not_associated |
| OLFML3 | 4 | IVW (fe) | 0.952 | 2.59 (0.50 - 13.31) | 0.254 | 0.745 | 0.0% | 71.6% | 0.913 | yes | MRPRESSO | Not_associated |
| PAM | 29 | IVW (mre) | 0.078 | 1.08 (0.84 - 1.39) | 0.537 | 0.862 | 13.2% | 99.6% | 0.703 | | | Not_associated |
| PCSK9 | 22 | IVW (fe) | -0.440 | 0.64 (0.37 - 1.11) | 0.115 | 0.489 | 0.0% | 97.2% | 0.623 | yes | MRPRESSO | Not_associated |
| PENK | 70 | IVW (mre) | -0.256 | 0.77 (0.62 - 0.97) | $2.5 \times 10^{-2}$ | 0.293 | 12.9% | 98.7% | $2.3 \times 10^{-2}$ | yes | IVW;WM;MRPRESSO;RAPS | Causal_with_pleiotropy |
| PGLYRP1 | 12 | IVW (fe) | 0.764 | 2.15 (0.94 - 4.88) | 0.068 | 0.415 | 0.0% | 94.2% | 0.299 | | | Not_associated |
| PKM | 1 | Wald ratio | 0.353 | | 0.824 | 0.901 | | | | yes | | Not_associated |
| PPA1 | 19 | IVW (mre) | 0.152 | 1.16 (0.57 - 2.37) | 0.675 | 0.862 | 6.3% | 94.5% | 0.306 | | | Not_associated |
| PRDX6 | 3 | IVW (fe) | 0.416 | | 0.640 | 0.862 | 0.0% | 87.0% | 0.807 | yes | | Not_associated |

| | | | | Inverse variance weighted | | | | | Intercept | | Association | |
|---|---|---|---|---|---|---|---|---|---|---|---|---|
| **Exposure[a]** | **IVs[b]** | **Method** | **Beta** | **OR (95% CI)** | ***p*-value** | ***p*-FDR[c]** | **$I^2$** | **$I^2_{GX}$** | ***p*-value[d]** | **Consistent[e]** | **Nominal[f]** | **Classification[g]** |
| PRTN3 | 74 | IVW (fe) | 0.223 | 1.25 (0.94 - 1.66) | 0.119 | 0.489 | 0.0% | 97.1% | 0.439 | | | Not_associated |
| PYGL | 3 | IVW (fe) | 1.395 | | 0.066 | 0.415 | 31.0% | 96.7% | 0.355 | yes | | Not_associated |
| RCAN1 | 1 | Wald ratio | -0.495 | | 0.781 | 0.881 | | | | yes | | Not_associated |
| REG3A | 38 | IVW (fe) | -0.125 | 0.88 (0.58 - 1.34) | 0.558 | 0.862 | 0.0% | 96.2% | 0.962 | | | Not_associated |
| RNASE3 | 38 | IVW (fe) | 0.125 | 1.13 (0.80 - 1.60) | 0.474 | 0.862 | 0.0% | 98.2% | 0.113 | | | Not_associated |
| ROR1 | 29 | IVW (mre) | -0.188 | 0.83 (0.51 - 1.34) | 0.441 | 0.862 | 8.8% | 97.0% | 0.055 | | | Not_associated |
| SAA1 | 43 | IVW (fe) | -0.099 | 0.91 (0.66 - 1.24) | 0.536 | 0.862 | 0.0% | 98.8% | 0.214 | | | Not_associated |
| SEMA3G | 17 | IVW (mre) | 0.131 | 1.14 (0.59 - 2.19) | 0.694 | 0.862 | 20.9% | 96.8% | 0.210 | | | Not_associated |
| SERPINA10 | 112 | IVW (mre) | -0.045 | 0.96 (0.79 - 1.16) | 0.649 | 0.862 | 0.9% | 99.0% | 0.930 | | | Not_associated |
| SERPINF2 | 15 | IVW (mre) | 0.777 | 2.18 (0.96 - 4.94) | 0.063 | 0.415 | 2.4% | 93.2% | 0.245 | | | Not_associated |
| SERPING1 | 53 | IVW (fe) | -0.497 | 0.61 (0.48 - 0.77) | $2.6 \times 10^{-5}$ | $1.6 \times 10^{-3}$ | 0.0% | 98.5% | 0.581 | yes | IVW;WM;MRPRESSO;RAPS | Causal_robust |

| | | | | | | | | | | | | |
|---|---|---|---|---|---|---|---|---|---|---|---|---|
| | Inverse variance weighted | | | | | | Intercept | | | Association | | |
| **Exposure**[a] | **IVs**[b] | **Method** | **Beta** | **OR (95% CI)** | ***p*-value** | ***p*-FDR**[c] | $I^2$ | $I^2_{GX}$ | ***p*-value**[d] | **Consistent**[e] | **Nominal**[f] | **Classification**[g] |
| SFTPD | 54 | IVW (fe) | 0.121 | 1.13 (0.90 - 1.41) | 0.284 | 0.802 | 0.0% | 99.2% | 0.788 | | | Not_associated |
| SMPDL3A | 12 | IVW (fe) | -0.345 | 0.71 (0.36 - 1.41) | 0.326 | 0.862 | 0.0% | 97.9% | 0.825 | | | Not_associated |
| SNCA | 1 | Wald ratio | -1.513 | | 0.507 | 0.862 | | | | yes | | Not_associated |
| SORD | 19 | IVW (fe) | 0.673 | 1.96 (1.03 - 3.72) | $3.9 \times 10^{-2}$ | 0.359 | 0.0% | 96.9% | 0.665 | yes | IVW;MRPRESSO;RAPS | Causal_likely |
| VAT1 | 3 | IVW (fe) | 0.098 | | 0.939 | 0.951 | 0.0% | 63.8% | 0.435 | yes | | Not_associated |

[a] Exposure abbreviations are defined in **eTable 1**. [b] Number of instrumental variables (IV). [c] *p*-value adjusted by false discovery rate (FDR). [d] *p*-value of MR Egger intercept. [e] A consistent association was considered when the direction of all methods was the same. [f] Methods with a significant *p*-value (<0.05). [g] Result of the primary analysis.

**Table 5**: Result of the colocalization analysis

| | Location[b] | | | | Posterior probabilities[c] | | | | |
|---|---|---|---|---|---|---|---|---|---|
| **Gene[a]** | **Chromosome** | **Initial** | **Final** | **Common SNPs** | **H0** | **H1** | **H2** | **H3** | **H4** |
| **GBS** | | | | | | | | | |
| CFHR4 | 1 | 196,350,241 | 197,418,972 | 2,218 | 0.00 | 0.73 | 0.00 | 0.21 | 0.06 |
| CNDP1 | 18 | 74,034,440 | 75,087,212 | 3,029 | 0.00 | 0.62 | 0.00 | 0.23 | 0.15 |
| CRISPLD2 | 16 | 84,319,984 | 85,420,768 | 5,979 | 0.00 | 0.60 | 0.00 | 0.34 | 0.07 |
| FCGR2B | 1 | 161,163,147 | 162,178,654 | 3,332 | 0.00 | 0.65 | 0.00 | 0.24 | 0.11 |
| GSTM4 | 1 | 109,156,081 | 110,174,836 | 2,344 | 0.00 | 0.75 | 0.00 | 0.21 | 0.04 |
| MMP8 | 11 | 102,211,795 | 103,227,050 | 3,175 | 0.00 | 0.71 | 0.00 | 0.20 | 0.09 |
| PENK | 8 | 55,936,674 | 56,946,734 | 3,166 | 0.00 | 0.74 | 0.00 | 0.22 | 0.04 |
| SERPING1 | 11 | 57,097,387 | 58,114,853 | 2,313 | 0.00 | 0.57 | 0.00 | 0.15 | 0.28 |
| SORD | 15 | 44,523,104 | 45,577,185 | 2,256 | 0.00 | 0.78 | 0.00 | 0.16 | 0.06 |

[a] Abbreviations are defined in **Table 1**. [b] Genome locations are referred to GRCh38 build. [c] **Posterior probabilities**: H0, neither trait has a genetic association in the region; H1 = only exposure has a genetic association in the region; H2 = only outcome has a genetic association in the region; H3 = both traits are associated with the same genomic region, but with different, independent variants; H4 = both traits are associated and share a single causal variant.

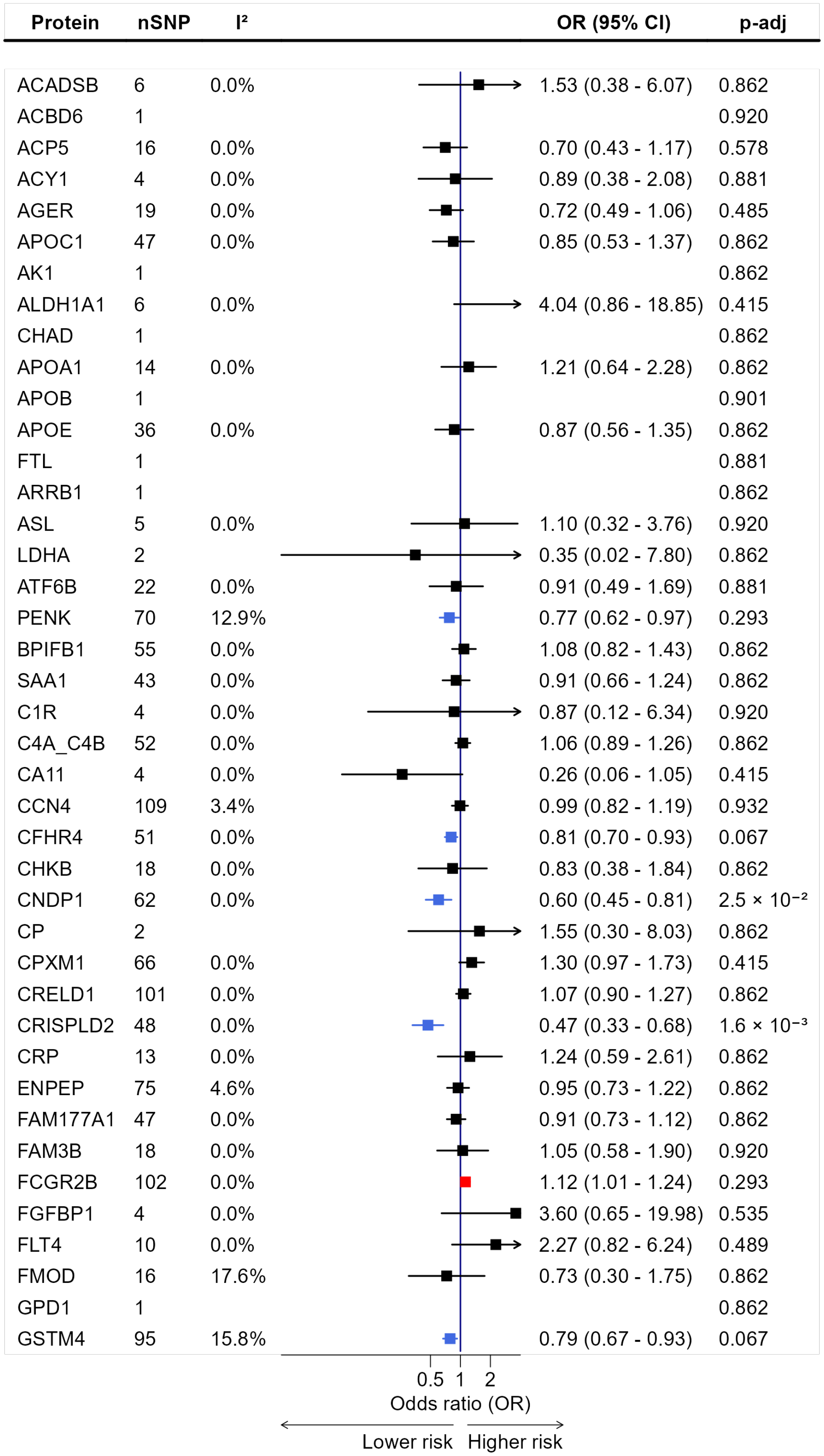

Protein nSNP I² OR (95% CI) p-adj
ACADSB 6 0.0% 1.53 (0.38 - 6.07) 0.862
ACBD6 1 0.920
ACP5 16 0.0% 0.70 (0.43 - 1.17) 0.578
ACY1 4 0.0% 0.89 (0.38 - 2.08) 0.881
AGER 19 0.0% 0.72 (0.49 - 1.06) 0.485
APOC1 47 0.0% 0.85 (0.53 - 1.37) 0.862
AK1 1 0.862
ALDH1A1 6 0.0% 4.04 (0.86 - 18.85) 0.415
CHAD 1 0.862
APOA1 14 0.0% 1.21 (0.64 - 2.28) 0.862
APOB 1 0.901
APOE 36 0.0% 0.87 (0.56 - 1.35) 0.862
FTL 1 0.881
ARRB1 1 0.862
ASL 5 0.0% 1.10 (0.32 - 3.76) 0.920
LDHA 2 0.35 (0.02 - 7.80) 0.862
ATF6B 22 0.0% 0.91 (0.49 - 1.69) 0.881
PENK 70 12.9% 0.77 (0.62 - 0.97) 0.293
BPIFB1 55 0.0% 1.08 (0.82 - 1.43) 0.862
SAA1 43 0.0% 0.91 (0.66 - 1.24) 0.862
C1R 4 0.0% 0.87 (0.12 - 6.34) 0.920
C4A_C4B 52 0.0% 1.06 (0.89 - 1.26) 0.862
CA11 4 0.0% 0.26 (0.06 - 1.05) 0.415
CCN4 109 3.4% 0.99 (0.82 - 1.19) 0.932
CFHR4 51 0.0% 0.81 (0.70 - 0.93) 0.067
CHKB 18 0.0% 0.83 (0.38 - 1.84) 0.862
CNDP1 62 0.0% 0.60 (0.45 - 0.81) 2.5 × 10⁻²
CP 2 1.55 (0.30 - 8.03) 0.862
CPXM1 66 0.0% 1.30 (0.97 - 1.73) 0.415
CRELD1 101 0.0% 1.07 (0.90 - 1.27) 0.862
CRISPLD2 48 0.0% 0.47 (0.33 - 0.68) 1.6 × 10⁻³
CRP 13 0.0% 1.24 (0.59 - 2.61) 0.862
ENPEP 75 4.6% 0.95 (0.73 - 1.22) 0.862
FAM177A1 47 0.0% 0.91 (0.73 - 1.12) 0.862
FAM3B 18 0.0% 1.05 (0.58 - 1.90) 0.920
FCGR2B 102 0.0% 1.12 (1.01 - 1.24) 0.293
FGFBP1 4 0.0% 3.60 (0.65 - 19.98) 0.535
FLT4 10 0.0% 2.27 (0.82 - 6.24) 0.489
FMOD 16 17.6% 0.73 (0.30 - 1.75) 0.862
GPD1 1 0.862
GSTM4 95 15.8% 0.79 (0.67 - 0.93) 0.067
0.5 1 2
Odds ratio (OR)
Lower risk Higher risk

| Protein | nSNP | I² | OR (95% CI) | p-adj |
|---|---|---|---|---|
| HNRNPAB | 2 | | 1.97 (0.12 - 31.52) | 0.862 |
| IBSP | 6 | 25.6% | 0.74 (0.13 - 4.10) | 0.862 |
| IDH1 | 8 | 0.0% | 0.61 (0.32 - 1.16) | 0.526 |
| IGFBP1 | 3 | | 0.38 (0.05 - 2.99) | 0.862 |
| IGL_IGHD_IGK | 2 | | 1.61 (0.19 - 13.60) | 0.862 |
| INHBC | 5 | 0.0% | 0.45 (0.10 - 2.01) | 0.811 |
| ITIH4 | 8 | 29.1% | 0.36 (0.08 - 1.58) | 0.578 |
| KLB | 68 | 0.0% | 1.14 (0.95 - 1.36) | 0.560 |
| LAG3 | 7 | 0.0% | 1.01 (0.35 - 2.93) | 0.985 |
| LAP3 | 5 | 0.0% | 0.52 (0.11 - 2.56) | 0.862 |
| LGALS3BP | 20 | 0.0% | 0.90 (0.52 - 1.56) | 0.862 |
| LGMN | 24 | 0.0% | 1.65 (0.91 - 2.98) | 0.485 |
| MMP13 | 1 | | | 0.862 |
| MMP8 | 18 | 0.0% | 1.90 (1.05 - 3.44) | 0.339 |
| MST1 | 28 | 0.0% | 0.91 (0.78 - 1.06) | 0.702 |
| MUSK | 3 | | 2.81 (0.29 - 27.59) | 0.862 |
| MX1 | 44 | 0.0% | 1.21 (0.79 - 1.85) | 0.862 |
| NANS | 7 | 0.0% | 0.44 (0.11 - 1.74) | 0.728 |
| NCF1 | 17 | 0.0% | 0.90 (0.61 - 1.32) | 0.862 |
| OLFML3 | 4 | 0.0% | 2.59 (0.50 - 13.31) | 0.745 |
| PAM | 29 | 13.2% | 1.08 (0.84 - 1.39) | 0.862 |
| PCSK9 | 22 | 0.0% | 0.64 (0.37 - 1.11) | 0.489 |
| PGLYRP1 | 12 | 0.0% | 2.15 (0.94 - 4.88) | 0.415 |
| PKM | 1 | | | 0.901 |
| PPA1 | 19 | 6.3% | 1.16 (0.57 - 2.37) | 0.862 |
| PRDX6 | 3 | | 1.52 (0.27 - 8.67) | 0.862 |
| PRTN3 | 74 | 0.0% | 1.25 (0.94 - 1.66) | 0.489 |
| PYGL | 3 | | 4.04 (0.91 - 17.87) | 0.415 |
| RCAN1 | 1 | | | 0.881 |
| REG3A | 38 | 0.0% | 0.88 (0.58 - 1.34) | 0.862 |
| RNASE3 | 38 | 0.0% | 1.13 (0.80 - 1.60) | 0.862 |
| ROR1 | 29 | 8.8% | 0.83 (0.51 - 1.34) | 0.862 |
| SEMA3G | 17 | 20.9% | 1.14 (0.59 - 2.19) | 0.862 |
| SERPINA10 | 112 | 0.9% | 0.96 (0.79 - 1.16) | 0.862 |
| SERPINF2 | 15 | 2.4% | 2.18 (0.96 - 4.94) | 0.415 |
| SERPING1 | 53 | 0.0% | 0.61 (0.48 - 0.77) | $1.6 \times 10^{-3}$ |
| SFTPD | 54 | 0.0% | 1.13 (0.90 - 1.41) | 0.802 |
| SMPDL3A | 12 | 0.0% | 0.71 (0.36 - 1.41) | 0.862 |
| SNCA | 1 | | | 0.862 |
| SORD | 19 | 0.0% | 1.96 (1.03 - 3.72) | 0.359 |
| VAT1 | 3 | | 1.10 (0.09 - 13.80) | 0.951 |

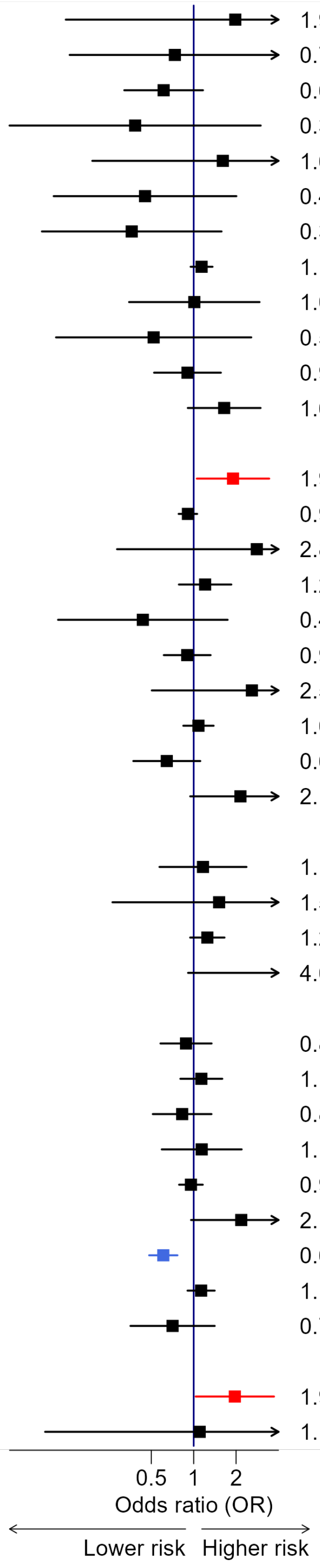

**Figure 1**: Causal effects of proteins on Guillain-Barré syndrome.

**eREFERENCES**

1. Skrivankova VW, Richmond RC, Woolf BAR, et al. Strengthening the Reporting of Observational Studies in Epidemiology Using Mendelian Randomization: The STROBE-MR Statement. *JAMA* 2021;326:1614 doi:10.1001/jama.2021.18236.

2. Eldjarn GH, Ferkingstad E, Lund SH, et al. Large-scale plasma proteomics comparisons through genetics and disease associations. *Nature* 2023;622:348–358 doi:10.1038/s41586-023-06563-x.

3. Zheng J, Haberland V, Baird D, et al. Phenome-wide Mendelian randomization mapping the influence of the plasma proteome on complex diseases. *Nature Genetics* 2020;52:1122–1131 doi:10.1038/s41588-020-0682-6.

4. EPIC- InterAct Consortium, Burgess S, Scott RA, Timpson NJ, Davey Smith G, Thompson SG. Using published data in Mendelian randomization: A blueprint for efficient identification of causal risk factors. *European Journal of Epidemiology* 2015;30:543–552 doi:10.1007/s10654-015-0011-z.

5. Burgess S, Davey Smith G, Davies NM, et al. Guidelines for performing Mendelian randomization investigations: Update for summer 2023. *Wellcome Open Research* 2023;4:186 doi:10.12688/wellcomeopenres.15555.3.

6. Bowden J, Davey Smith G, Haycock PC, Burgess S. Consistent Estimation in Mendelian Randomization with Some Invalid Instruments Using a Weighted Median Estimator. *Genetic Epidemiology* 2016;40:304–314 doi:10.1002/gepi.21965.

7. Bowden J, Davey Smith G, Burgess S. Mendelian randomization with invalid instruments: Effect estimation and bias detection through Egger regression. *International Journal of Epidemiology* 2015;44:512–525 doi:10.1093/ije/dyv080.

8. Zhao Q, Wang J, Hemani G, Bowden J, Small DS. Statistical inference in two-sample summary-data Mendelian randomization using robust adjusted profile score. *The Annals of Statistics* 2020;48 doi:10.1214/19-AOS1866.

9. Cochran WG. The Combination of Estimates from Different Experiments. *Biometrics* 1954;10:101 doi:10.2307/3001666. Accessed at: https://www.jstor.org/stable/3001666. Accessed March 10, 2026.

10. Higgins JPT, Thompson SG. Quantifying heterogeneity in a meta-analysis. *Statistics in Medicine* 2002;21:1539–1558 doi:10.1002/sim.1186.

11. Bowden J, Del Greco M. F, Minelli C, Davey Smith G, Sheehan NA, Thompson JR. Assessing the suitability of summary data for two-sample Mendelian randomization analyses using MR-Egger regression: The role of the I2 statistic. *International Journal of Epidemiology* Epub 2016 Sep.:dyw220 doi:10.1093/ije/dyw220.

12. Verbanck M, Chen C-Y, Neale B, Do R. Detection of widespread horizontal pleiotropy in causal relationships inferred from Mendelian randomization between complex traits and diseases. *Nature Genetics* 2018;50:693–698 doi:10.1038/s41588-018-0099-7.

13. Wang G, Sarkar A, Carbonetto P, Stephens M. A Simple New Approach to Variable Selection in Regression, with Application to Genetic Fine Mapping. *Journal of the Royal Statistical Society Series B: Statistical Methodology* 2020;82:1273–1300 doi:10.1111/rssb.12388.

14. Team RC. R: A Language and Environment for Statistical Computing. 2017.

15. RStudio-Team. RStudio: Integrated Development for R. 2022.

16. Hemani G, Zheng J, Elsworth B, et al. The MR-Base platform supports systematic causal inference across the human phenome. *eLife* 2018;7:e34408 doi:10.7554/eLife.34408.
